# Examining the Impact of Increasing Vaccine Coverage and Nonpharmaceutical Interventions against Coronavirus Disease 2019 In Ghana using Mathematical Modeling

**DOI:** 10.1101/2022.07.09.22277456

**Authors:** Sylvia K. Ofori, Jessica S. Schwind, Kelly L. Sullivan, Gerardo Chowell, Benjamin J. Cowling, Isaac Chun-Hai Fung

## Abstract

Seroprevalence studies assessing community exposure to severe acute respiratory syndrome coronavirus 2 (SARS-CoV-2) in Ghana concluded that population-level immunity remained low as of February 2021. Thus, it is important to demonstrate how increasing vaccine coverage reduces the economic and public health impacts associated with transmission of the novel coronavirus. To that end, this study used a Susceptible-Exposed-Presymptomatic-Symptomatic-Asymptomatic-Recovered-Dead-Vaccinated compartmental model to simulate coronavirus disease 2019 (COVID-19) transmission and the role of public health interventions in Ghana. The impact of increasing vaccination rate and decline in transmission rates due to nonpharmaceutical interventions (NPIs) on cumulative infections and deaths averted was explored under different scenarios. Latin hypercube sampling-partial rank correlation coefficient (LHS-PRCC) was used to investigate uncertainty and sensitivity of the outcomes to the parameters. Simulation results suggest that increasing the vaccination rate to achieve 50% coverage was associated with almost 30,000 deaths and 25 million infections averted. In comparison, a 50% decrease in the transmission coefficient was associated with about 50 million infections and 120,000 deaths averted. The LHS-PRCC results also found that cumulative infections and deaths averted were most sensitive to three model parameters: Transmission rate, vaccination rate, and waning immunity rate from infection. There is a need to increase vaccination coverage by ensuring an increased supply. Adherence to NPIs and increased vaccine uptake would successfully mitigate the impact of COVID-19 in Ghana.

## Introduction

The first cases of coronavirus disease 2019 (COVID-19) in Ghana, a lower-middle-income country, were reported on March 12, 2020.^1^ Among countries in West Africa, Ghana serves as a role model in economic development and democratic governance.^2^ Ghana’s healthcare system is relatively well-resourced in the region and thus provides the infrastructure needed to implement a COVID-19 vaccination campaign nationwide. Sibiri and colleagues reported a timely response and innovative measures such as the COVID-19 tracker app and the ‘pool testing’ technique were taken in Ghana to curb the widespread transmission of COVID-19.^3^ Ghana also led continent in terms of the number of COVID-19 tests conducted per million people, utilizing enhanced and technologically advanced contact tracing and testing as well as drone technology to deliver samples to laboratories.^3^

Before December 2020, when vaccines against severe acute respiratory syndrome coronavirus 2 (SARS-CoV-2) were absent, the implementation of significant nonpharmaceutical interventions (NPIs) was required to delay the global spread of the virus in order to give time for vaccines to be developed. Some of the NPIs enacted in Ghana included a 14-day mandatory quarantine for all persons entering the country, school and church closures, a lockdown of the major cities in the country, and internal movement restrictions.^4^ With the approval of COVID-19 vaccines and other vaccine candidates under development, policymakers and public health officials needed to plan and assess a vaccination rollout plan that would significantly reduce the number of cases and fatalities, especially in lower-and-middle-income economies like Ghana.

Even with the carefully planned interventions implemented to meet the objectives set by the government, compliance with these measures reduced over time. Evidence from seroprevalence studies in Africa suggested that most SARS-CoV-2 infections were asymptomatic and affected more people than reported.^5–7^ An adjusted seroprevalence of 12.3% (95% CI: 8.2%- 16.5%) was reported among five hundred healthcare workers in Malawi between May 22 and June 19, 2020.^5^ In Nigeria, Majiya and colleagues reported a seroprevalence of 25.41% for immunoglobulin G (IgG) and 2.16% for immunoglobulin M (IgM) between June 26 and June 30, 2020.^8^ A seroprevalence of 5.2% (95% CI: 3.7%, 7.1%) was estimated among blood donors in Kenya between April 30 and June 16, 2020, where 90% of them were found to be asymptomatic.^6^ In Ghana, a study to estimate the community exposure to SARS-CoV-2 found 19% of the population tested positive for IgM, IgG, or both antibodies in August 2020.^7^ All four studies concluded most of the populations remained susceptible, leaving the country at risk of major outbreaks. The Ghana study was repeated in 2021 at major malls and a hospital, with reported seroprevalence values of 27% and 25%, respectively,^7^ reiterating vaccine uptake in a larger percentage of the population is needed and reinforcing the need to implement NPIs that have been proven effective.

Ghana was the first country to receive 600,000 doses of the AstraZeneca/Oxford vaccine from the Serum Institute of India, which arrived on February 24, 2021, through the COVAX program.^9^ The AstraZeneca/Oxford vaccine is a recombinant adenoviral vector containing SARS-CoV-2 structural surface glycoprotein antigen with an overall efficacy of 70.4% in initial clinical trials given in two doses over 8-12 weeks intervals.^10, 11^ COVAX is led by Gavi, the Vaccine Alliance, the World Health Organization, and the Coalition for Epidemic Preparedness Innovations (CEPI), in addition to other organizations.^12^ The rollout plan for the vaccination program prioritized those older than 60 years, those with underlying severe health issues, and essential workers (health care professionals, teachers, police, and journalists).^13^ The Ghana Food and Drugs Authority (FDA) also authorized the Sputnik V vaccines for emergency use. As of October 2021, about 2.8% of Ghanaians had received two doses of the COVID-19 vaccines.^14^ Vaccines used in the country as of October 2021 are AstraZeneca, Sputnik-V, Moderna, and Jansen (Johnson and Johnson), with AstraZeneca accounting for more than 90% of vaccines administered.^1^

The transmission dynamics of infectious diseases have been well-studied with mathematical models. The compartmental “Susceptible-Infected-Recovered” (SIR) model proposed by Kermack and McKendrick in 1922 uses dynamical systems to track the progression of different epidemiological states over time.^15^ The SIR model and the SEIR (Susceptible-Exposed-Infected-Recovered) model with a compartment for individuals in the latent period have been extensively investigated for diseases like the plague and influenza.^16^ However, these simple models may not represent the natural history of COVID-19 adequately, especially the contribution to the transmission potential by asymptomatic and presymptomatic individuals.^17–19^ Chen and colleagues proposed a “Susceptible-Exposed-Infectious-Asymptomatic-Recovered” (SEIAR) model, which introduced a compartment to account for infectious individuals who do not show symptoms.^16^ However, their model did not account for the transmission before symptom onset. In this study, the Susceptible-Exposed-Presymptomatic-Symptomatic-Asymptomatic-Recovered-Dead-Vaccinated (SEPIARD-V) compartmental model is proposed to model the dynamics of SARS-CoV-2 and the role of interventions on deaths and cumulative infections.^20^

The SEPIARD-V model acknowledges that individuals who are initially asymptomatic and later develop symptoms transmit the virus while in the presymptomatic phase. Individual studies included in a systematic review on the transmission dynamics provided evidence of the presymptomatic transmission of COVID-19 1-3 days before symptom onset.^17^ The model is suitable for studying transmission dynamics of COVID-19 in Ghana due to the growing evidence that both symptomatic and asymptomatic patients transmit the infection.^21–23^

In this study, we used the SEPIARD-V model to assess the role of vaccination in averting deaths and cumulative infections due to COVID-19 in Ghana. We also explored the impact of NPIs in reducing deaths and cumulative infections due to COVID-19. Through simulations of counterfactuals, we explored scenarios with varying vaccination coverage and transmission reduction through timely NPI compliance. To identify the outcomes’ parameter sensitivity, we used Latin hypercube sampling (LHS) to explore the parameter space and the partial rank correlation coefficient (PRCC) to quantify its correlation.

## Methods

### Model formulation

In the SEPIARD-V model, the population is initially susceptible until an infectious individual is introduced. After contact with an infectious person, we assume susceptible individuals are infected and move to the latent period (E), infected but not transmitting the virus. Exposed individuals can either be asymptomatically (A) or presymptomatically infectious (P). Presymptomatic infectious individuals become symptomatic (I) before recovery (R). Asymptomatic individuals move to the recovery compartment without showing symptoms.^24, 25^ A death compartment (D) was included to account for symptomatic infectious individuals who die from COVID-19. Susceptible individuals who are vaccinated with 2 doses of AstraZeneca vaccine (V) stay immune and are protected from infection for a period until they return to be susceptible due to waning immunity. Recovered individuals enjoy temporary immunity.

### Model equations

The total population of Ghana, N, is assumed to be a constant over the study’s time frame and equals the sum of S, E, A, P, I, R, D, and V at any time (Eq. 1). Birth and death from non-COVID-19 causes are excluded from the model as they are assumed to not affect the infection dynamics in the population over the study period.

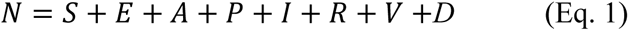

The model also assumes homogenous mixing, which means each individual has the same chance of interacting with others and becoming infected. Susceptible individuals may become infected after contact with presymptomatic, asymptomatic, and symptomatic persons with a force of infection, λ. Given a frequency-dependent model formulation, λ is directly proportional to the prevalence of the infection (the probability of having an infectious contact), the number of contacts a susceptible individual is expected to make per unit time, and the probability of successful infection given a contact (Eq. 2). The product of the latter two is represented as the transmission coefficient, β. Since the probability of successful infection is lower for asymptomatic or presymptomatic contacts than for symptomatic contacts, two parameters, u and r, were introduced to reflect this reality, respectively, with 0<u<1 and 0<r<1.^26^

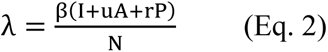

Susceptible individuals are infected at a rate of λ (Eq. 3). The mean of the latent period is 1/*k* (Eq.4). At the end of their latent period, a proportion (δ) of individuals become pre-symptomatically infected at a rate of *k* (Eq. 5), and the rest (1-δ) become asymptomatic at the same rate of *k* (Eq.6).^16, 27^ Asymptomatic individuals recover at a rate of *q* (Eq.6 and Eq.8). Presymptomatic individuals become symptomatic at a rate of *c* (Eq. 7). The mean symptomatic period is 1/*f* (Eq. 7). A proportion, *z,* of symptomatic individuals may die from COVID-19 while the rest (1-*z*) will recover (Eq. 8 and Eq. 9). Susceptible individuals become fully vaccinated at a daily rate of *v* with a 2-dose vaccine efficacy of *σ* (Eq. 10). Thus, susceptible individuals are vaccinated and become immune from infection at a rate of *vσ,* and individuals become susceptible again at a rate of χ once vaccine-induced immunity wanes (Eq. 3 and Eq. 10). Recovered individuals stay immune from infection for a mean duration of 1/*w*. Once their natural infection-induced immunity wanes, they become susceptible again (Eq. 3 and Eq. 8).

The model equations are summarized below:

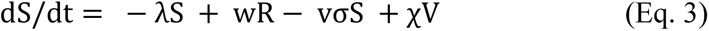

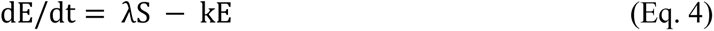

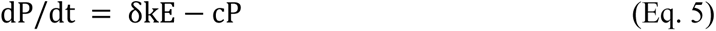

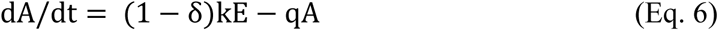

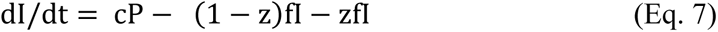

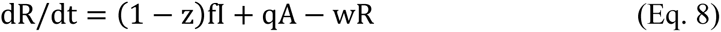

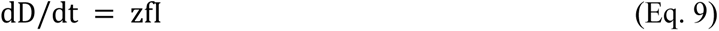

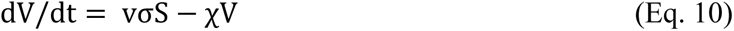

The rate of change in cumulative case count is the product of the number of individuals in the latent period (E) and their rate of becoming infectious (k).

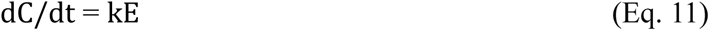

The model’s Ordinary Differential Equations (ODE) system was solved following the Runge-Kutta 4 method in the deSolve package in R version 4.1.1. ^28^

### Model parameters

The parameters for the model were obtained from published literature and summarized in Table 1. The latent period (1/*k*) has a mean of 1.85 days and ranges from 1 to 3 days.^29, 30^ Presymptomatic period, 1/*c*, is assumed to be 2.9 days on average and ranges from 1.2 to 8.2 days.^31^ The mean duration of infectiousness is 15.7 days (95% CI: 13, 18.4) and 7.25 days (95% CrI: 5.1, 8.69) for symptomatic individuals (1/*f*) and asymptomatic individuals (1/*h*), respectively.^32–35^ The transmission coefficient, β, is estimated from the basic reproduction number (R_0_), which is assumed to be 3.13 (95% CrI: 1.24, 6.35) as reported by Armachie and colleagues for the initial strain in Ghana.^36^ According to the Centers for Disease Control and Prevention (CDC) COVID-19 pandemic planning scenarios, the relative transmissibility of asymptomatic and presymptomatic individuals, *u* and *r*, are assumed to be 0.75, respectively.^37^ The vaccination rate, *v*, is back-calculated, assuming that 2.8% of the population were fully vaccinated, giving a daily vaccination rate of 0.0003519.^14^ Two doses of AstraZeneca COVID-19 vaccine were reported to have an efficacy (σ) of 0.745 (95% CrI: 0.684, 0.794) against infection.^10, 38^ Immunity is acquired from either natural infection or vaccination. In the base case scenario, vaccination-induced immunity offers protection from infection for six months (1/χ = 180 days), while immunity from natural infection is about one year (1/*w* = 365 days).^39^ Finally, the case fatality ratio, *z*, is the proportion of symptomatic individuals who die from SARS-CoV-2.^40^

**Table 1:** Estimated Parameter Values for the SEPIARD-V COVID-19 Model.

| Parameter | Symbol | Point estimate | Range/95% CI/SD * | Distribution | Reference |
| --- | --- | --- | --- | --- | --- |
| Proportion of susceptible individuals infected at the start of the simulation |  | 5,700,000<br>(Seroprevalence of 19% as of August 2020) | N/A | N/A | Quashie et al.,<sup>7</sup> |
| Mean latency period (the period from exposure to infectiousness) | $1/k$ | 1.85 days | 1-3 days | Gamma | Abbasi et al.,<sup>57</sup>,<br>Liu et al.<sup>30</sup> |
| Infectious period for symptomatic persons | $1/f$ | 15.7 days | 6.7 days (SD) | Gamma | Byrne et al.,<sup>33</sup><br>Cai et al.,<sup>32</sup><br>Xing et al.,<sup>35</sup> |
| Infectious period for asymptomatic persons | $1/q$ | 7.25 days | 5.91, 8.69 days | Gamma | Ma et al.,<sup>58</sup>,<br>Byrne et al.<sup>33</sup> |
| Infectious period for presymptomatic to symptomatic | $1/c$ | 2.9 days | 1.2-8.2 days | Gamma | Tindale et al.,<sup>31</sup>,<br>Byrne et al.,<sup>33</sup> |
| Basic reproduction number | $R_0$ | 3.13 | 1.4-3.5 | Uniform | Armachie et al.,<sup>36</sup><br>Dwumoh et al.,<sup>59</sup> |
| Infection rate | $\beta$ | 0.302 | 0.15 (SD) | Gamma | Estimated |
| Decline in transmission rate | $l$ | 0 | 0-1 | N/A | Assumed |
| Probability of exposed person becoming pre-symptomatically infected | $\delta$ | 0.30 | 0.26-0.37 | Uniform | Chen et al.,<sup>16</sup><br>Buitrago-Garcia et al.,<sup>27</sup> |
| Vaccine efficacy | $\sigma$ | 0.745 | 0.6-0.8 | Uniform | Bernal et al.,<sup>38</sup> |
| Proportion of the population vaccinated with two doses per day per capita | $v$ | 0.0003519<br>2.8 per 100 people over 80 days. | 0- 0.0042<br>(max value assuming 50% of the population receives two doses) | Uniform | Mathieu et al.,<sup>60</sup> |
| Relative transmissibility of asymptomatic individuals | $u$ | 0.75<sup>a</sup> | 0.25-1 | Gamma | CDC<sup>37</sup> |
| Relative transmissibility of presymptomatic individuals | $r$ | 0.75<sup>a</sup> | 0.25-1 | Gamma | CDC<sup>37</sup> |
| Rate at which immunity wanes for vaccinated individuals | $\chi$ | 0.0055 | 0.0018-0.033 | Uniform | Lacobucci<sup>61</sup> |
| Rate at which immunity wanes from natural infection | $w$ | 0.0027 | 0.0018-0.033 | Uniform | Good & Hawke,<sup>39</sup> |
| Case fatality rate | $z$ | 0.009 | 0.0061-0.0092 | Uniform | Our world in Data (Range from January 26 and November 12, 2021) |

CI, confidence interval. SD, standard deviation. \*This is a range if the distribution is uniform. This is a 95% CI if the distribution is gamma.

### Initialization of the model simulation

As an approximation to the population size of Ghana, N is set to 30,800,000.^41^ Given that 19% of the population was infected with SARS-CoV-2 at the initiation of the simulation (August 2020), 5,852,000 individuals were in the R compartment initially, leaving the S compartment with 24,948,000 individuals. We also assumed that for the base case scenario, there was one symptomatic index case at the beginning of the simulation (I=1, A=0, P=0, D=0, and V=0). The model was run for 500 days to allow enough time for the first wave of the epidemic to subside and observe when the second wave began to emerge.

### The basic reproduction number (R_0_)

The basic reproduction number (R_0_) is an epidemiological measure for the spread of an infectious disease. It is defined as the average number of secondary cases generated by an infectious individual in a totally susceptible population in the absence of behavioral changes or public health interventions.^26^ Given that R_0_ is the average number of susceptible individuals infected by an infectious person when introduced into a totally susceptible population, a value greater than one implies that the spread of the epidemic continues. In contrast, a value less than one means the epidemic will have limited spread before it dies out.^42^

### The R_0_ For the SEPIARD-V model

The equation for deriving R_0_ is explained here. ^20, 43^ The R_0_ contribution from each infectious compartment is the product of the transmission rate, the proportion of the population entering that compartment, and the average time spent in the compartment. As mentioned above, asymptomatic, presymptomatic, and symptomatic individuals contribute to the infection process. The transmissibility coefficient for symptomatic individuals is *β*. The relative transmissibility of asymptomatic and presymptomatic individuals is a product of *u* and *β*, and *r* and *β*, respectively.

The proportion of infected individuals who become presymptomatic and then symptomatic is *δ,* and the proportion of infected individuals who become asymptomatic is 1-*δ*.

The average time spent in the asymptomatic compartment is 1/*q*, the presymptomatic compartment is 1/*c*, and the symptomatic compartment is 1/*f*.

Thus, given that R_0_ is the sum of R_0_ contribution from each of the infectious compartments,

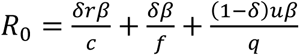

Hence, 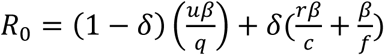.^20^

### Scenario analysis using the homogeneous mixing model

The first scenario we assessed was the impact of reducing transmission rates on symptomatic infections, cumulative deaths, and cumulative infections. We introduced a new parameter, *l*, where 0<*l*<1, to account for the extent of the decline in the transmission. To account for the decrease in transmission, λ is updated as:

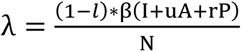

In the base case scenario, *l*=0, there was no change in transmission rate. Next, we assumed that NPIs result in a 20% reduction in β, followed by 30%, 40%, and 50%. The peak magnitude and timing for each decline were assessed and compared. We also identified which declines delayed the peak beyond 500 days.

Secondly, we varied the daily vaccination rate of COVID-19. We explored the impact on the peak of symptomatic infections and the cumulative number of deaths and infections in the population. The base case scenario was the trajectory of the pandemic, assuming vaccines were not available. The next option was using the daily vaccination rate as of October 2021, when this study was completed. We also modeled the effect of doubling the recent vaccination coverage, which was 5.6%. The last two assumptions in this scenario used daily vaccination rates needed to achieve 20% and 50% vaccination coverage over four months.

Next, we investigated the cumulative number of deaths and infections averted due to the effectiveness of NPIs on declining transmission rates in Ghana. The model’s behavior was explored over a range of *l* values from 0 to 1. The baseline scenario was when all other parameter values were held constant. We repeated the simulation by varying and increasing vaccination rates to achieve 50% coverage. The number of new infections and deaths per day was calculated from the model to examine the total fatalities and infections averted.

Then, we assessed the impact of reducing transmission rates and increasing vaccination rates on the deaths and cumulative infections averted.

### Uncertainty analysis using the homogeneous mixing model

The robustness of the outcomes of our model was assessed through an uncertainty analysis using Latin Hypercube Sampling (LHS). The LHS method is a stratified random sampling method implemented by dividing a range of values for a given parameter into equal intervals of probability density and samples taken without replacement.^44^ Using the LHS allows for unbiased estimates of our modeling outcomes using fewer simulations.^45^ The distribution of each parameter used in the LHS can be found in Table 1. Thirteen uncertain parameters were identified in our model, and 500 simulations were run. Thus, the parameter space for the LHS simulation was made of 6500 values (13 dimensions by 500 samples). Ranges or confidence intervals for each parameter used in populating the LHS matrix can be found in Table 1. The uncertainty analysis was repeated for a) varying percentage reduction in transmission rates (*l*) and b) varying the daily vaccination rates to achieve 50% coverage over four months (*v*). For each run, the total infections and deaths averted were calculated. The results of the LHS uncertainty analysis were summarized with box and whisker plots.

### Global sensitivity analysis using Partial Rank Correlation Analysis with the homogeneous mixing model

Sensitivity analyses were performed to identify the parameters that were influential on our outcomes using the Partial Rank Correlation method with the function pcc() in the R package ‘sensitivity.’^46^ The PRCCs and the associated 95% confidence intervals were estimated to assess the level of influence. Generally, PRCC values close to +1 or -1 strongly influence the outcome, and confidence intervals that do not include zero mean significant impact. The sign of the ranked coefficient indicates the qualitative relationship where a positive sign shows the LHS parameter is directly proportional to the outcome measure.^47^

### Programming Language and Code

All simulations were conducted in R version 4.1.1 (R Core Team, R Foundation for Statistical Computing, Vienna, Austria). The R code is provided in the Online Supplementary Materials.

### Ethics

This project was determined to be exempt from full review by Georgia Southern University’s Institution Review Board (H20364) under the G8 exemption category (non-human subjects determination) under Code of Federal Regulations Title 45 Part 46.

## Results

### Impact of Reduced Transmission Rates Due to NPIs on Symptomatic Infections, Cumulative Deaths, and Cumulative Infections

The findings of our study indicated NPIs that decrease transmission rates by at least 40% delayed the peak of symptomatic infections beyond 500 days. In the base case scenario, the expected total deaths from COVID-19 were almost 40,000 in the population by the end of the first year with the number of deaths increasing again after 400 days. A 20% decline in transmission rate was associated with a decrease in approximately 20,000 deaths by the end of the first year. Further reduction in transmission rates was followed by substantive decreases in the number of expected deaths (Figure 2).

**Figure 1:**
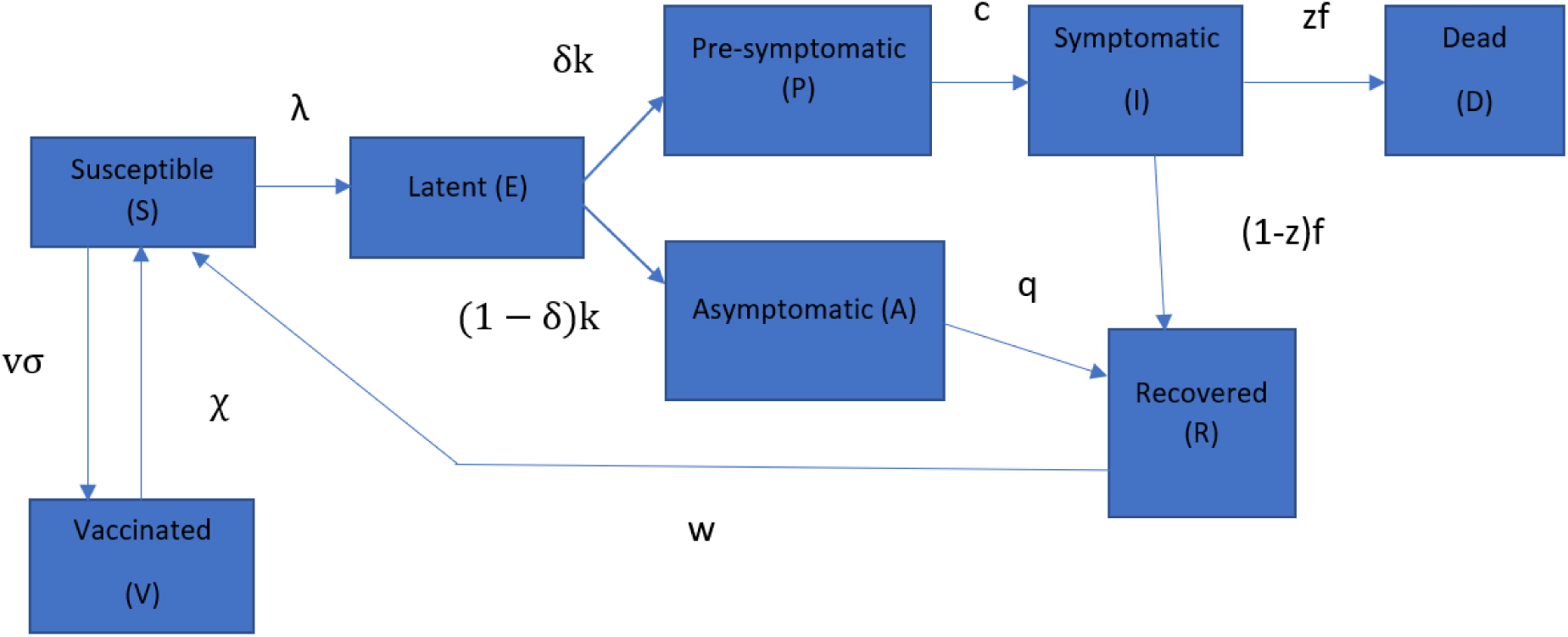
Flow diagram of the Susceptible-Exposed-Presymptomatic-Symptomatic-Asymptomatic Recovered-Dead-Vaccinated (SEPIARD-V) Model used to represent COVID-19 infection, disease progress, and vaccination of individuals in Ghana.

**Figure 2:**
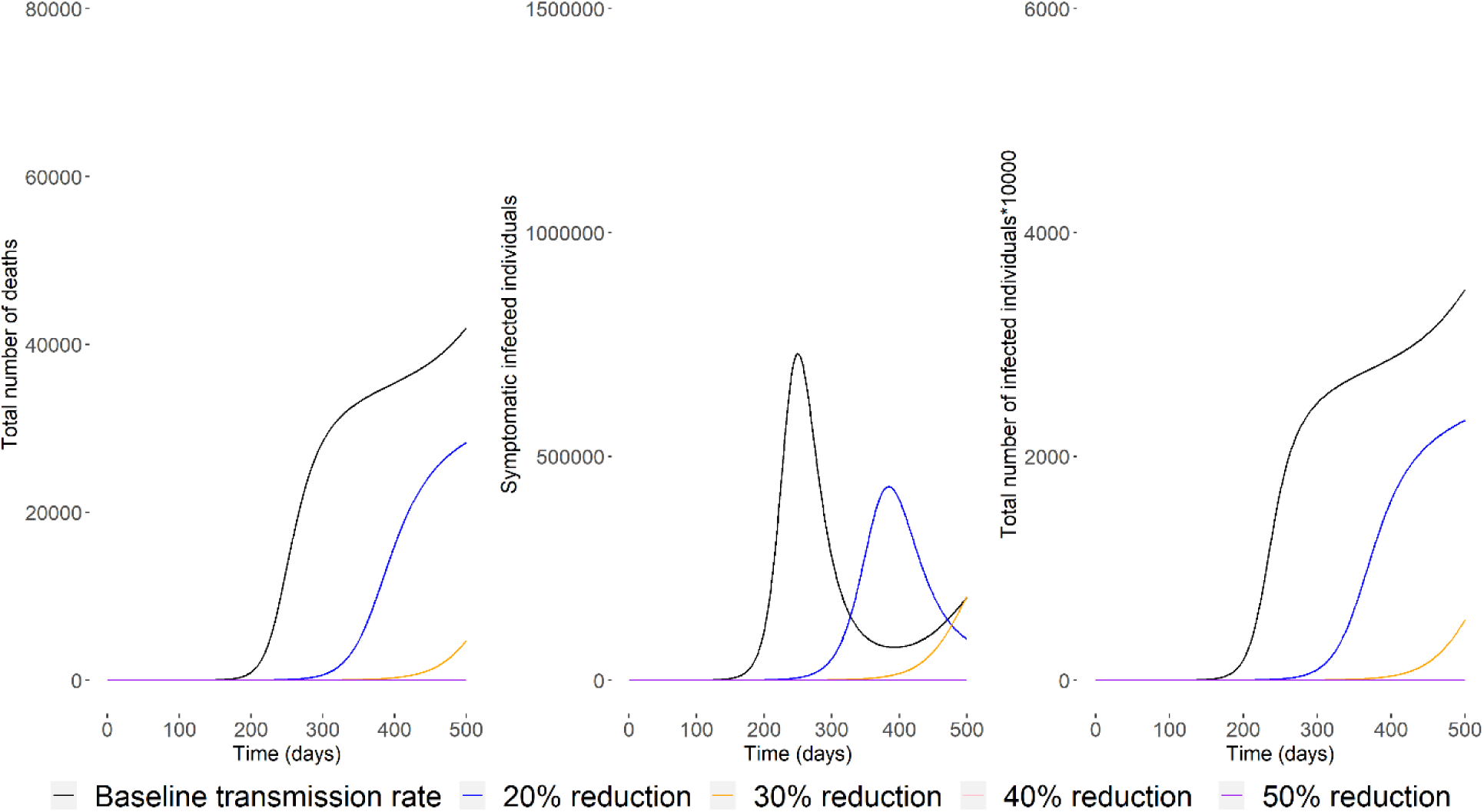
The cumulative number of deaths (left panel), the daily number of (prevalent) symptomatic infectious individuals (middle panel), and the total number of infected individuals (x 10000) (right panel) in Ghana over 500 days under five different scenarios with varying transmission rates: baseline (black), 20% reduction (blue), 30% reduction (orange), 40% reduction (pink), and 50% reduction (purple).

The timing and magnitude of the peak of symptomatic infections were also associated with a decline in transmission rates. For example, at the baseline, <1,000,000 persons were symptomatic, with the peak at almost 250 days. However, a 20% decline in transmission rate suggested a reduction to < 500,000 symptomatic infections, and the timing of the peak was delayed to almost 400 days. The peaks were delayed beyond the simulation timeframe of 500 days for a 30% or more decline in transmission rates (Figure 2).

The total cumulative infections in the baseline transmission rate were almost 40,000,000 by the end of the 500 days. Achieving a 30% decline in transmission rates was associated with less than 20,000,000 cumulative infections in Ghana by the end of the simulation (Figure 2).

### Impact of vaccination rollout on symptomatic infections, cumulative deaths, and cumulative infections

The expected number of deaths from COVID-19 was similar in the scenarios of 0%, 2.8%, and 5.6% vaccination coverage in the first 200 days of the simulation. However, compared with the no vaccination scenario, the vaccination rate at baseline (2.8%) was associated with a decline in cumulative deaths (approximately 40,000) by the end of the simulation. Based on our simulation, increasing the coverage to 20% or more in four months was associated with deaths below 8,000 by the end of the simulation (Figure 3).

**Figure 3:**
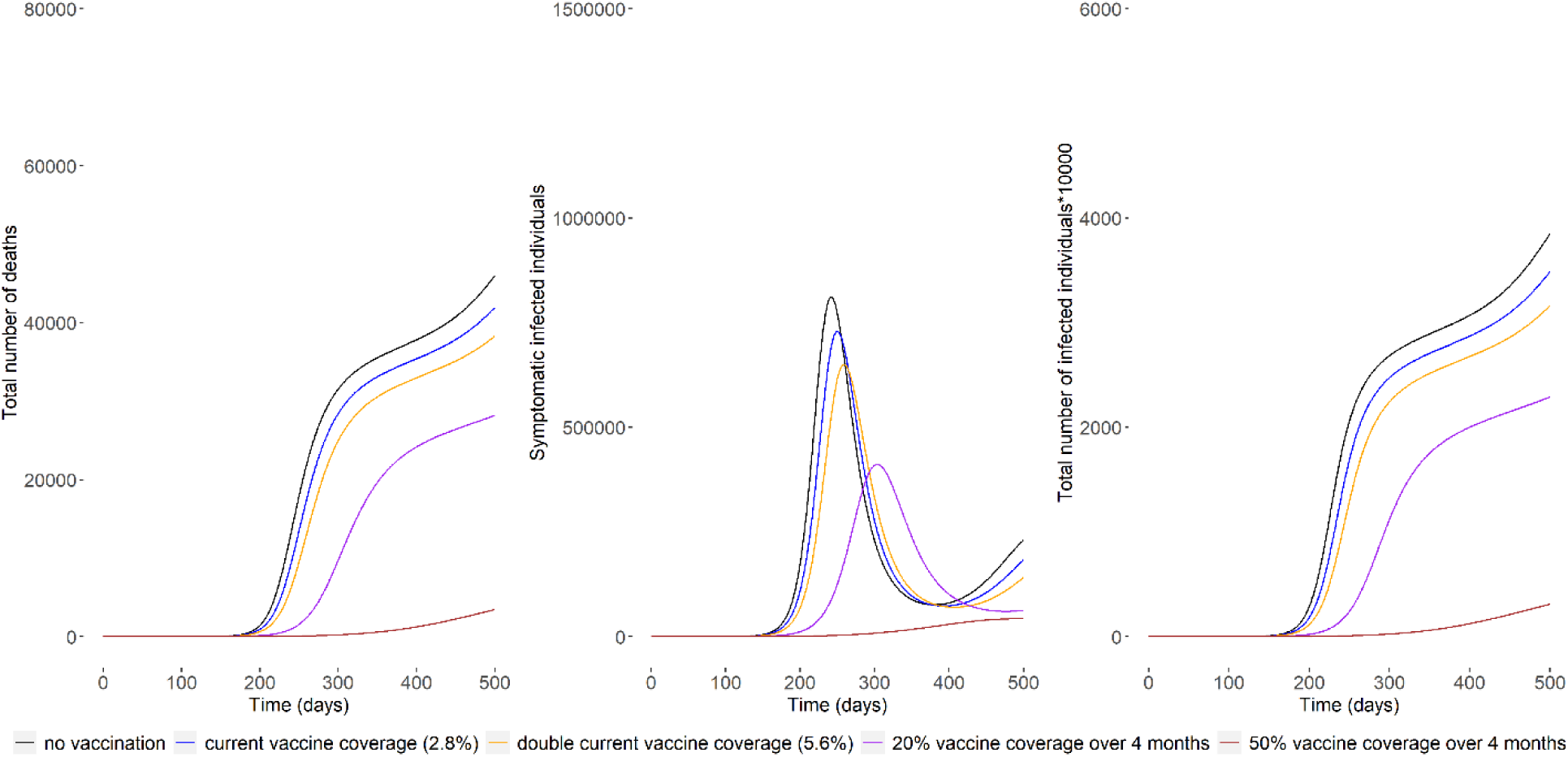
The cumulative number of deaths (left panel), the daily number of (prevalent) symptomatic infectious individuals (middle panel), and the total number of infected individuals (x 10000) (right panel) in Ghana over 500 days under five different vaccination rates scenarios: no vaccination (black), current vaccine coverage (2.8%) (blue), double the current vaccine coverage (5.6%) (orange), 20% vaccine coverage over four months (purple), and 50% vaccine coverage over four months (brown).

A similar trend was observed with the peak of symptomatic and cumulative infections. There was a slight difference in the magnitude and timing of the peak under the first three coverage scenarios (500,000 - 1,000,000 infections). However, achieving 20% vaccine coverage decreased the magnitude to <500,000 infections before the 400th day, while a 50% vaccine coverage successfully delayed and flattened the peak beyond 500 days. The findings in the first three coverage scenarios also showed that a second wave emerged in the second year of the pandemic. The cumulative number of infections due to COVID-19 decreased to less than 30 million when the vaccine coverage increased to 20% or more in the first year (Figure 3).

### Impact of reduction in transmission rates and increasing vaccination rates on the deaths and cumulative infections averted

Figures 4 and 5 showed that increased vaccine coverage and reduced transmission due to adherence to NPIs were associated with an increase in the number of infections and deaths averted.

**Figure 4:**
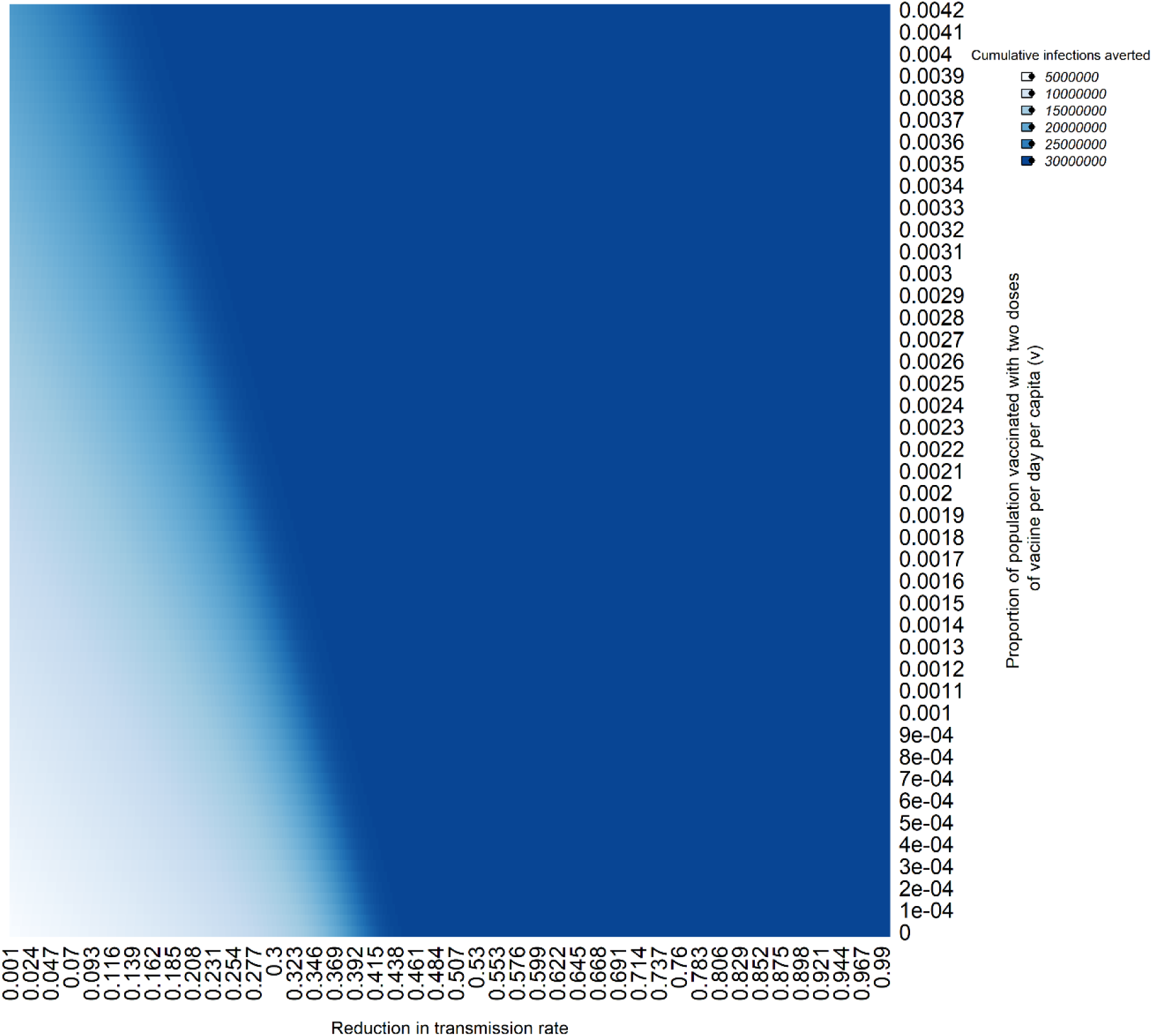
Cumulative infections averted with a reduction in transmission rates and increasing vaccination rates in Ghana over 500 days.

**Figure 5:**
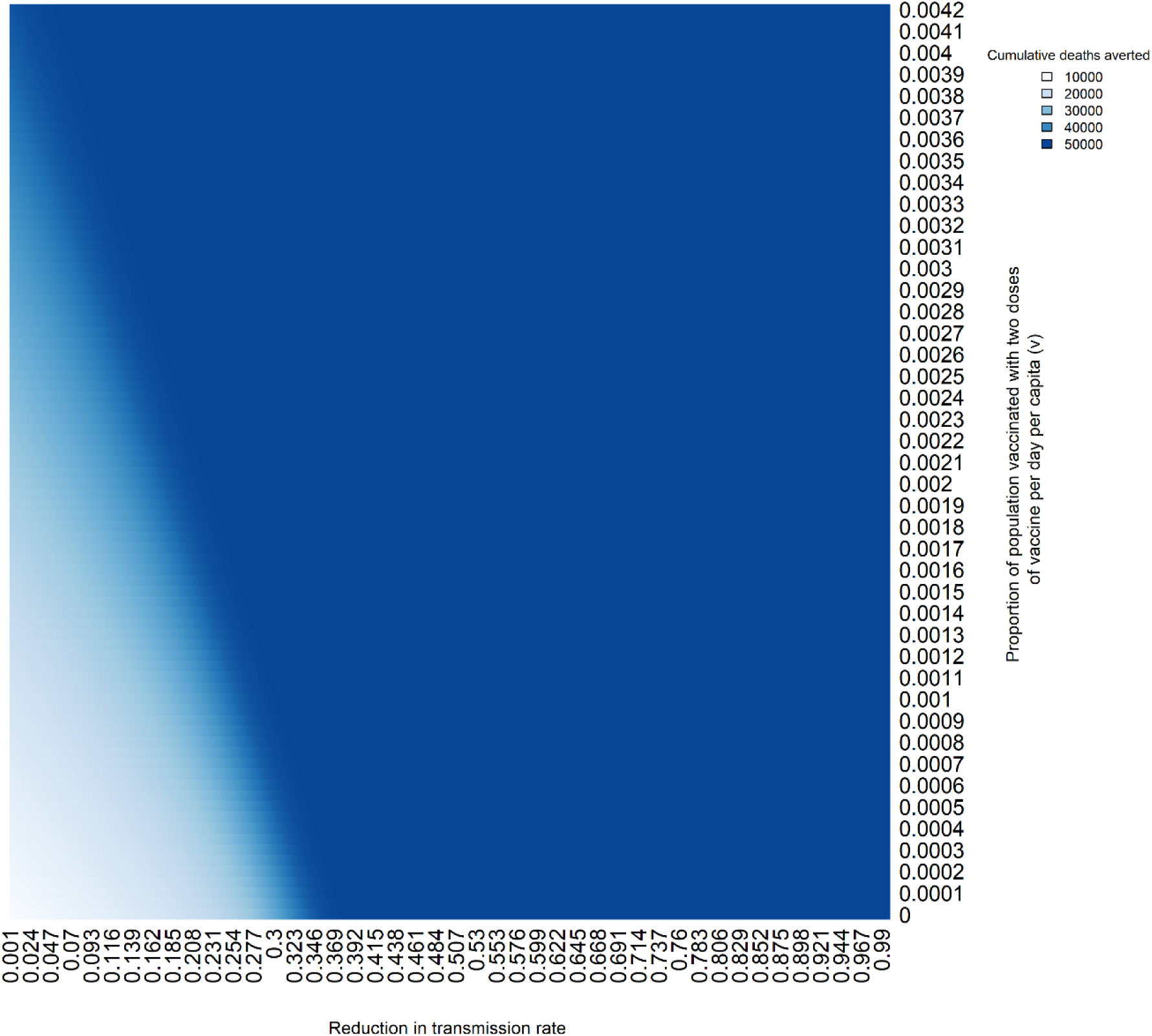
Cumulative deaths averted with a reduction in transmission rates and increasing vaccination rates in Ghana over 500 days.

The results suggested NPIs that declined the transmission rate by at least 35% were associated with more than 25,000,000 cumulative infections averted and >40,000 deaths averted even with a low vaccination rate. The number of infections (5,000,000) and deaths (10,000) averted were the lowest when the vaccination rate was low and the reduction of transmission rates due to NPIs was minimal.

### The uncertainty analysis on the impact of the decline in transmission rates on deaths and infections averted

The results suggested the cumulative infections and deaths averted reached a plateau after about a 50% decline in transmission rates implying that the epidemic has been delayed beyond the simulation timeframe of 500 days (Figure 6). About 50,000,000 million infections and approximately 120,000 deaths were averted at the base case scenario. Per the uncertainty analysis, the plateau was observed after a 50% decline with a median of roughly 50,000,000 infections and 120,000 deaths averted. Our baseline values were slightly lower than the medians beyond a 60% decline in transmission for the deaths averted.

**Figure 6:**
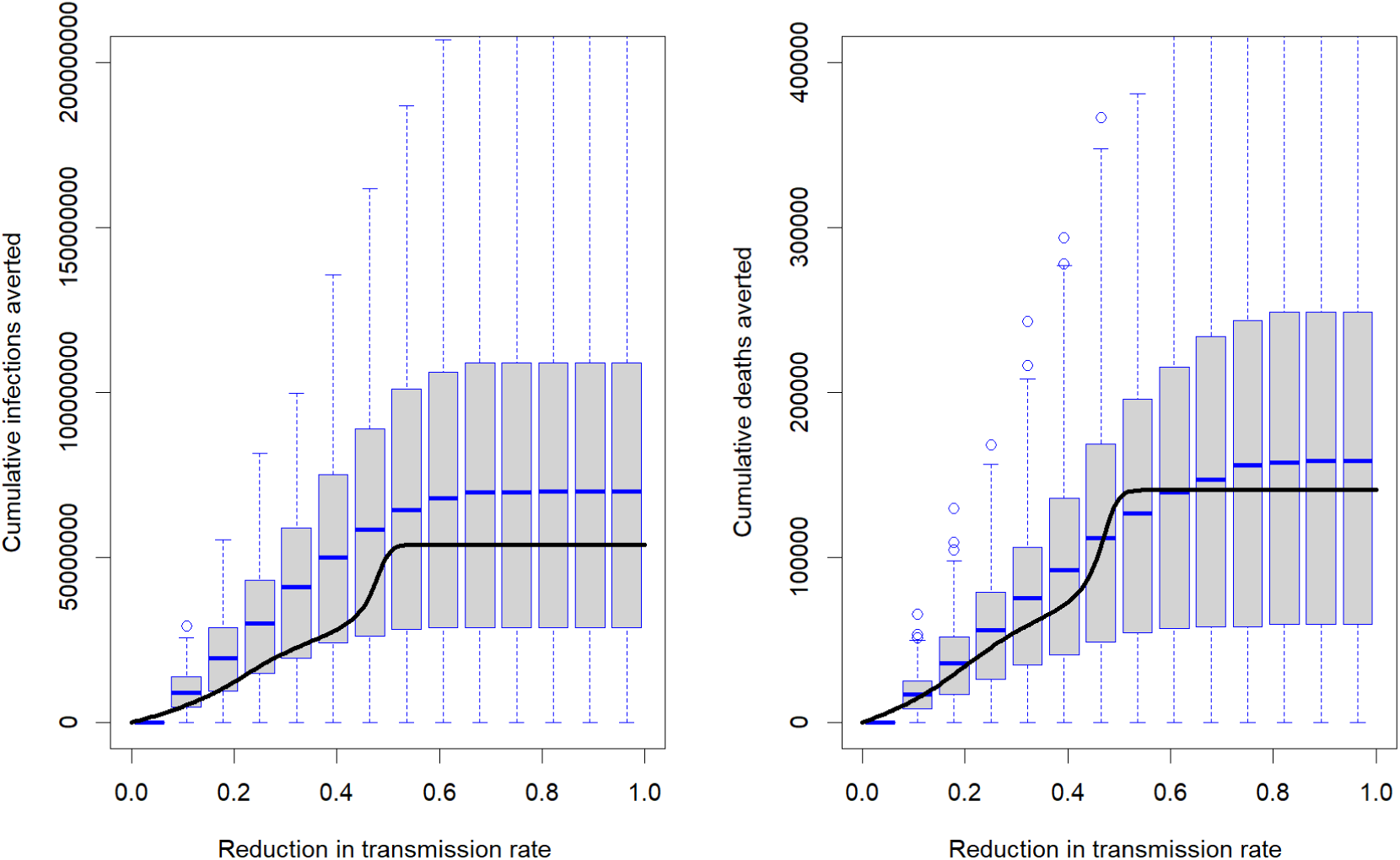
Cumulative infections and deaths averted with varying transmission rates and sensitivity analysis of parameters using Latin hypercube sampling in Ghana over 500 days with a black line showing results for the baseline scenario.

### The uncertainty analysis on the impact of increasing vaccination rates on deaths and infections averted

In the base case scenario, about 25,000,000 infections were prevented when the daily vaccination rate was 0.0042, which is when 50% of the population had received two doses of the vaccine. The uncertainty analysis showed our base-case scenario resulted in values at the 75th percentile. Almost 30,000 deaths were averted when the daily vaccination rate increased to 0.0042. The uncertainty analysis suggests our base-case scenario estimates for the deaths averted were around the median values (Figure 7).

**Figure 7:**
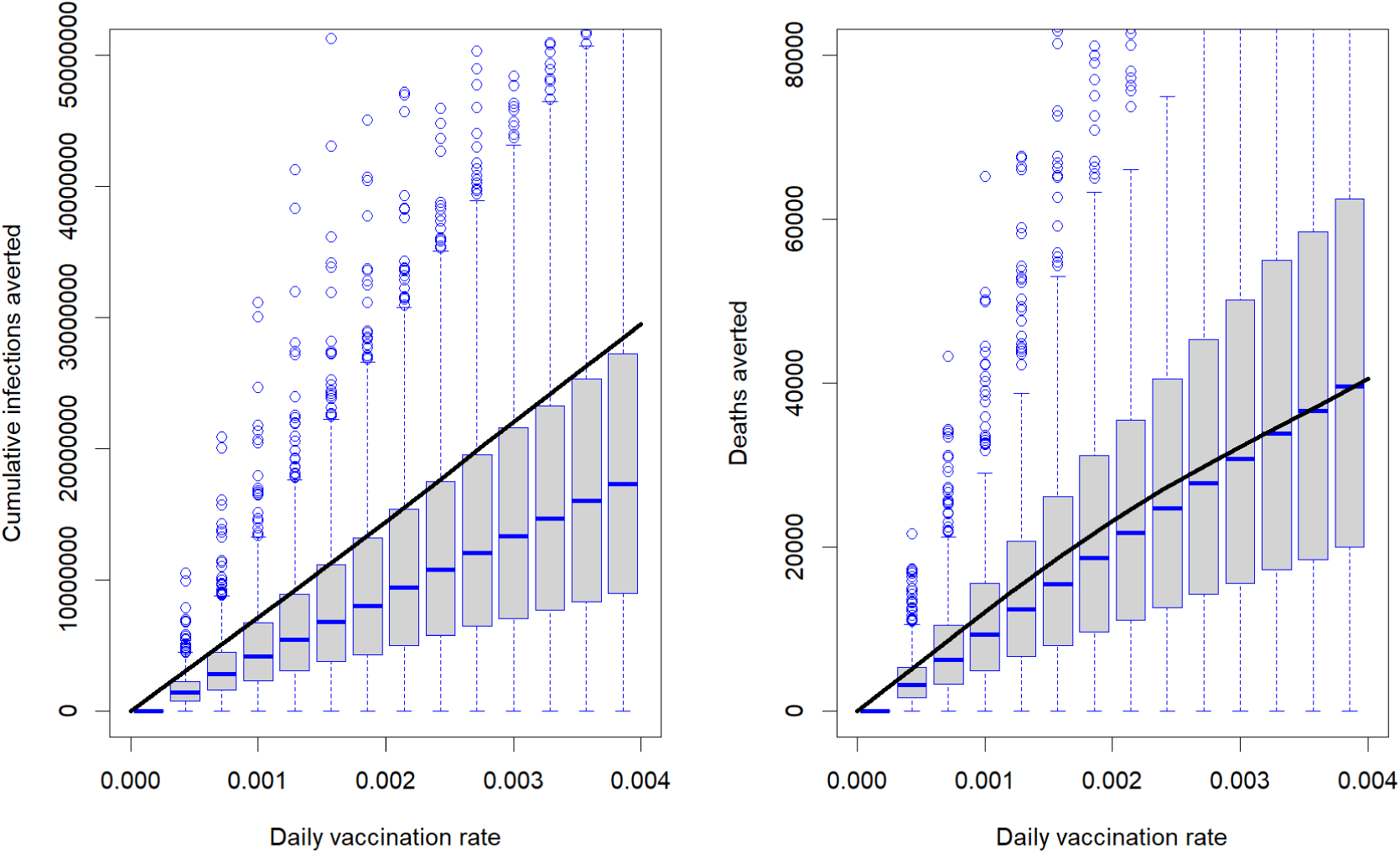
Cumulative infections and deaths averted with increasing daily vaccination rate and sensitivity analysis of parameters using Latin hypercube sampling in Ghana over 500 days with a black line showing results for the baseline scenario.

### Global sensitivity analysis of the impact of parameters on cumulative infections and deaths averted with a decline in transmission rates using partial rank correlation coefficient

We ran a global sensitivity analysis of parameter values on cumulative infections and deaths averted, across scenarios when transmission rates declined (Table 2). The global sensitivity analysis indicated uncertainty in cumulative infections averted were most sensitive to the transmission rate (*β*) (PRCC, 0.598, 95% CI:0.581, 0.614), decline in transmission rates (*l*) (PRCC, 0.636, 95% CI:0.622, 0.650), the rate at which immunity wanes (*w*) (PRCC, 0.383, 95% CI: 0.360, 0.409), rate of recovery for asymptomatic individuals (*q*) (PRCC, -0.302, 95% CI:-0.322, -0.282) and the rate that individuals leave the symptomatic compartment (*f*) (PRCC, -0.249, 95% CI: - 0.279, -0.219). Similar correlations were observed with the deaths averted. The uncertainty in the number death averted was most sensitive to the transmission rate (*β*) (PRCC, 0.570, 95% CI:0.554, 0.590), decline in transmission rate (*l*) (PRCC, 0.656, 95% CI: 0.642, 0.668), the rate at which immunity wanes (*w*) (PRCC, 0.364, 95% CI: 0.342, 0.387), the rate of recovery from asymptomatic infection (*q*) (PRCC, -0.272, 95% CI: -0.293, -0.253), and the rate that individuals leave the symptomatic compartment (*f*) (PRCC, -0.223, 95% CI: -0.252, -0.199) (Table 2).

**Table 2:** Global sensitivity analysis of the impact of parameters on cumulative infections and deaths averted with a decline in transmission rates using Partial rank correlation coefficient.

| Parameter | Symbol | Cumulative infections averted |  | Deaths averted |  |
| --- | --- | --- | --- | --- | --- |
|  |  | PRCC | 95% CI | PRCC | 95% CI |
| Probability of an exposed person becoming pre-symptomatically infected | $\delta$ | 0.056 | 0.03, 0.077 | 0.131 | 0.101, 0.157 |
| Rate at which individuals move from exposure period to infectiousness period | $k$ | 0.009 | -0.016, 0.034 | 0.003 | -0.020, 0.027 |
| Rate at which presymptomatic individuals move from presymptomatic to symptomatic | $c$ | -0.029 | -0.051, -0.007 | -0.018 | -0.046, -0.001 |
| Infection rate | $\beta$ | 0.598 | 0.581, 0.614 | 0.570 | 0.554, 0.590 |
| Rate of recovery for asymptomatic individuals | $q$ | -0.302 | -0.322, -0.282 | -0.272 | -0.293, -0.253 |
| Rate of recovery (or death) for symptomatic individuals (The inverse of the mean symptomatic period) | $f$ | -0.249 | -0.279, -0.219 | -0.223 | -0.250, -0.199 |
| Relative transmissibility of asymptomatic individuals | $u$ | 0.116 | 0.0091, 0.139 | 0.098 | 0.070, 0.125 |
| Relative transmissibility of presymptomatic individuals | $r$ | 0.005 | -0.020, 0.026 | 0.001 | -0.028, 0.020 |
| The rate at which immunity wanes from infection | $w$ | 0.383 | 0.360, 0.409 | 0.364 | 0.342, 0.387 |
| The rate at which immunity wanes for vaccinated individuals | $\chi$ | 0.083 | 0.056, 0.104 | 0.068 | 0.044, 0.091 |
| Vaccine efficacy | $\sigma$ | -0.013 | -0.036, 0.0082 | -0.014 | -0.037, 0.011 |
| Case fatality rate | $z$ | 0.008 | -0.020, 0.031 | 0.102 | 0.083, 0.129 |
| The proportion of the population vaccinated with two doses per day per capita | $\nu$ | -0.099 | -0.122, -0.071 | -0.086 | -0.114, -0.057 |
| Reduction in transmission rate | $l$ | 0.636 | 0.622, 0.650 | 0.656 | 0.642, 0.668 |

CI, confidence intervals; PRCC, partial rank correlation coefficient.

### Global sensitivity analysis of the impact of parameters on cumulative infections and deaths averted with an increase in vaccination rates using partial rank correlation coefficient

We ran a global sensitivity analysis of parameter values on cumulative infections and deaths averted across scenarios when vaccination rates increased (Table 3). The global sensitivity analysis indicated the number of cumulative infections was most sensitive to the proportion of the population vaccinated with two doses per capita per day (*v*) (PRCC, 0.654, 95% CI: 0.737, 0.671) and the rate that immunity wanes for vaccinated individuals (*χ*) (PRCC, -0.342, 95% CI: -0.374, - 0.319). Similar results were observed for the number of deaths averted.

**Table 3:** Global sensitivity analysis of the impact of parameters on cumulative infections and deaths averted with an increase in vaccination rates using Partial rank correlation coefficient.

| Parameter | Symbol | Cumulative infections averted |  | Deaths averted |  |
| --- | --- | --- | --- | --- | --- |
|  |  | PRCC | 95% CI | PRCC | 95% CI |
| Probability of an exposed person becoming pre-symptomatically infected | $\delta$ | 0.012 | -0.010, 0.036 | -0.047 | -0.07, -0.022 |
| Rate at which individuals move from exposure period to infectiousness period | $k$ | -0.063 | -0.091, -0.034 | 0.005 | -0.015, 0.029 |
| Rate at which presymptomatic individuals move from presymptomatic to symptomatic | $c$ | -0.013 | -0.042, 0.012 | -0.042 | -0.068, -0.018 |
| Infection rate | $\beta$ | 0.240 | 0.217, 0.264 | 0.272 | 0.251, 0.300 |
| Rate of recovery for asymptomatic individuals | $q$ | -0.066 | -0.091, -0.04 | -0.059 | -0.083, -0.0376 |
| Rate of recovery (or death) for symptomatic individuals (The inverse of the mean symptomatic period) | $f$ | -0.066 | -0.094, -0.043 | -0.148 | -0.175, -0.122 |
| Relative transmissibility of asymptomatic individuals | $u$ | 0.051 | 0.026, 0.076 | 0.028 | 0.005, 0.057 |
| Relative transmissibility of presymptomatic individuals | $r$ | 0.026 | 0.006, 0.058 | 0.031 | 0.005, 0.057 |
| The rate at which immunity wanes from infection | $w$ | 0.266 | 0.242, 0.289 | 0.328 | 0.305, 0.348 |
| The rate at which immunity wanes for vaccinated individuals | $\chi$ | -0.342 | -0.374, -0.319 | -0.353 | -0.375, -0.330 |
| Vaccine efficacy | $\sigma$ | 0.074 | 0.049, 0.102 | 0.040 | 0.019, 0.067 |
| Case fatality rate | $z$ | 0.084 | 0.064, 0.112 | -0.019 | -0.041, 0.0085 |
| The proportion of the population vaccinated with two doses per day per capita | $\nu$ | 0.654 | 0.637, 0.671 | 0.654 | 0.636, 0.674 |
| Reduction in transmission rate | $l$ | -0.012 | -0.042, 0.002 | -0.012 | -0.046, 0.005 |

CI, confidence intervals; PRCC, partial rank correlation coefficient.

## Discussion

A compartmental model was developed to simulate the transmission of COVID-19 in Ghana. This study explored the impact of vaccination coverages and the decline in transmission of COVID-19 under various scenarios. Scenarios with no vaccine coverage performed worse than all vaccination scenarios for our assessed outcomes. The results demonstrated that increasing the vaccination rate to achieve 50% coverage in Ghana could avert approximately 30,000 deaths and 25,000,000 infections in a 500-day time frame. A 50% decline in transmission was associated with 50,000,000 million infections and about 120,000 deaths averted. Reduction in transmission rates remained a vital tool to significantly decline symptomatic and cumulative infections and deaths in the population.

A modeling study of the effect of NPIs in mitigating the COVID-19 pandemic in Kenya reported a 20% decline in transmission rate from the unmitigated scenario was associated with a reduction of cumulative number of infections from about 46 million to about 44 million within a 1-year time frame.^48^ They also found that a decline in transmission by 60% led to a reduction in symptomatic infections at the peak from about 3 million in the unmitigated scenario to a value of <500,000 while the cumulative number of infections decreases from 46 million to about 35 million within a 1-year time frame. The values reported in their study were slightly higher than those found in this study, which may be due to differences in model specification and parameterization, as well as population size differences between Kenya and Ghana. Another study by Bhowmik and colleagues in the United States concluded NPIs leading to a 50% stay-at-home rate were associated with a 33% reduction in cases.^49^ The effect of NPIs on deaths averted have also been reported in other countries. A study by Flaxman and colleagues suggested about 3 million deaths could be prevented across 11 countries in the presence of NPIs.^50^

Vaccination remains a relevant measure in reducing the burden of COVID-19 due to the high level of susceptible persons in the population.^7^ Even with low vaccination rates, a slight reduction in symptomatic infections, cumulative infections, and deaths were observed over time. A 50% coverage successfully flattened the curve, and the peak of the wave was successfully delayed. A 40% coverage in Lebanon led to only a 10% decline in symptomatic infections.^51^ Moghadas and colleagues reported that vaccination reduced the attack rate from 9% to 4.6% in the United States.^52^ Therefore, it is imperative to increase vaccine supplies through sustained efforts from donors and policymakers to ensure a higher coverage is reached over time. Our findings demonstrate the intertwined effectiveness of both vaccination and NPI, as fewer deaths and cumulative infections were observed with the combination of the two measures. Consistent with the conclusion by Moghadas et al., compliance with NPIs remains relevant in the control and mitigation of COVID-19, especially in regions with limited vaccine supplies and slow vaccine rollout.^52^ In Italy, it was reported that fast vaccination rollout was associated with a reduction in deaths from 298,000 deaths to 51,000, even with weak NPIs.^53^

The global sensitivity analysis indicated that the estimates of the effect of increasing vaccination on cumulative infections and deaths averted were most sensitive to the rate at which immunity wanes from infection and vaccination, transmission rates, and vaccination rates. Another study in Senegal supported these finding.^54^ In contrast to their study, cumulative infections and deaths averted were robust to uncertainty in vaccine efficacies. Another study in Pakistan reported that the basic reproductive number of COVID-19 is sensitive to the parameter value of the relative transmissibility of asymptomatically infected individuals, whereas in this study cumulative infections averted and deaths averted are minimally sensitive to the relative transmissibility of asymptomatic individuals. ^55^

The study is not without limitations. First, we assumed the probability of infections was independent of the population contact patterns, assuming it was the same for everyone. We did not consider the availability and use of other COVID-19 vaccines. However, the AstraZeneca COVID-19 vaccine is the most used in Ghana. The analysis also assumed a single wave outbreak and did not account for other variants against which existing vaccines might have different efficacy values. Furthermore, we assumed that vaccination prevented infection. Future studies should consider incorporating more factors into the model.^56^ Moreover, the model assumed individuals who recovered from COVID-19 had a symptomatic period of the same length as those who died. The model did not assess the impact of specific NPIs (e.g., face masks, social distancing, school closures, and lockdowns) but assumed the decline in the transmission coefficient represented their collective impact.

In conclusion, the study’s findings indicate the need to increase vaccination coverage by ensuring increased supply, access, faster rollout, and a coordinated public health effort to scale-up vaccine uptake. The results also demonstrate the need to comply with NPIs and vaccination to ensure COVID-19 is mitigated in Ghana, especially with limited resources.

## Data Availability

This is an epidemiological modeling study. We have provided the R source code such that the study can be reproduced by others.

## Supplementary Materials

First, we load packages that are required for the R simulation.

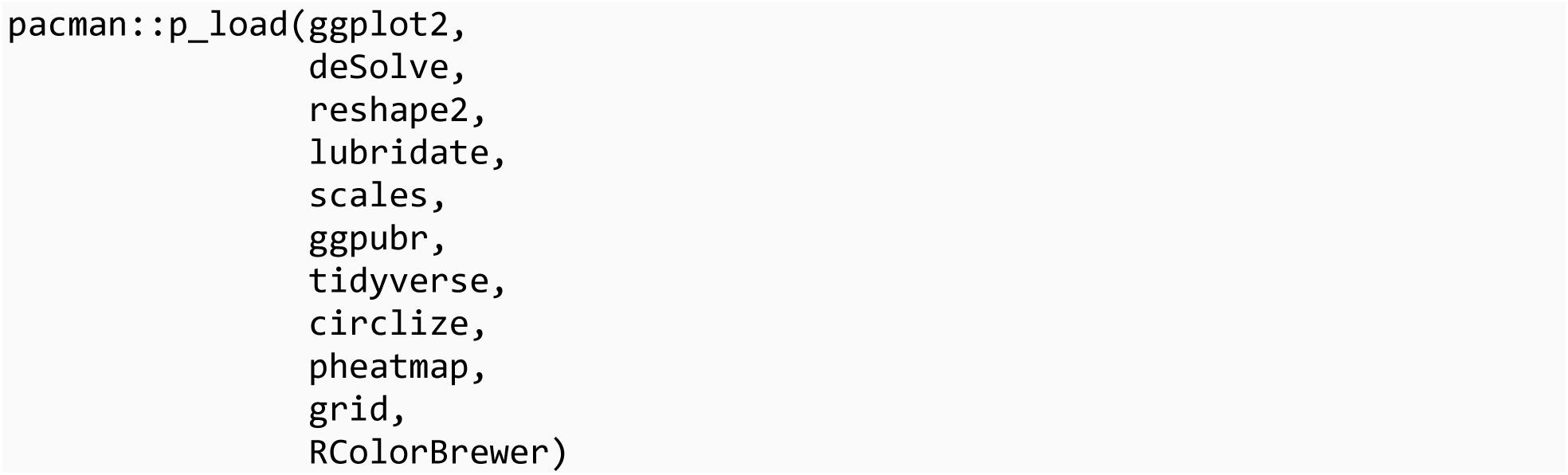

We set a seed so that the same simulation outputs can be obtained when simulations are repeated.

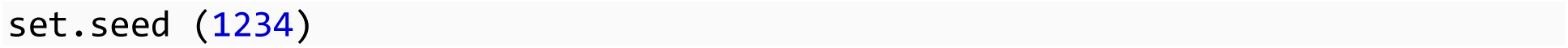

## SEAPIRD-V model

• S refers to susceptible individuals.
• E refers to exposed individuals who are in the latent state.
• A refers to asymptomatic individuals: A percentage of the infected population makes transitions into the asymptomatic state. They are infectious.
• P refers to pre-symptomatic individuals: A percentage of the infected population makes transitions into the pre-symptomatic state. They are infectious.
• I refers to symptomatic individuals. They are infectious.
• R refers to recovered: Asymptomatic and Symptomatic individuals enter the recovery state.
• D refers to symptomatic individuals who died from COVID-19.
• V refers to individuals who become immune from COVID-19 infection after receiving two doses of the AstraZeneca vaccine.

### Parameter definitions

Parameters:

• c: transition rate from presymptomatic to symptomatic= 1/(incubation period - latency period)
• beta: transmission coefficient
• q: recovery rate for asymptomatic individuals
• delta: probability that exposed persons become presymptomatically infected
• f: the inverse of the duration of individuals being symptomatic (equals to the recovery rate for symptomatic individuals)
• u: relative transmissibility of asymptomatic individuals
• r: relative transmissibility of pre-symptomatic individuals
• sigma: vaccine efficacy
• v: vaccination rate
• chi: waning immunity of vaccinated individuals
• Ro: reproduction number
• l: the proportion of decline in transmission (between 0 and 1)
• z: case fatality ratio (between 0 and 1)

First we estimate beta (transmission rate) by simulating values for parameters used in the formula for calculating beta. The definitions for parameters explained above.

Values are assigned to the parameters for their uncertainties which will be used later in the analysis.

### Assigning parameter values

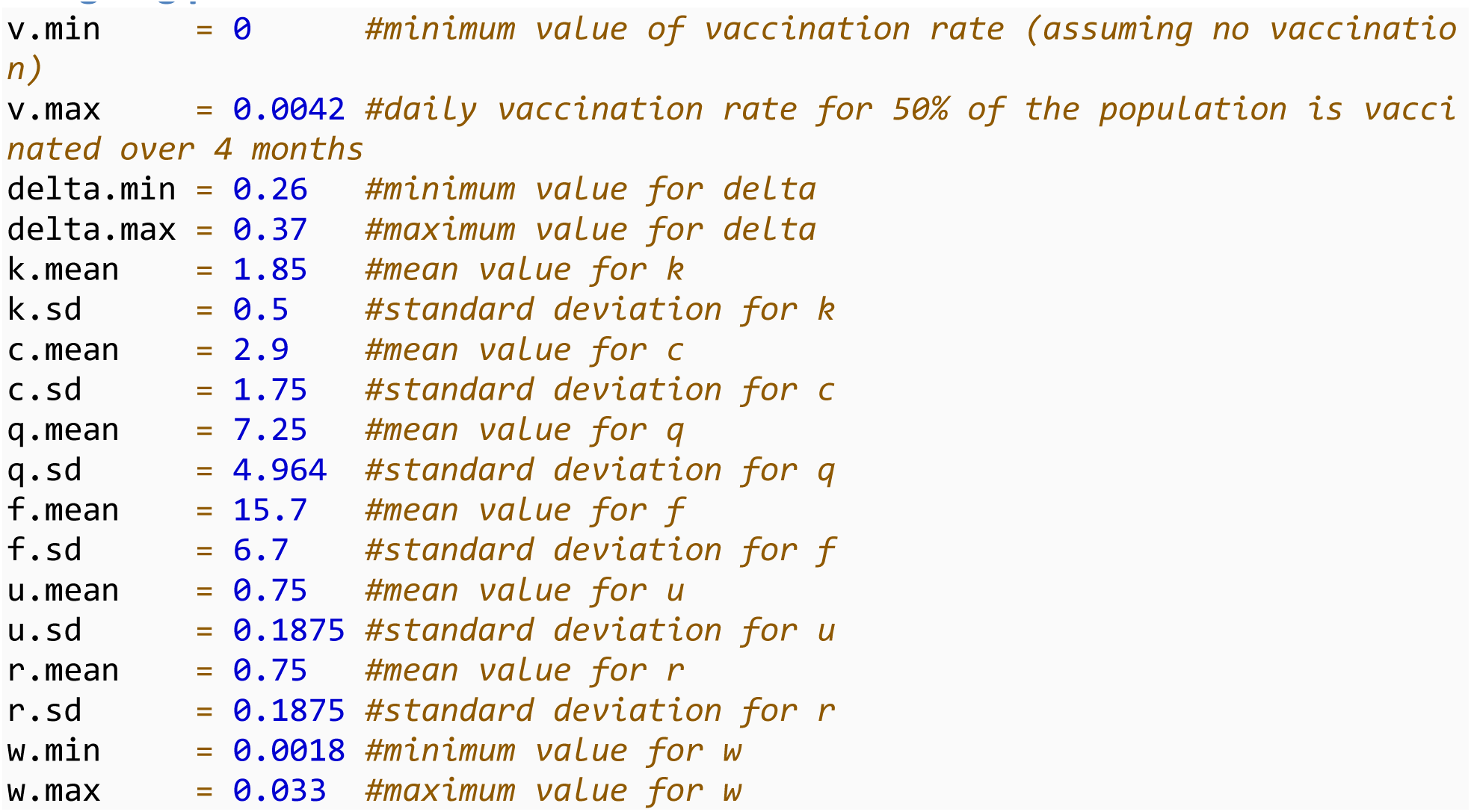
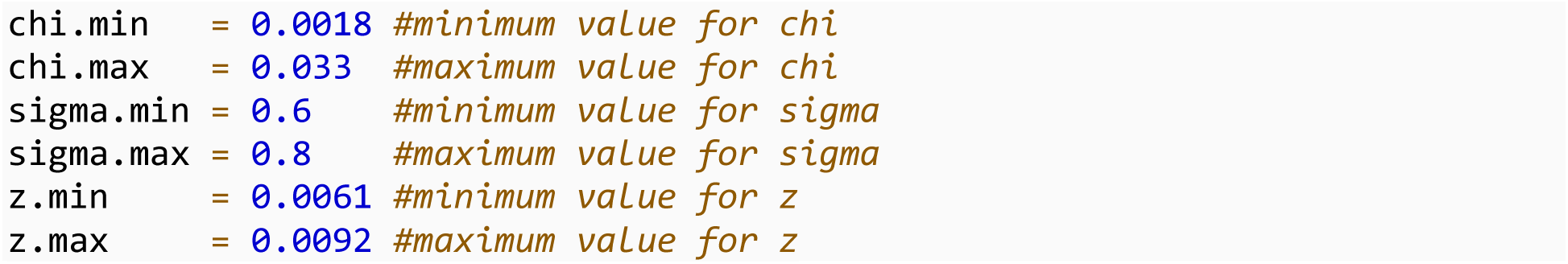

### Calculating the transmission rate

First a function was created for the formula for calculating beta.

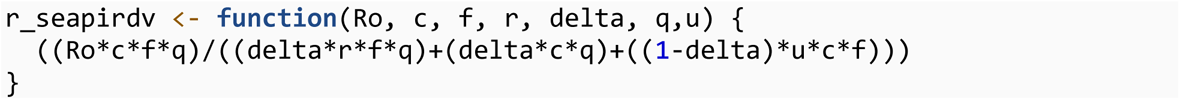

Set the number of simulations

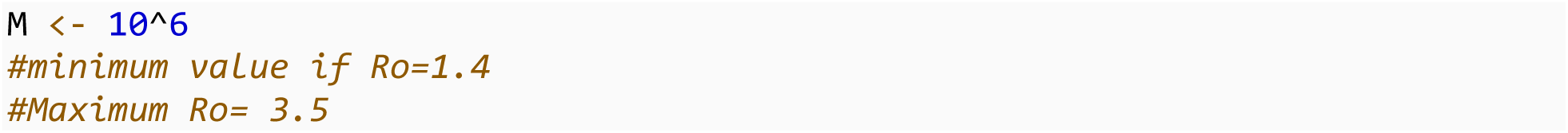

Draw random numbers from the distributions of these variables

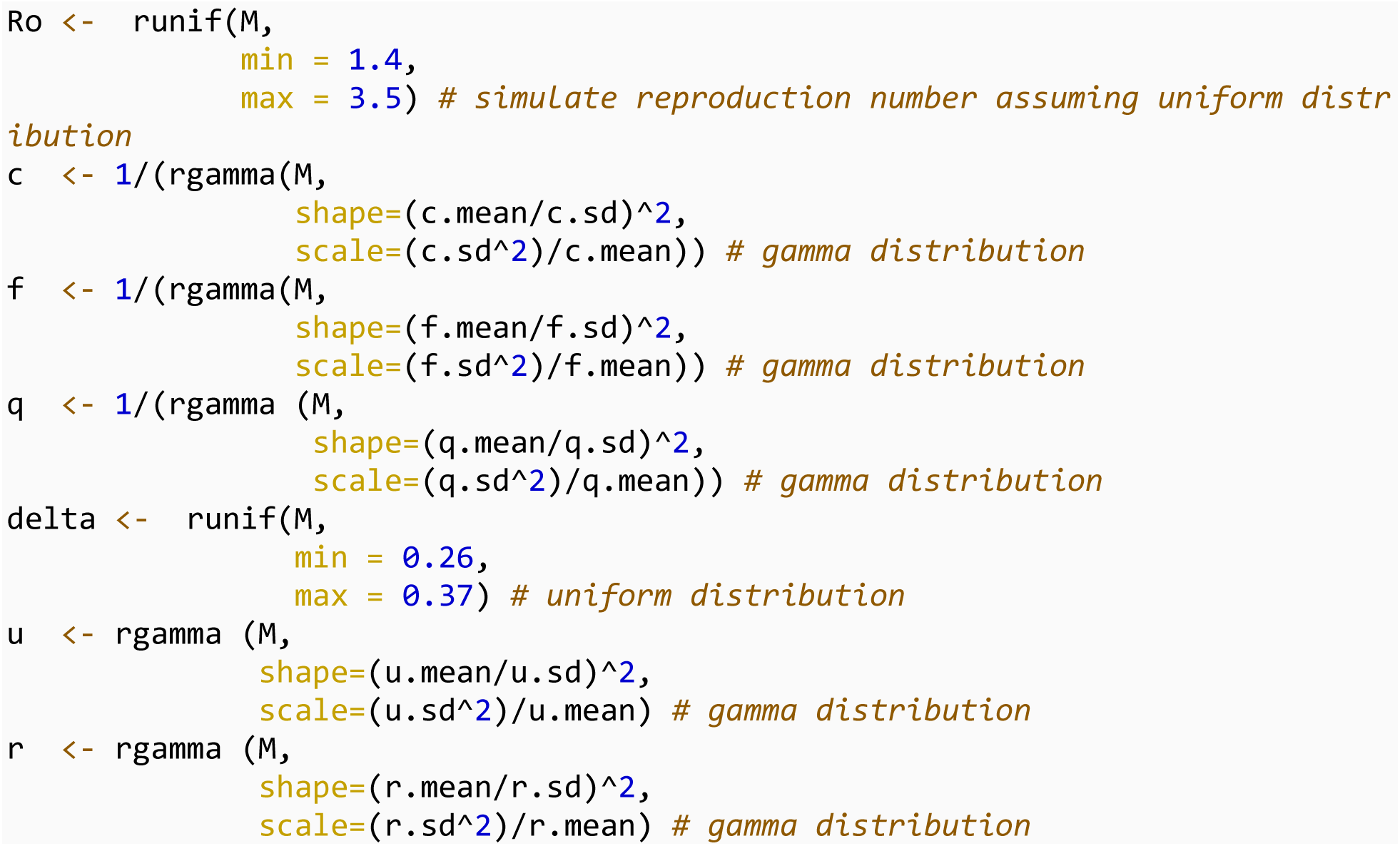

Next we calculate the beta using the function we created:

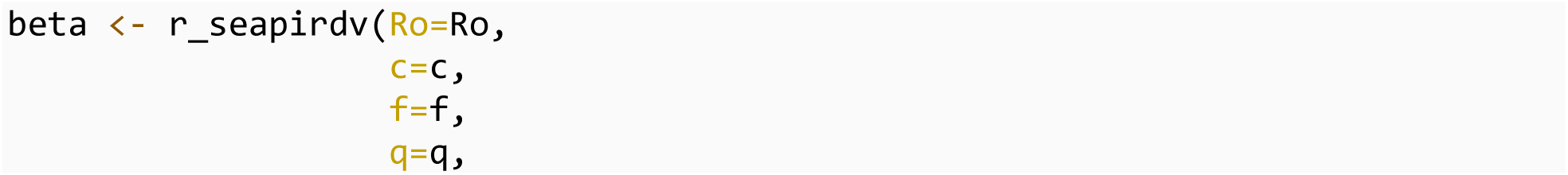
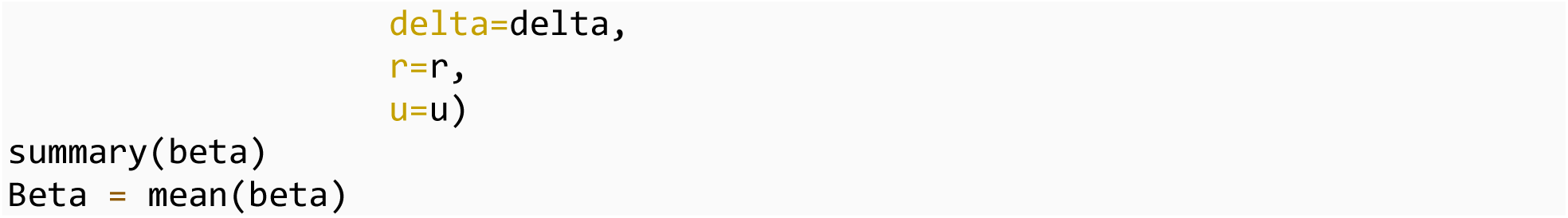

### Setting initial state for compartments

We assumed 19% of the population was already infected, hence they were in the recovered compartment at the beginning of the simulation.

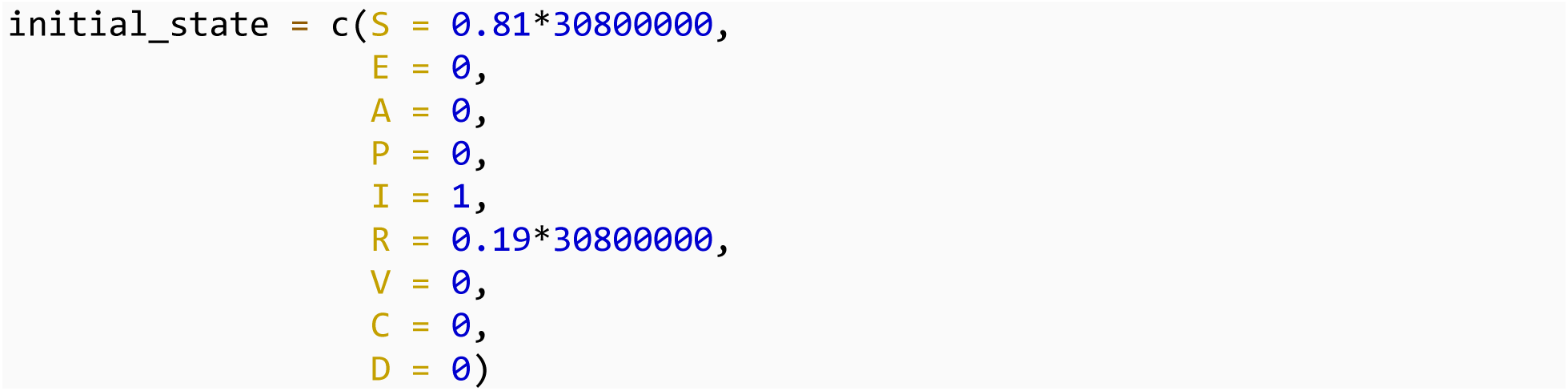

The model was simulated for 500 days to allow enough time for the second wave to emerge.

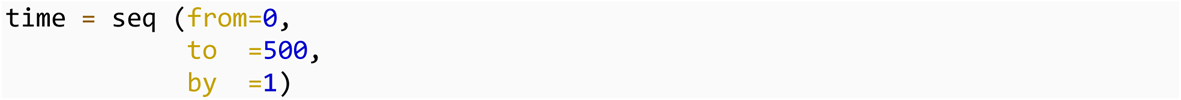

### Solve the equations

Next we created a function with the set of ordinary differential equations that defines the model.

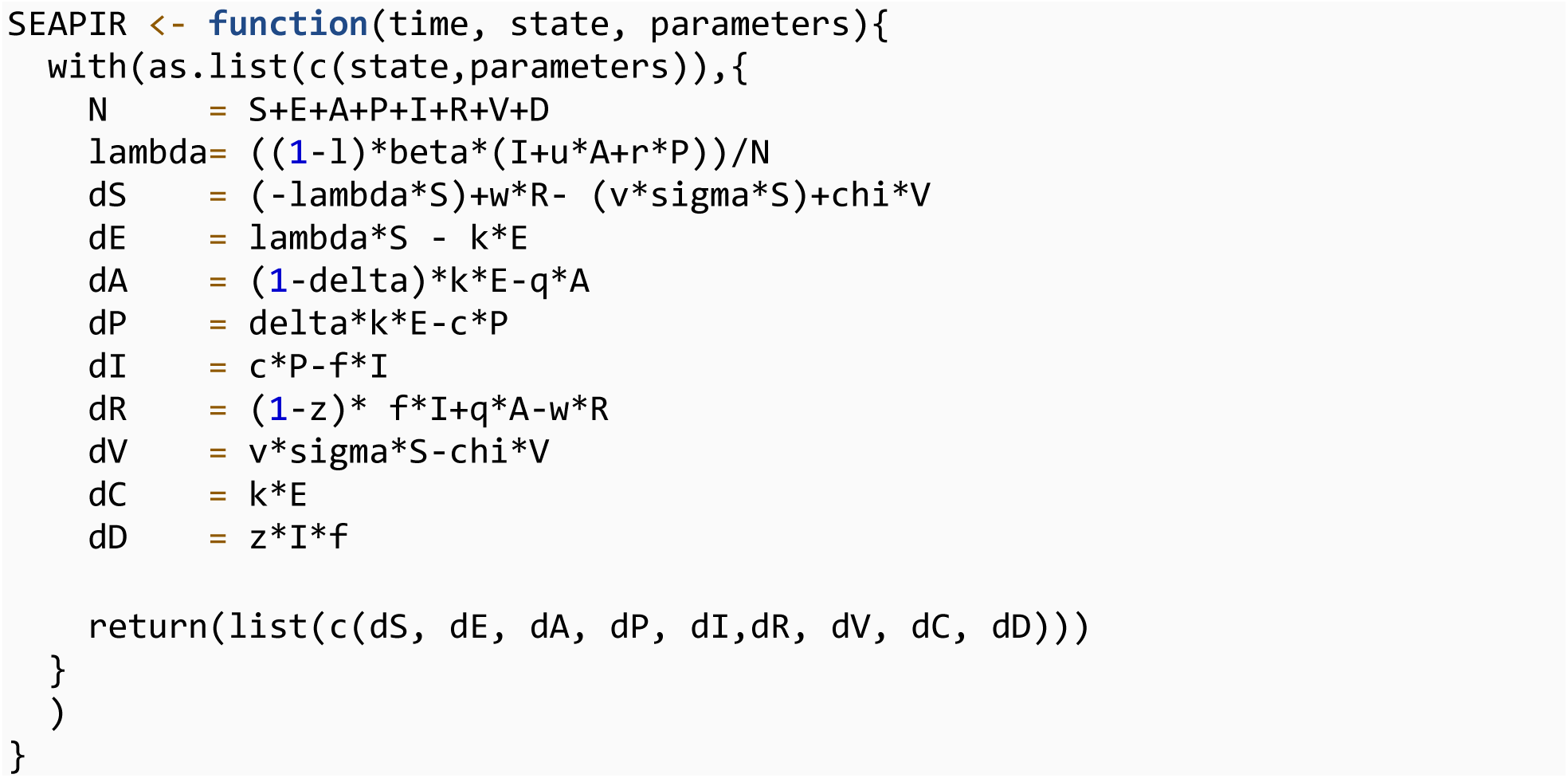

We created a vector for the parameters.

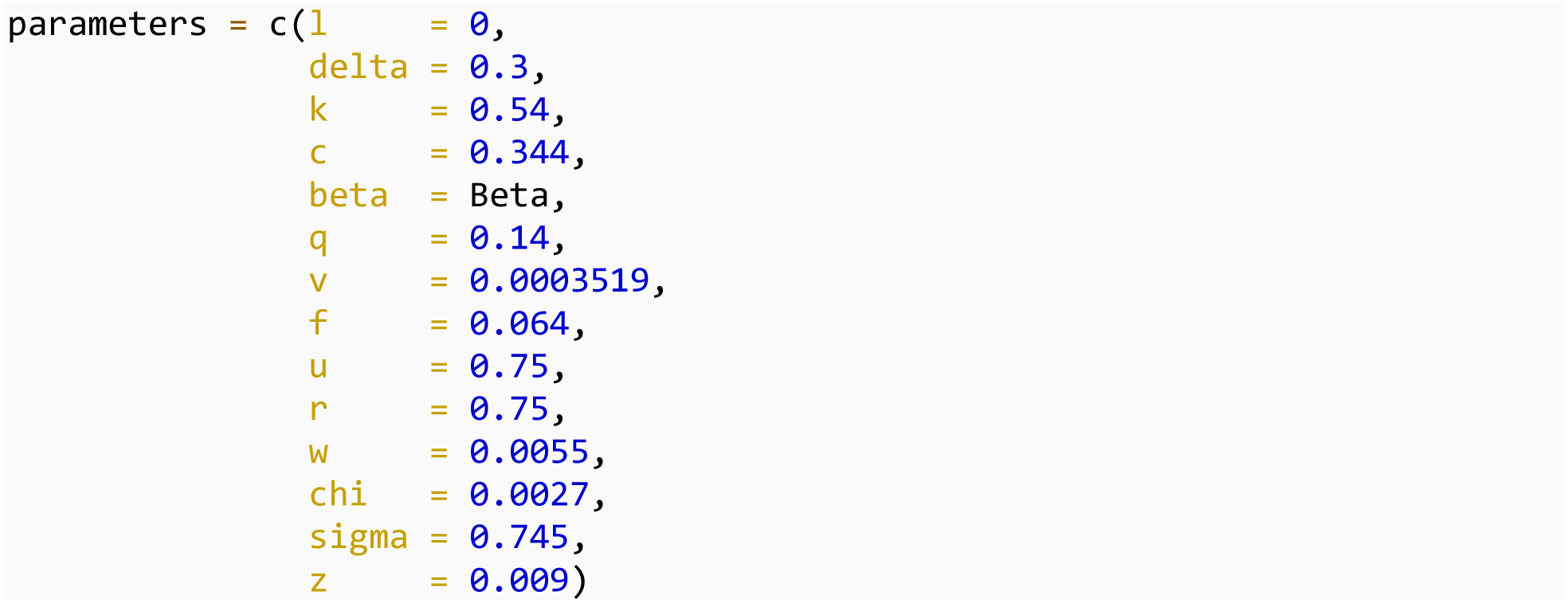

We solved the differential equations using the ode() function in the deSolve package:

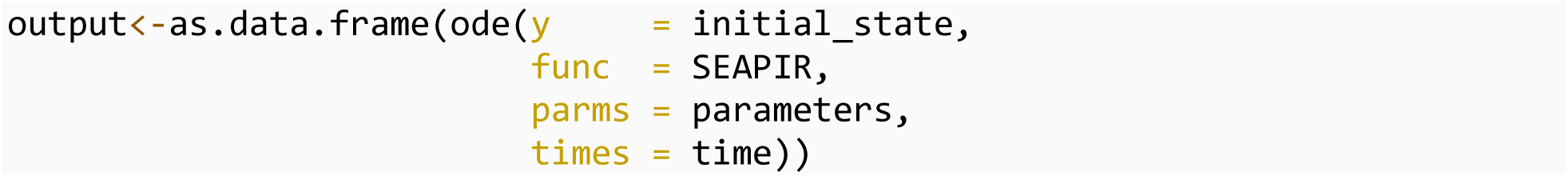

We converted the data into a long format using the melt() function in the reshape2 package:

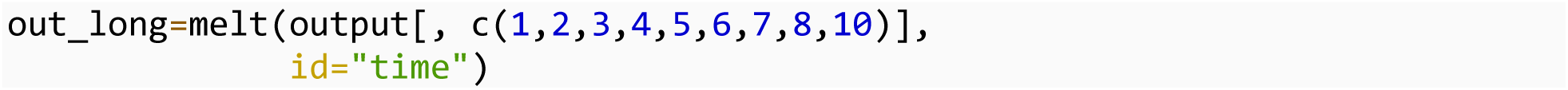

### Plot simulation results

We plotted our simulation results

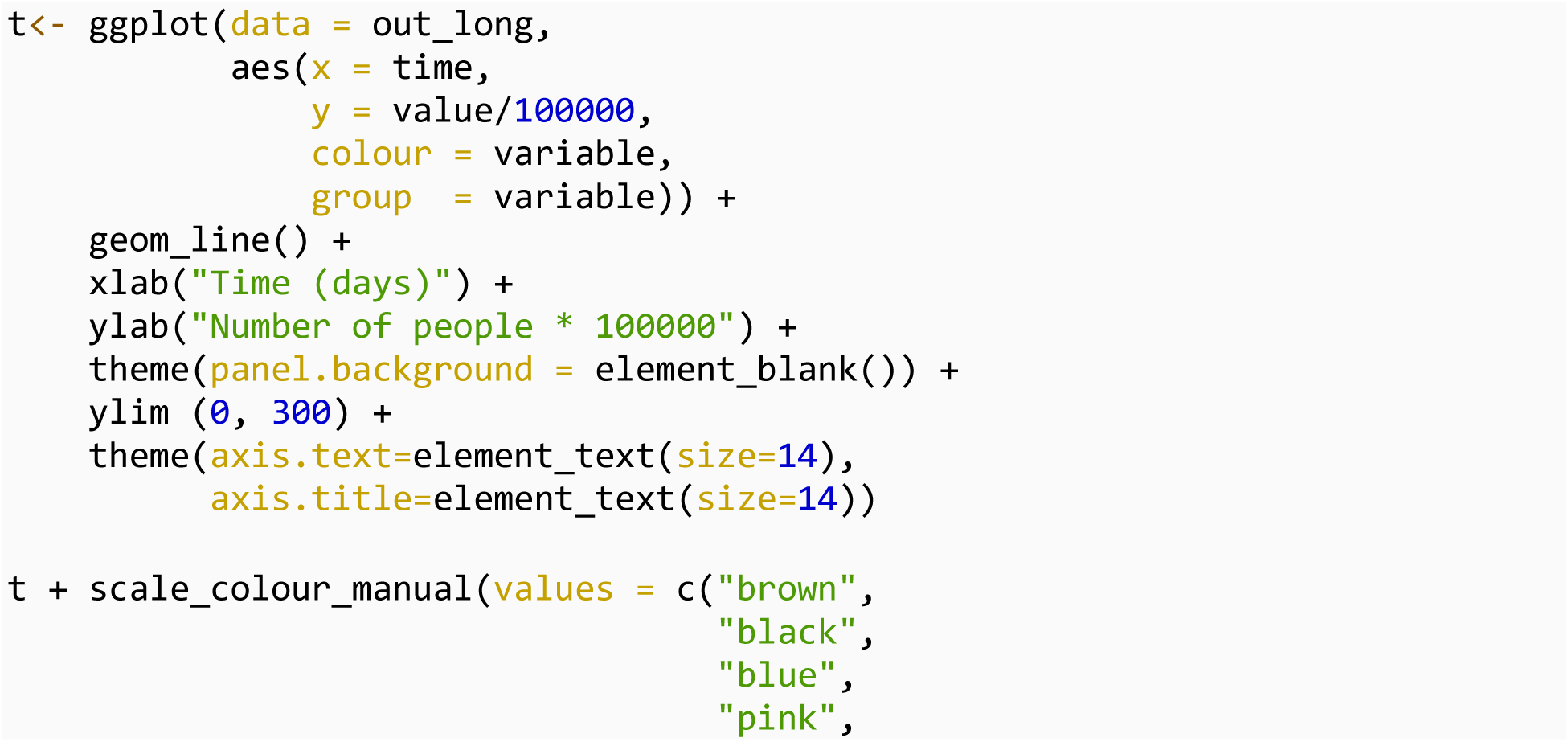
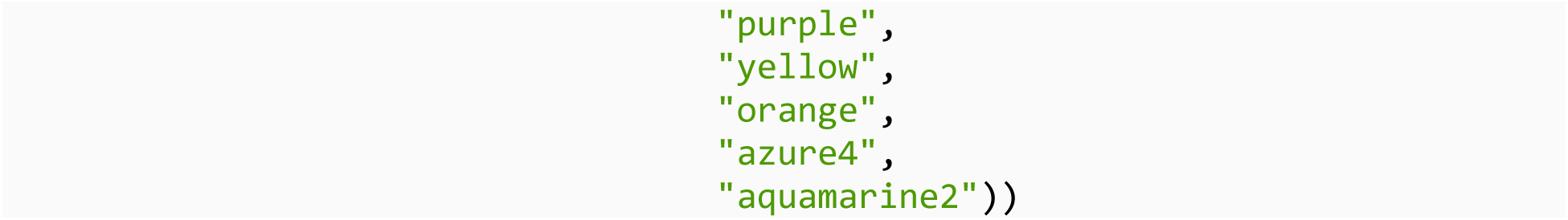

## Imact of declining transmission

Next, we explored the impact of declining transmission on cumulative infections and deaths

### First scenario: l=0

There is no change in transmission rate

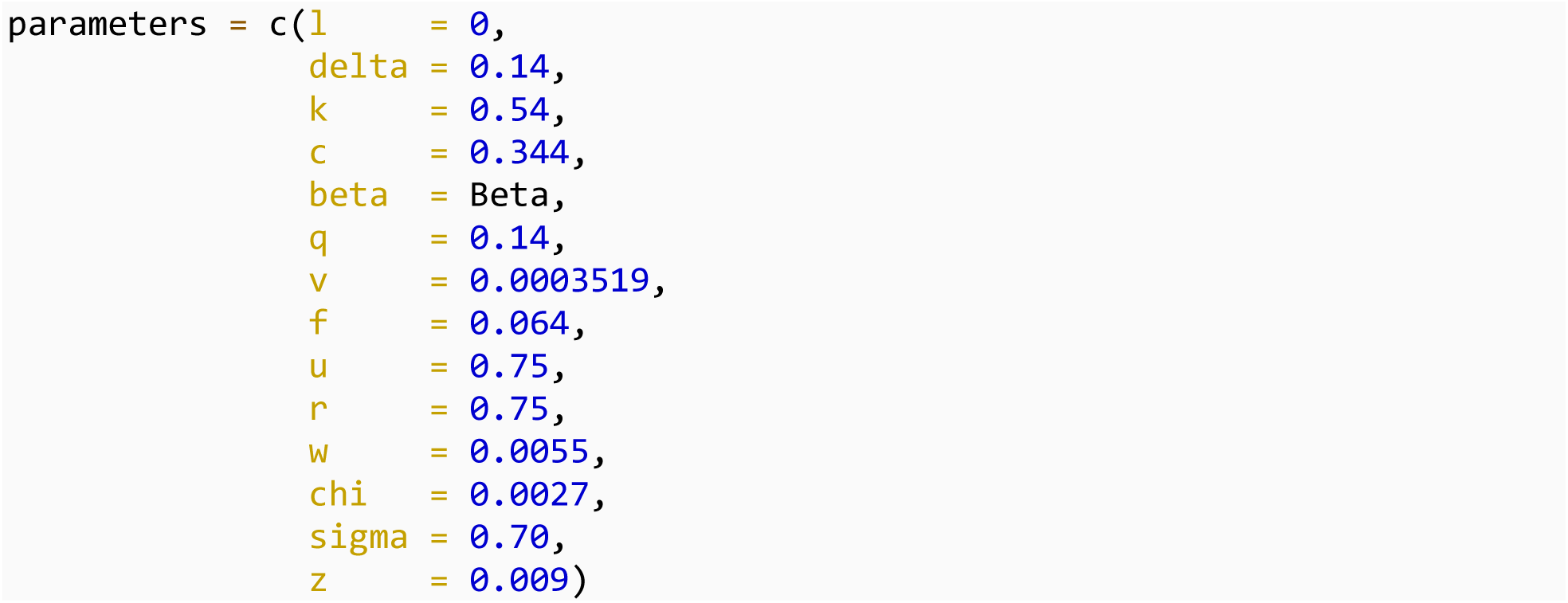

We solved the ordinary differential equations using the ode() function in the deSolve package:

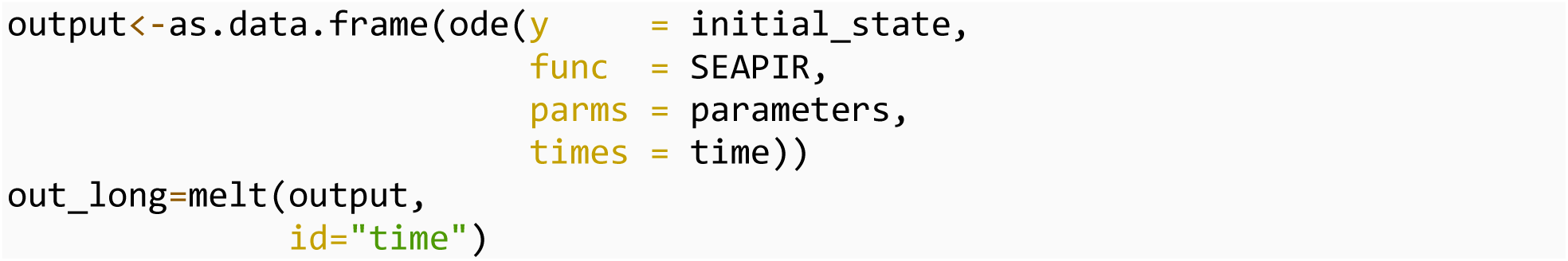

### Second scenario: l=0.2

We assumed a 20% decline in transmission rate

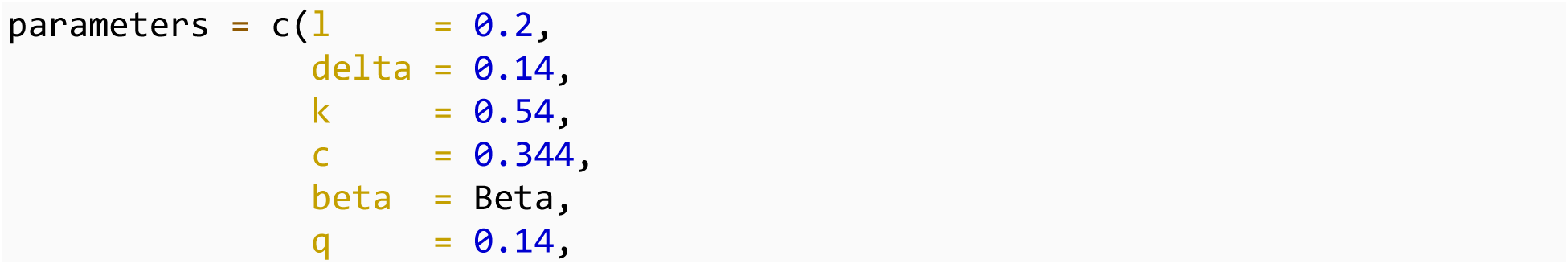
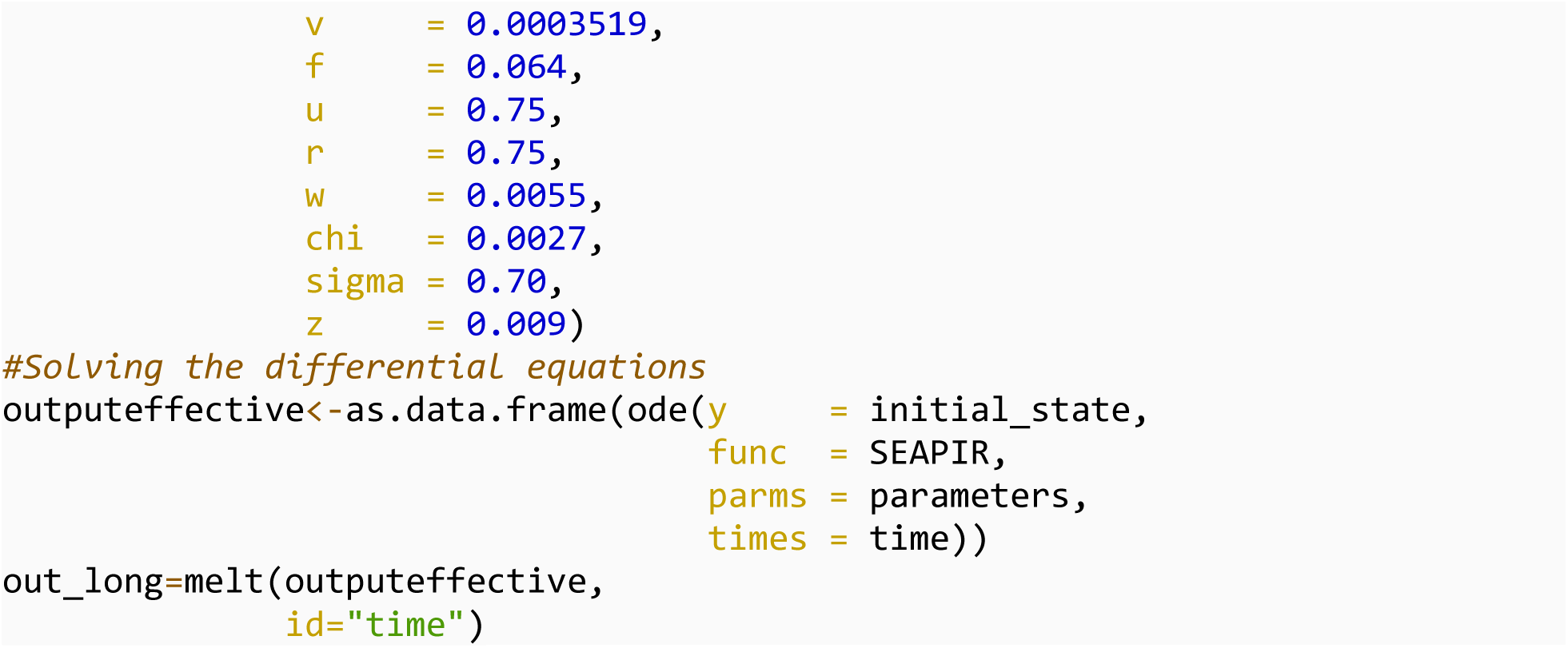

### Third scenario: l=0.3

We assumed a 30% decline in transmission

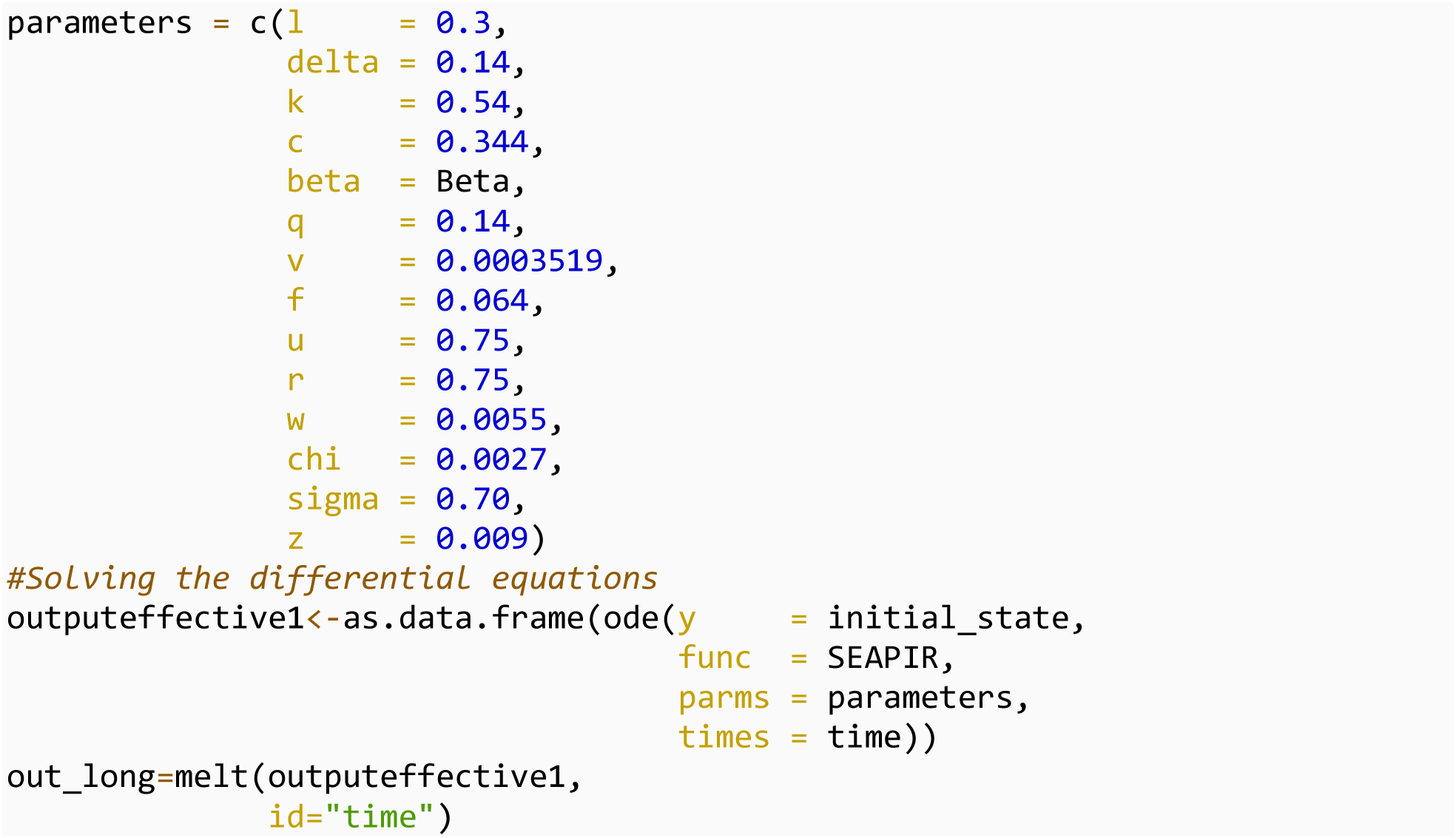

### Fourth scenario: l=0.4

We assumed a 40% decline in transmission

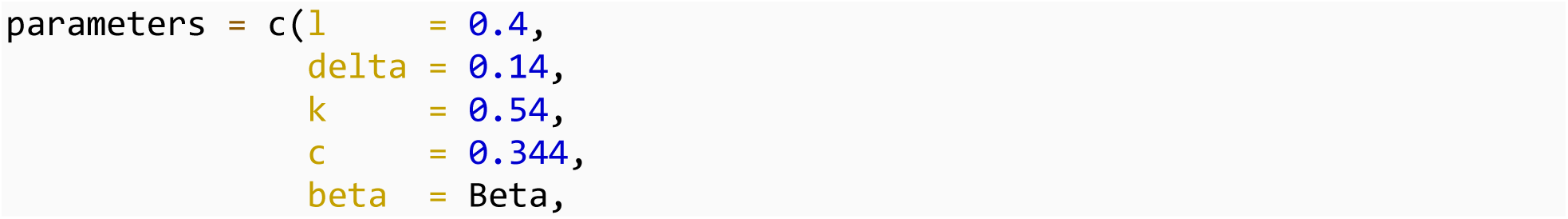
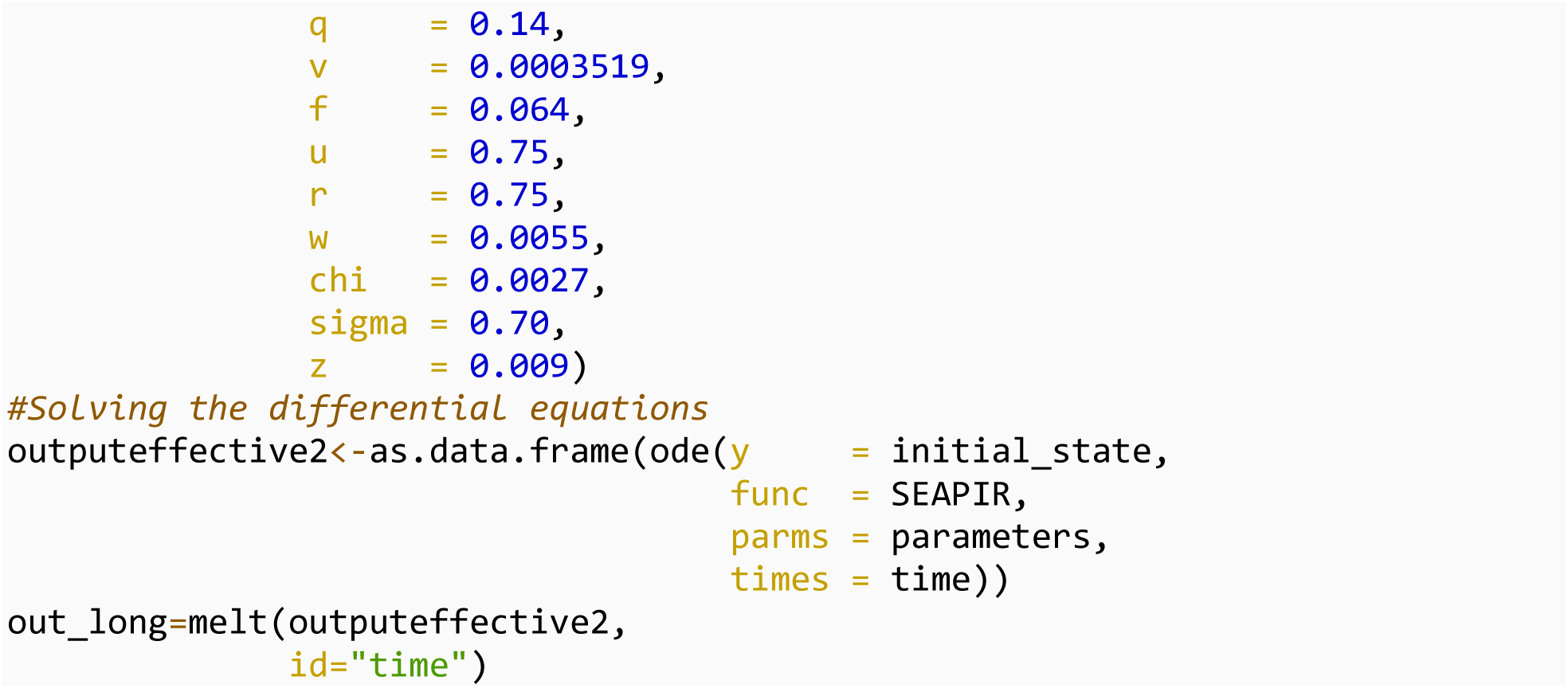

### Fifth scenario assuming a 50% decline in transmission l=0.5

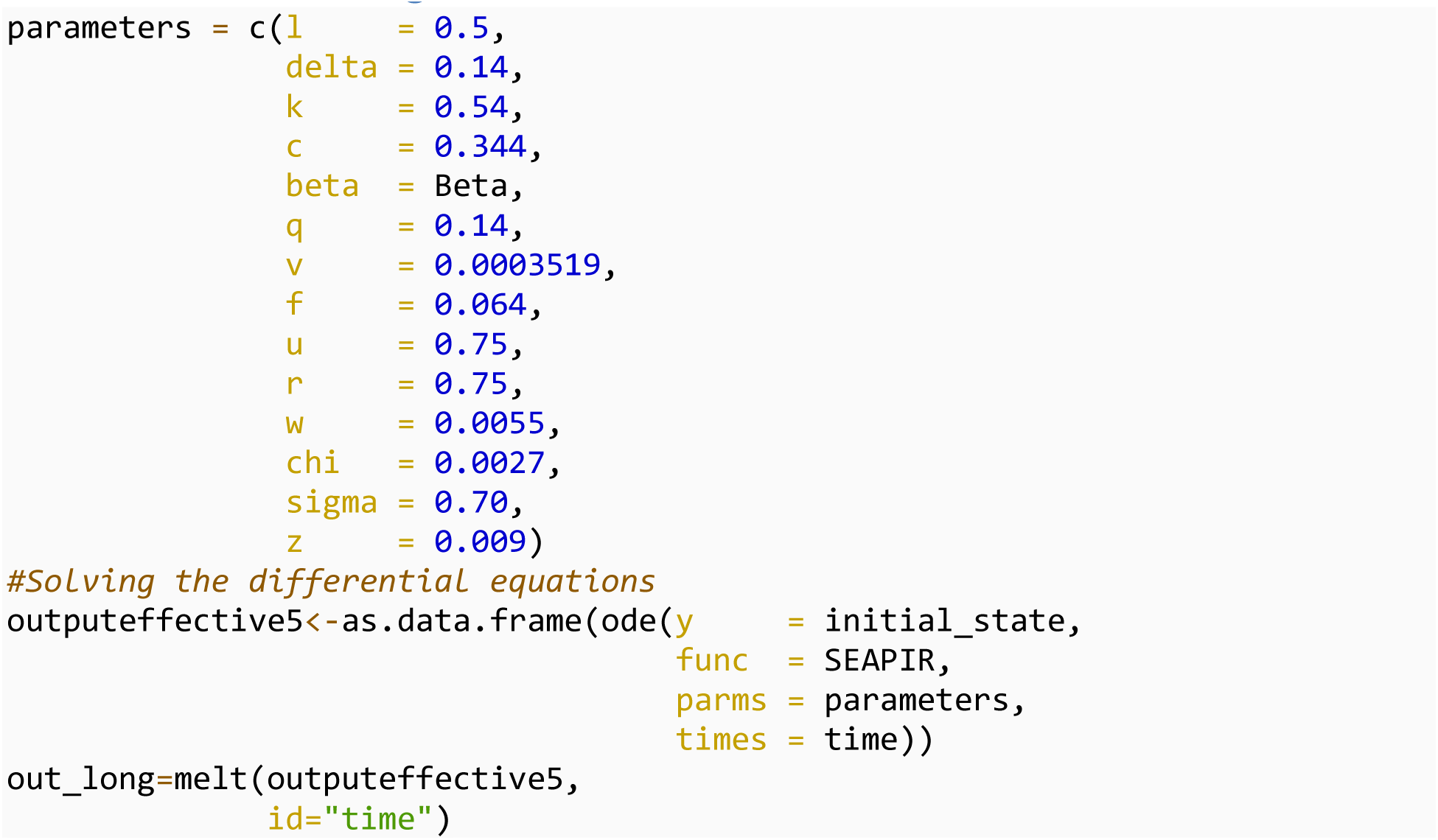

### Plot

Next we plot our results.

### Plot the total number deaths for each beta value

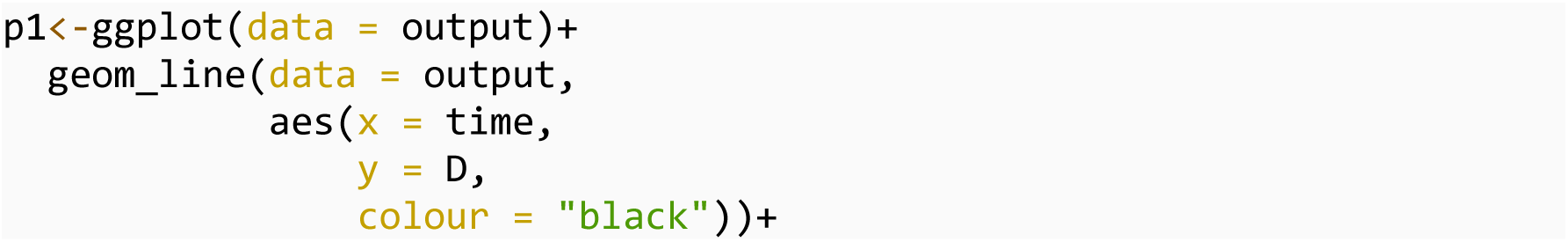
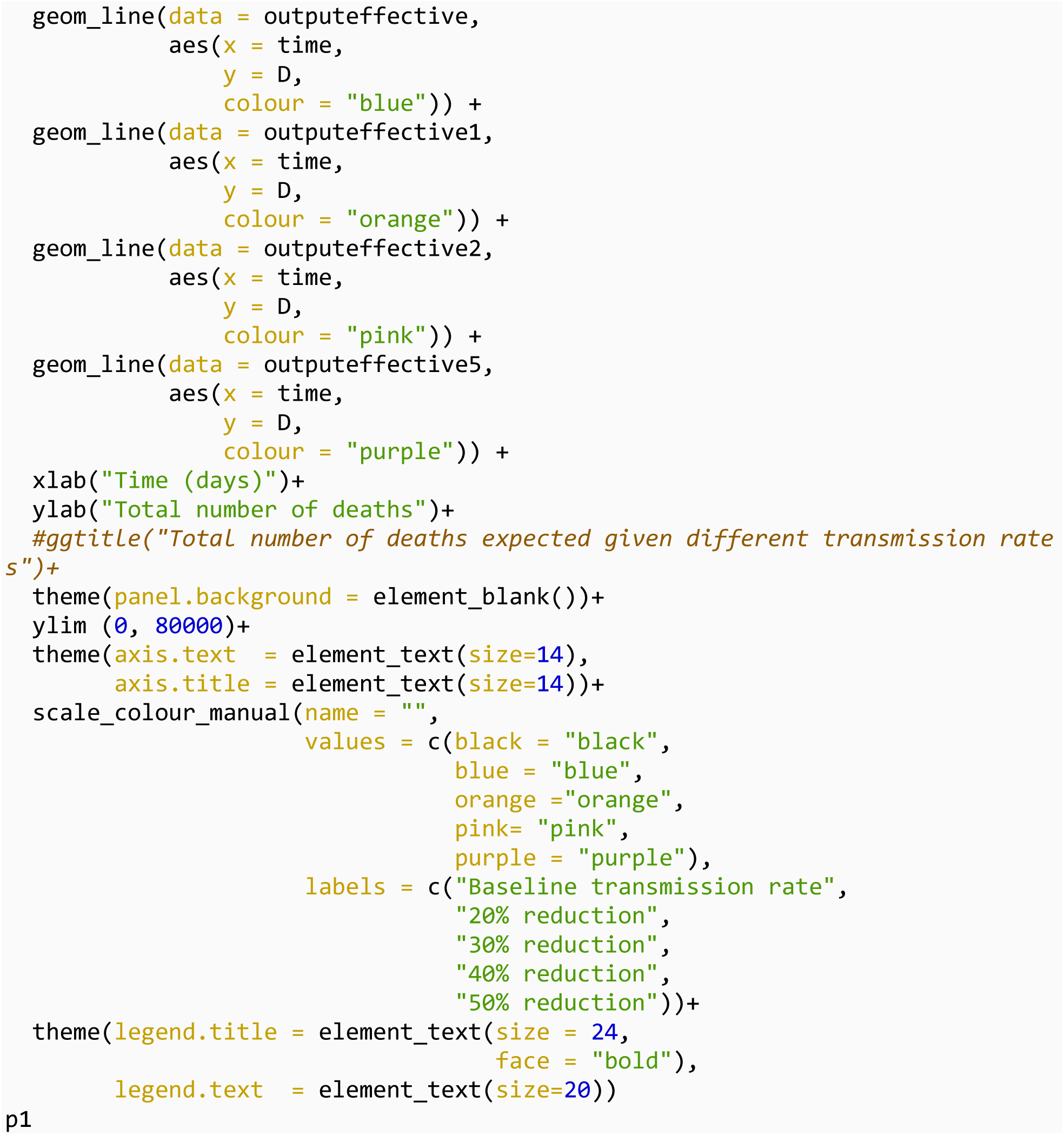

### Plot the number of symptomatic infected individuals at the peak

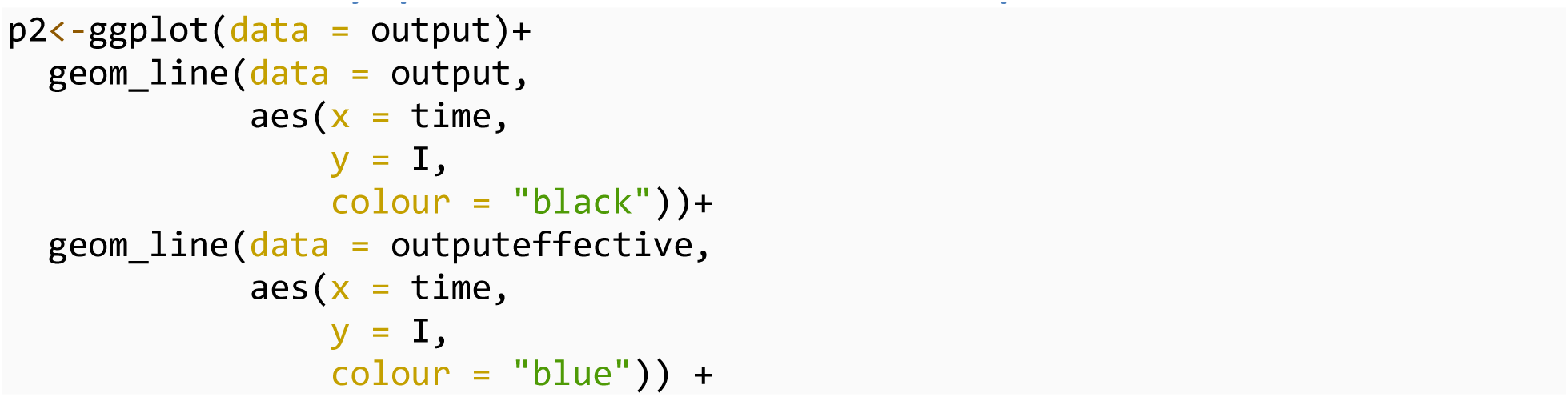
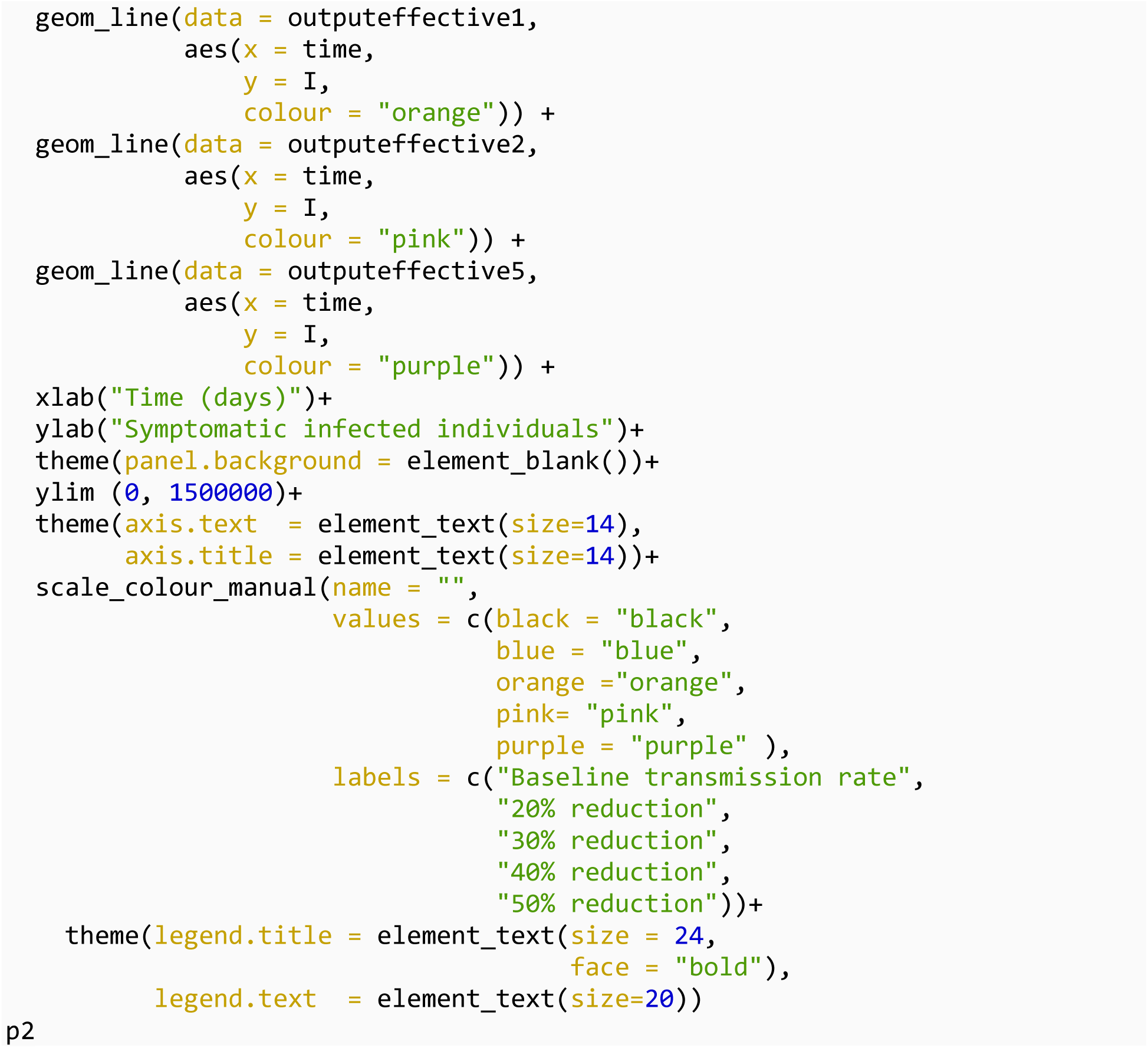

### Plot for cumulative infections

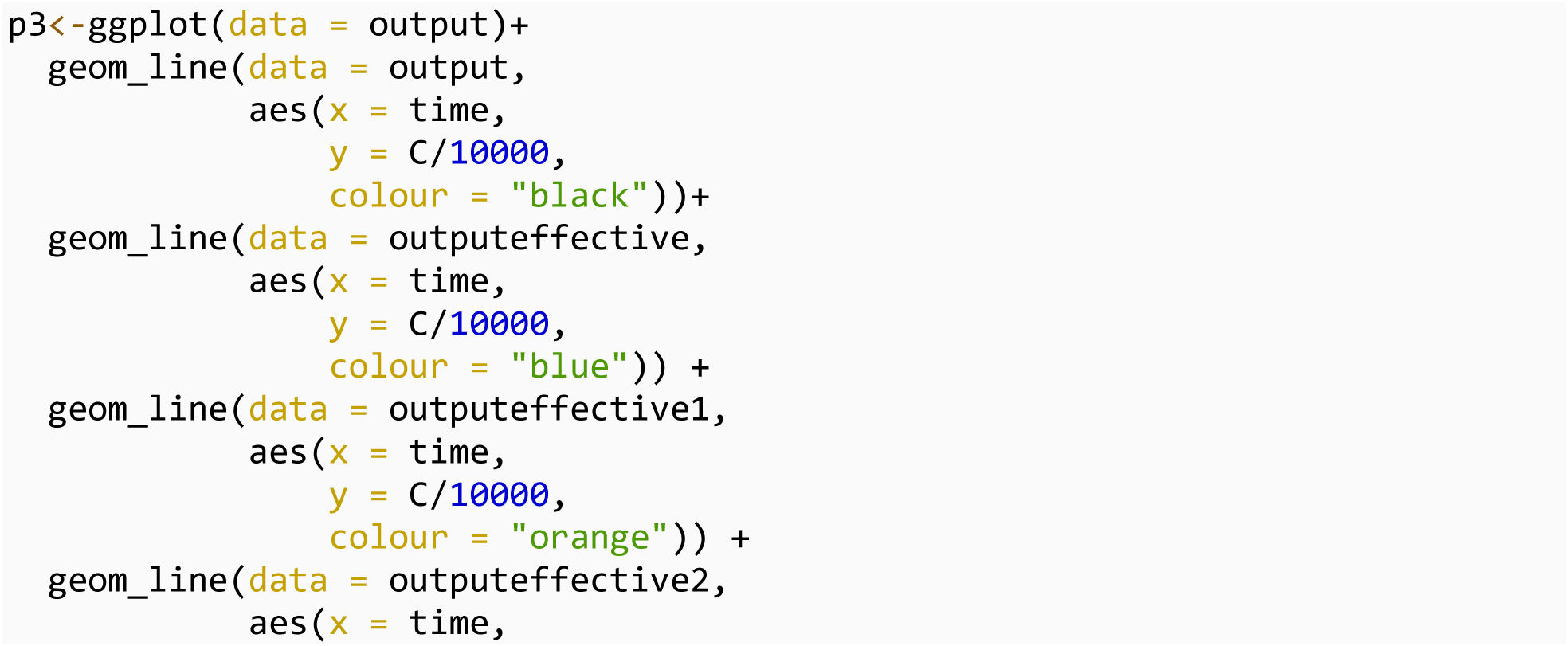
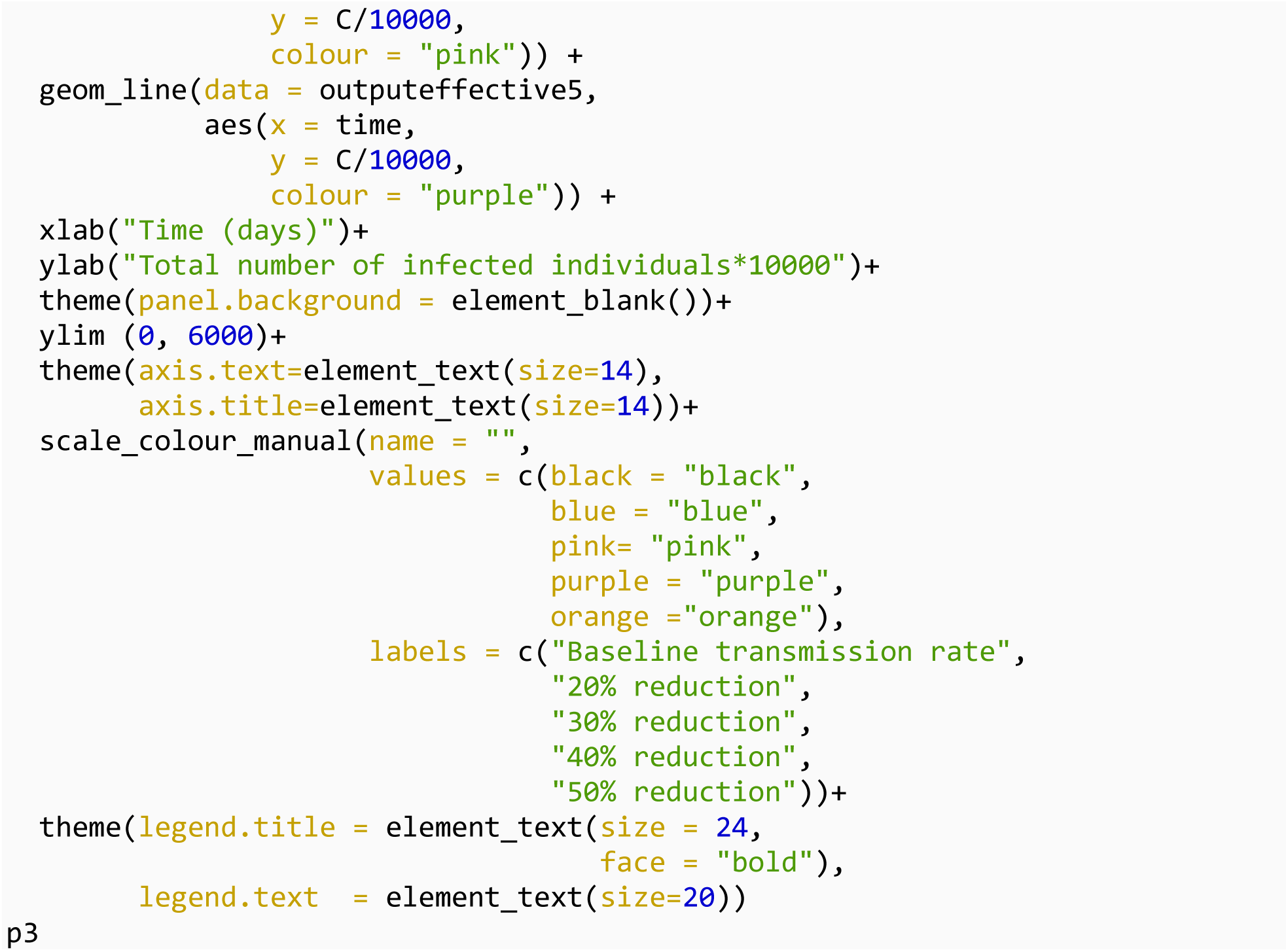

### Combination of multiple plots into one figure

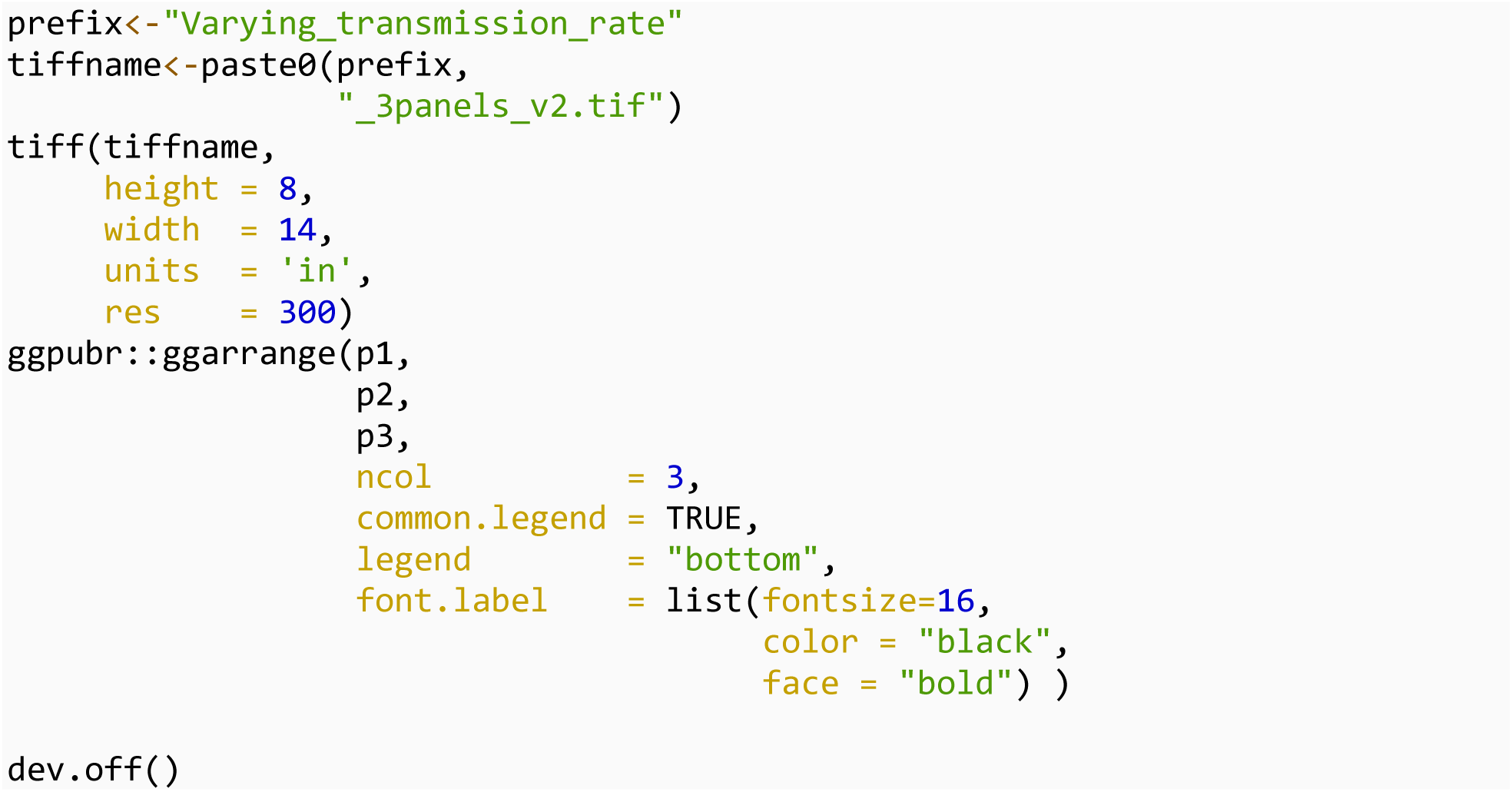

## The impact of vaccination coverage on outcomes

Next we explore the impact of vaccination coverage on outcomes. The daily vaccination rate was calculated by assuming various population coverage can be achieved in four months.

### First scenario

We used the coverage as of October 2021 (2.8% of population)= “current vaccination rate”

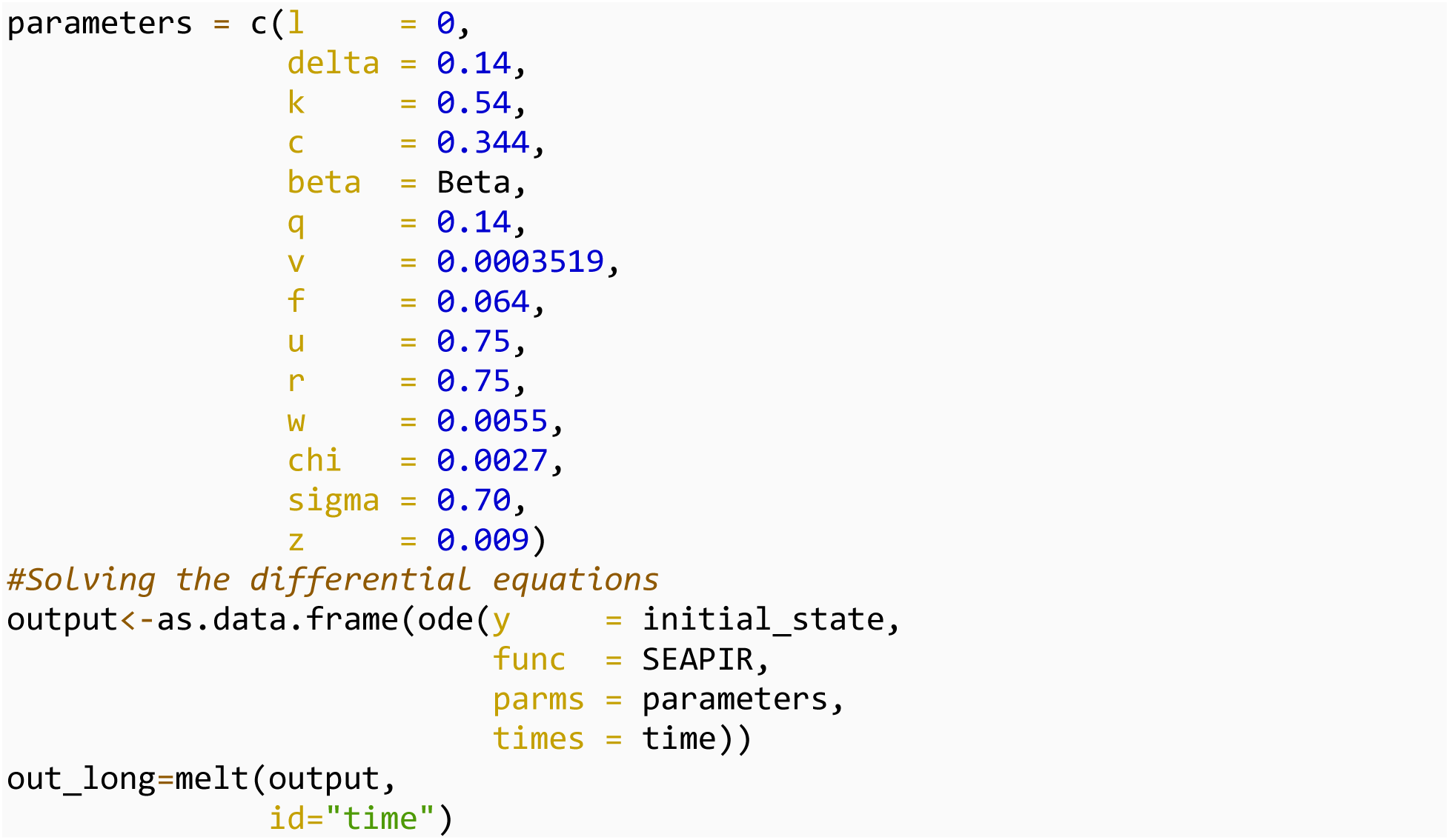

### Second scenario

*v* = 0.0007 doubling current vaccination rate

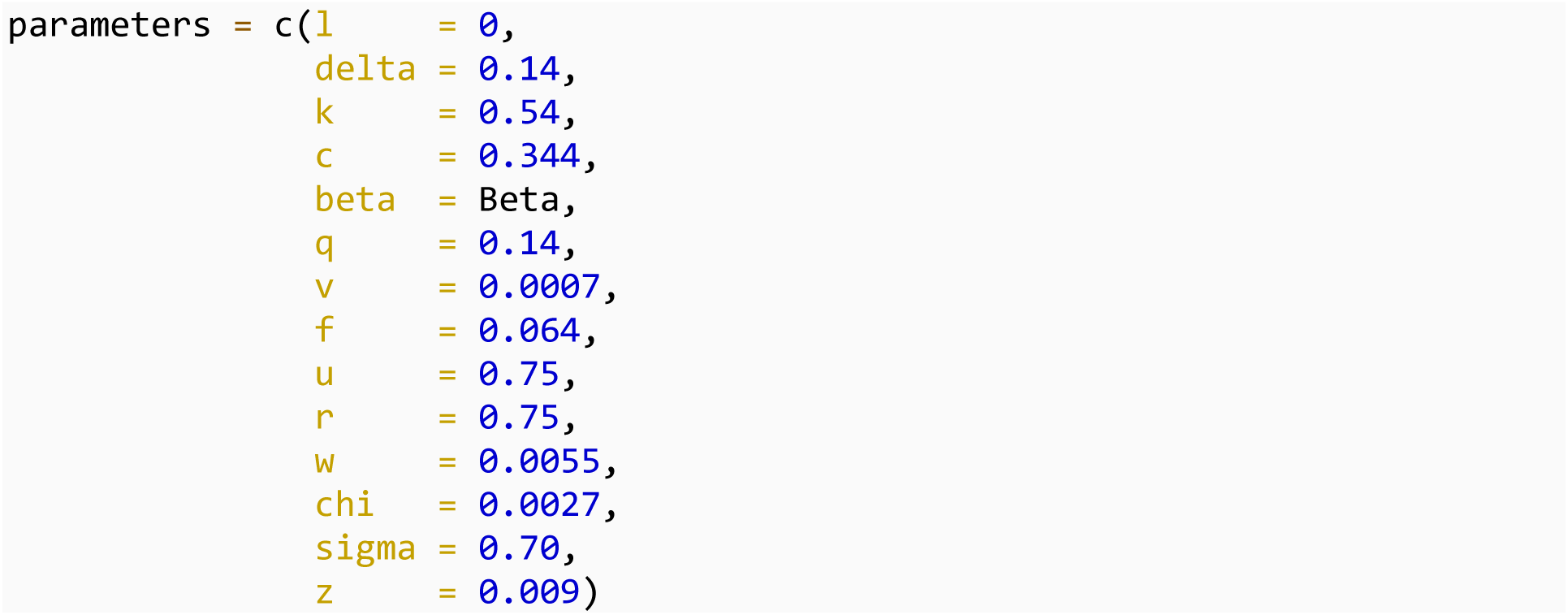
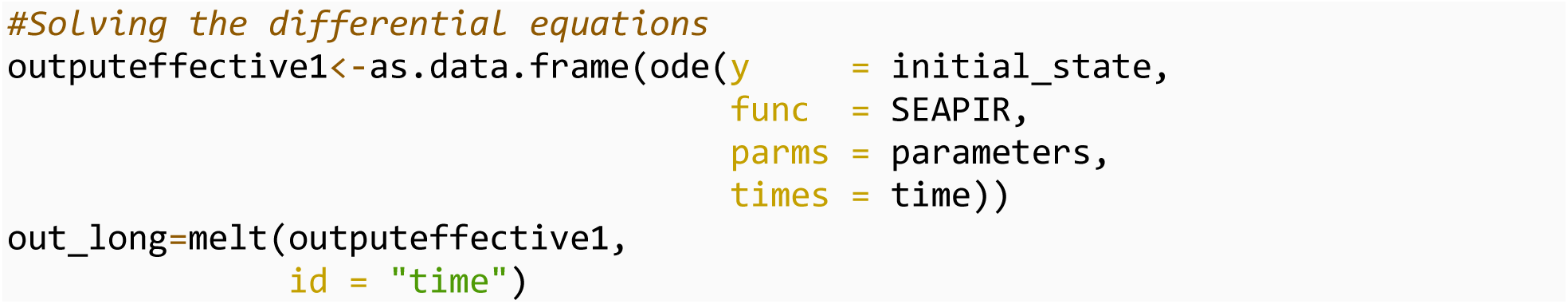

### Third scenario

*v*=0.0017, 20% of population vaccinated

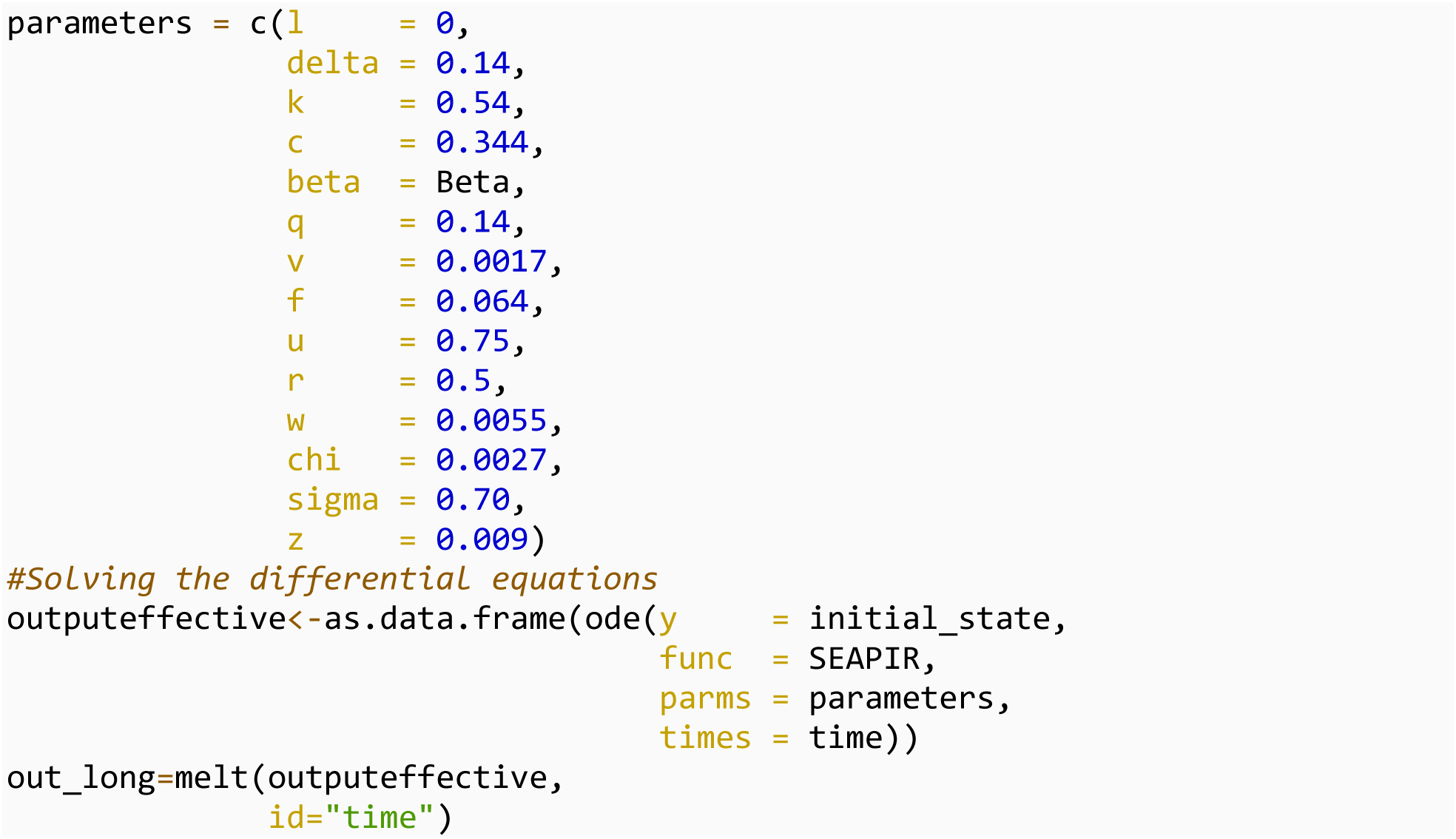

### Fourth scenario

*v*= 0.0042 50% vaccine coverage

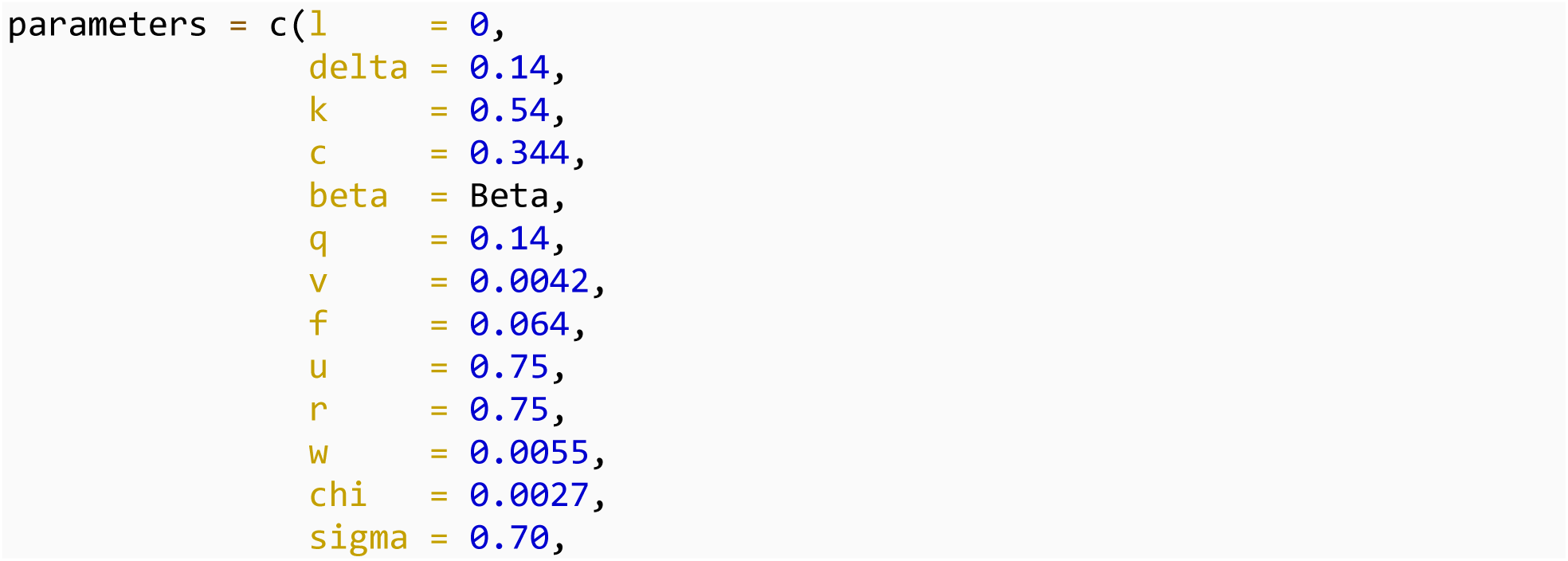
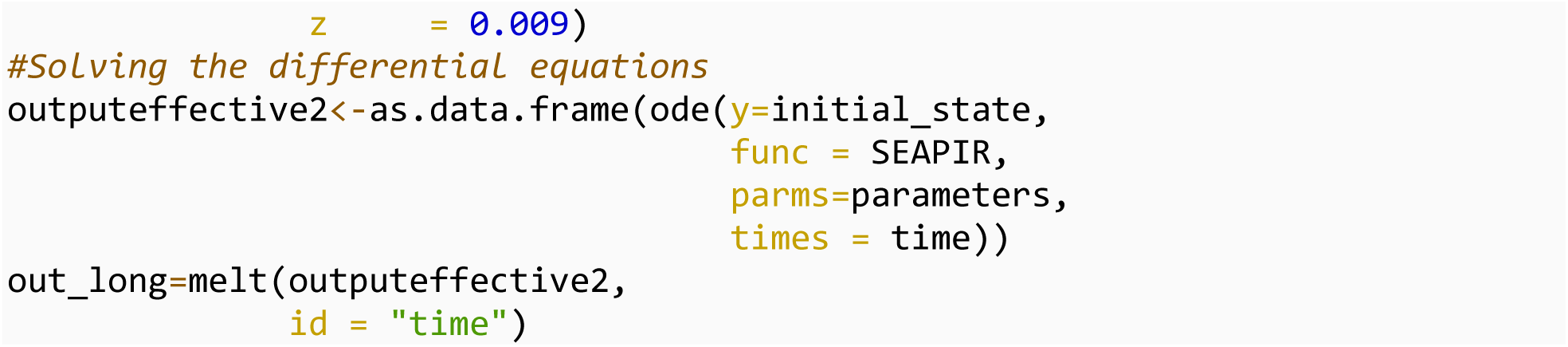

### Fifth scenario

*v*= 0 no vaccination

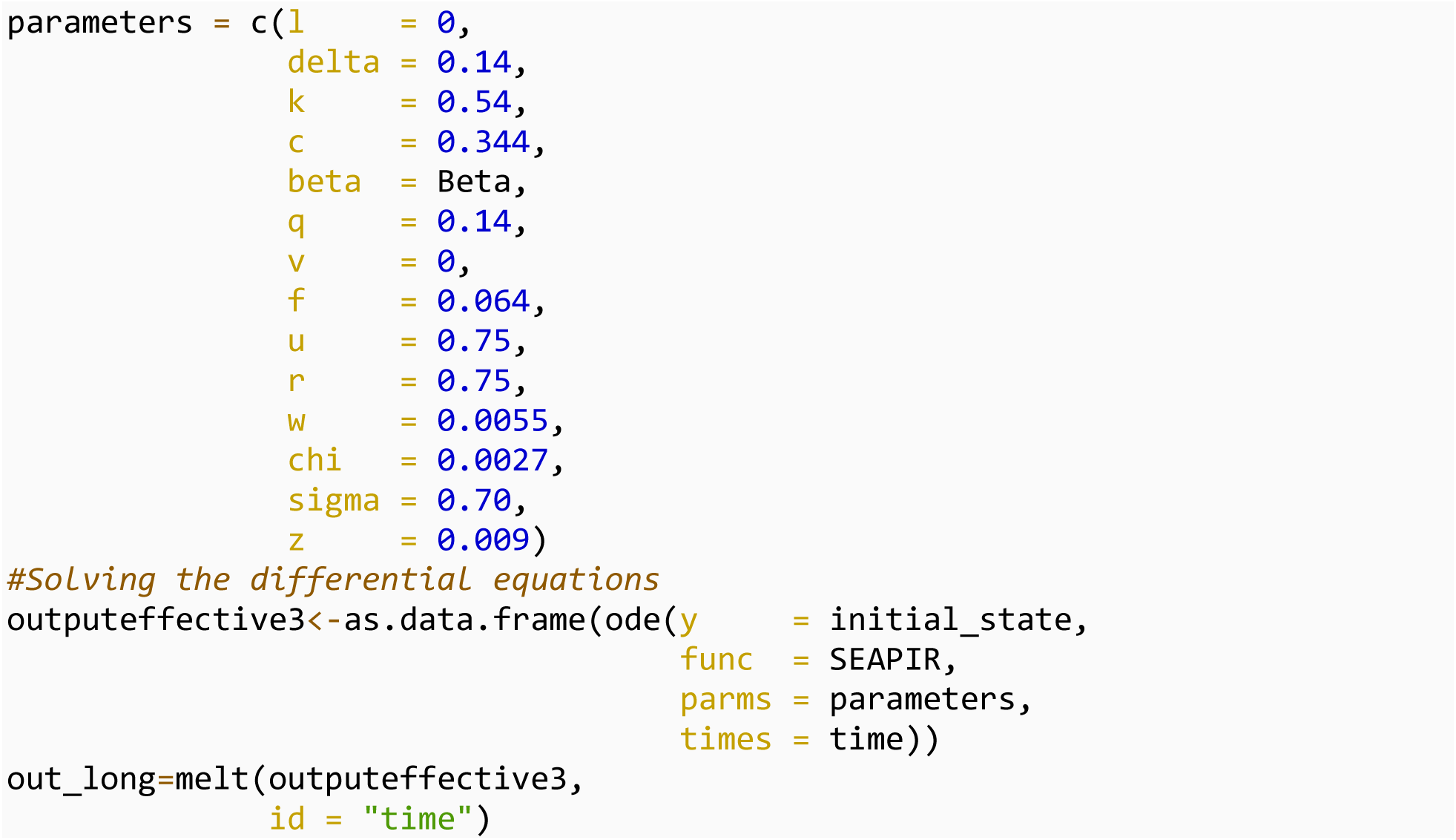

### Plot

### Number of deaths

We plotted number of deaths for varying vaccination rate.

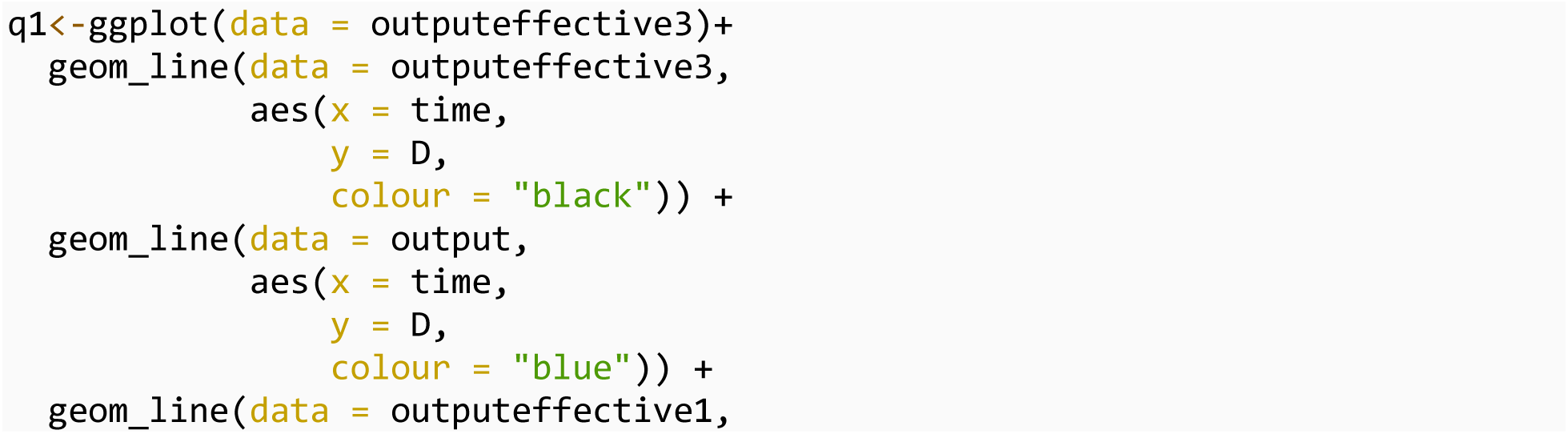
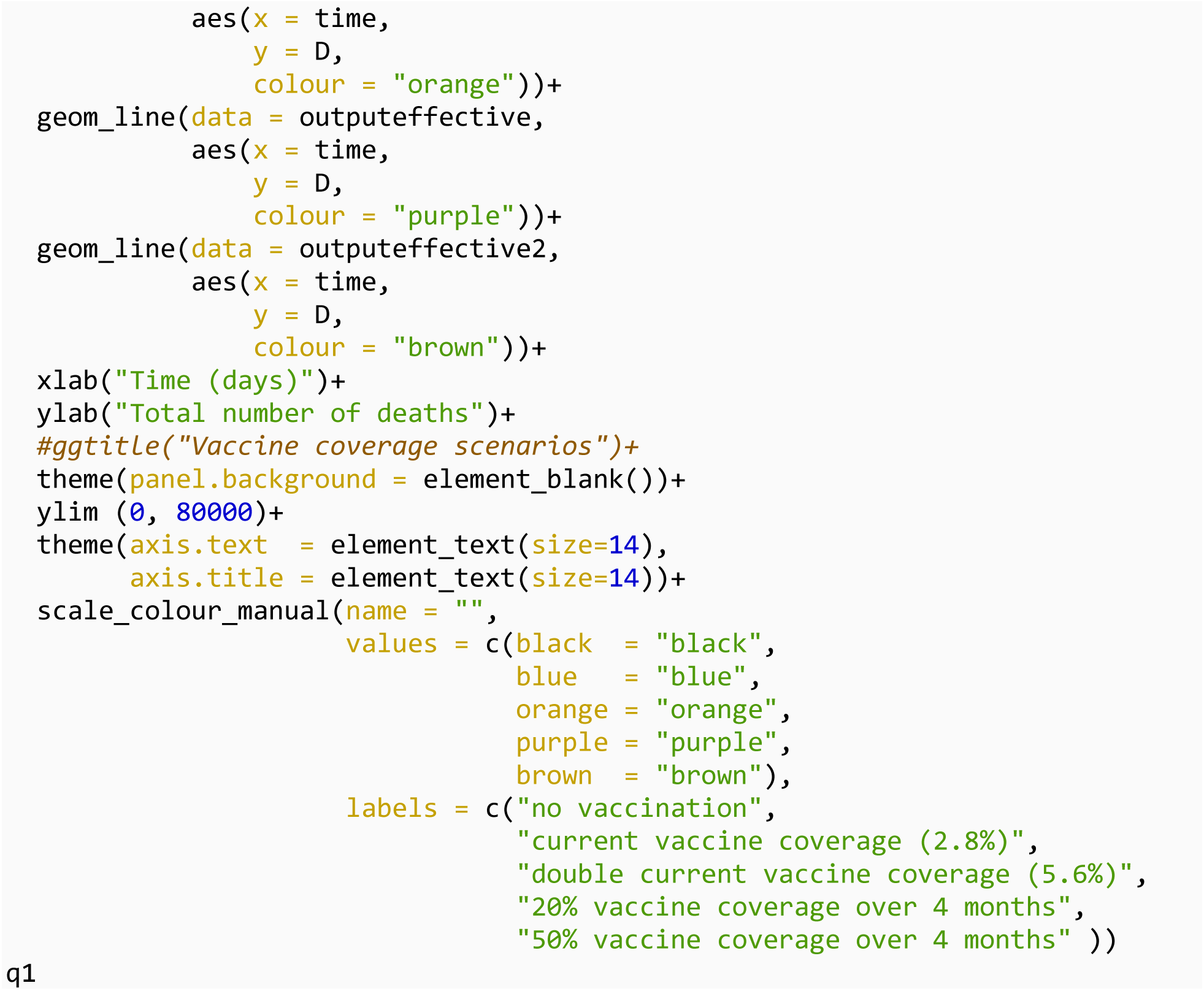

### Size of the peak

We plotted the number of symptomatic infected individuals at the peak for varying vaccination rate

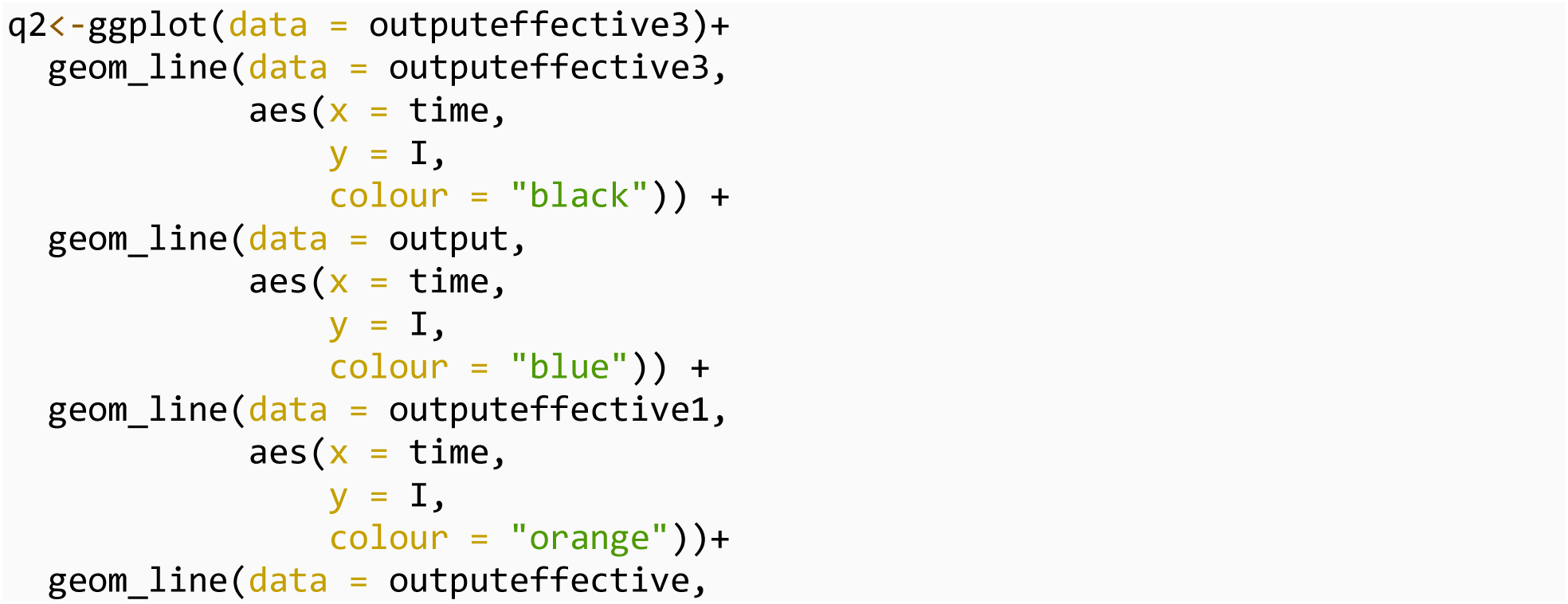
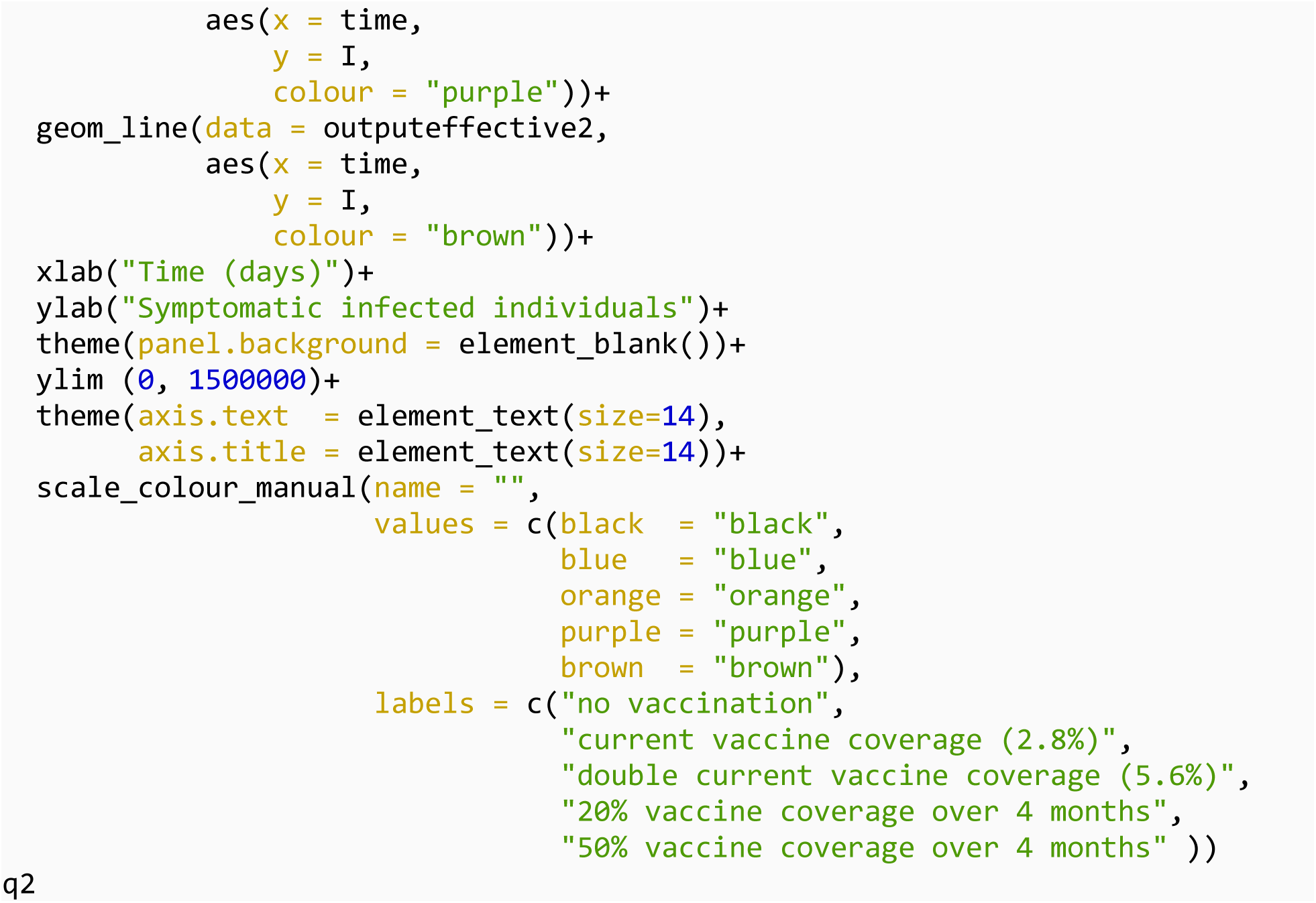

### Final outbreak size

We plotted total number of infected individuals for varying vaccination rate

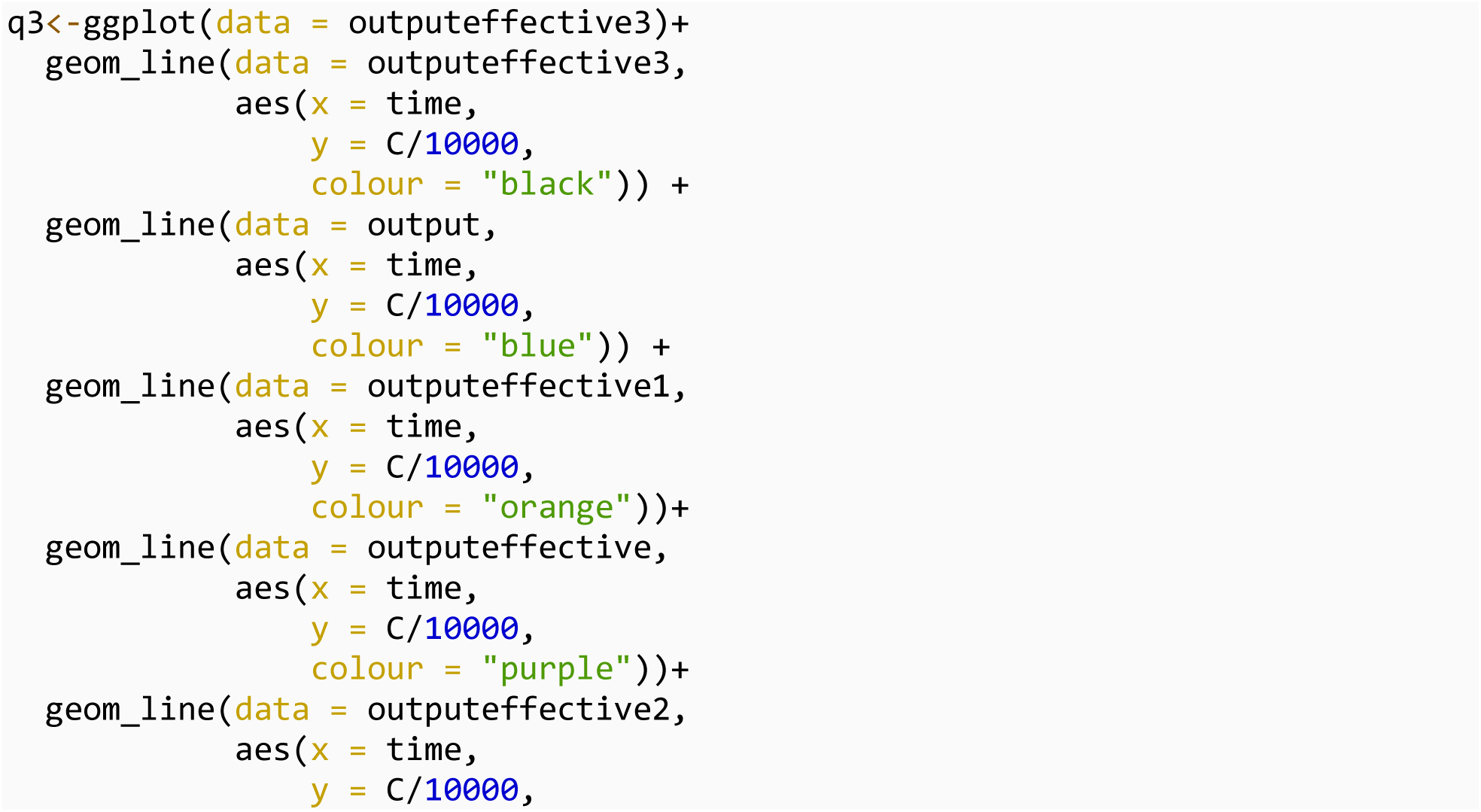
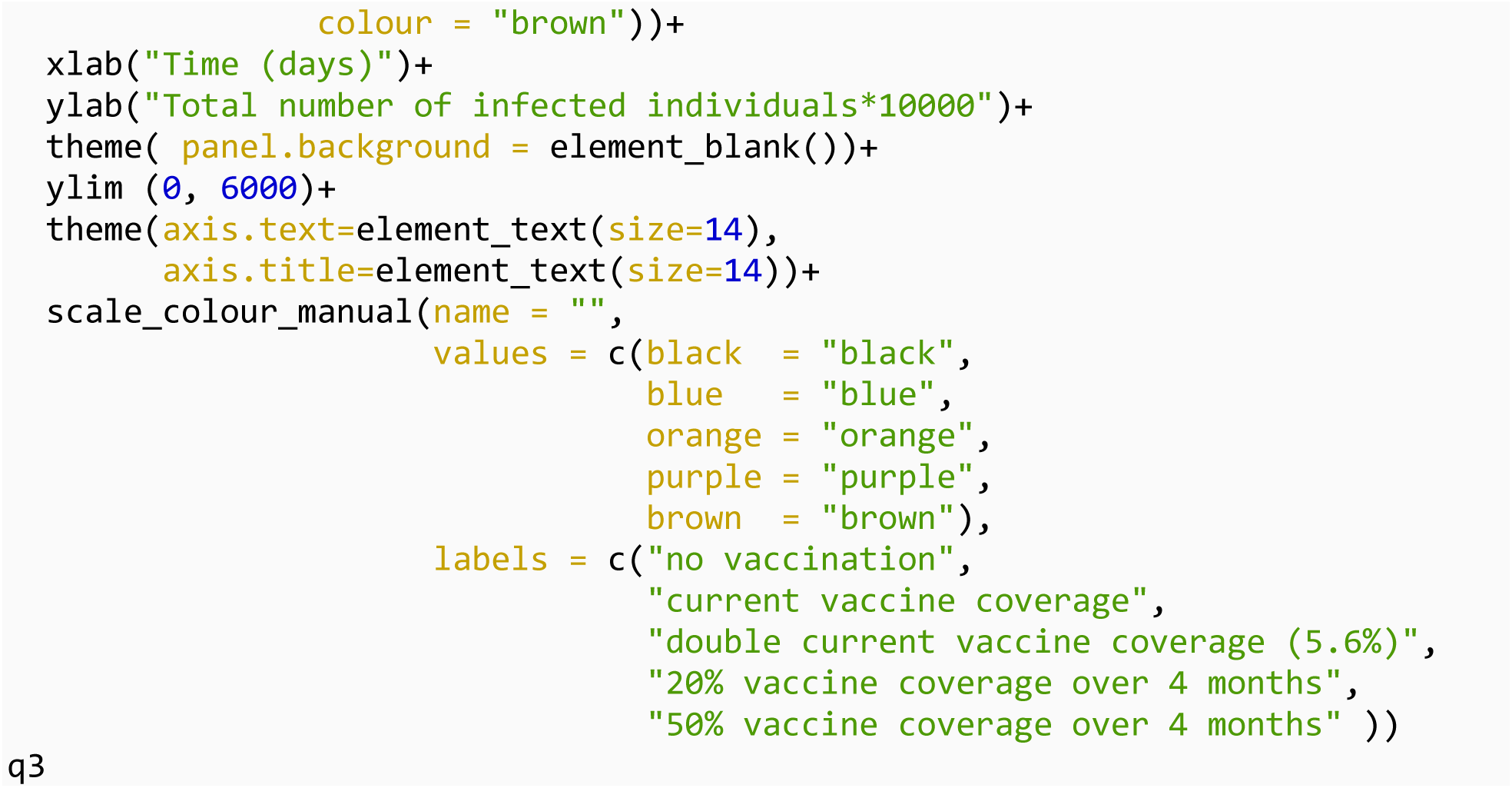

### Combination of multiple plots into one figure

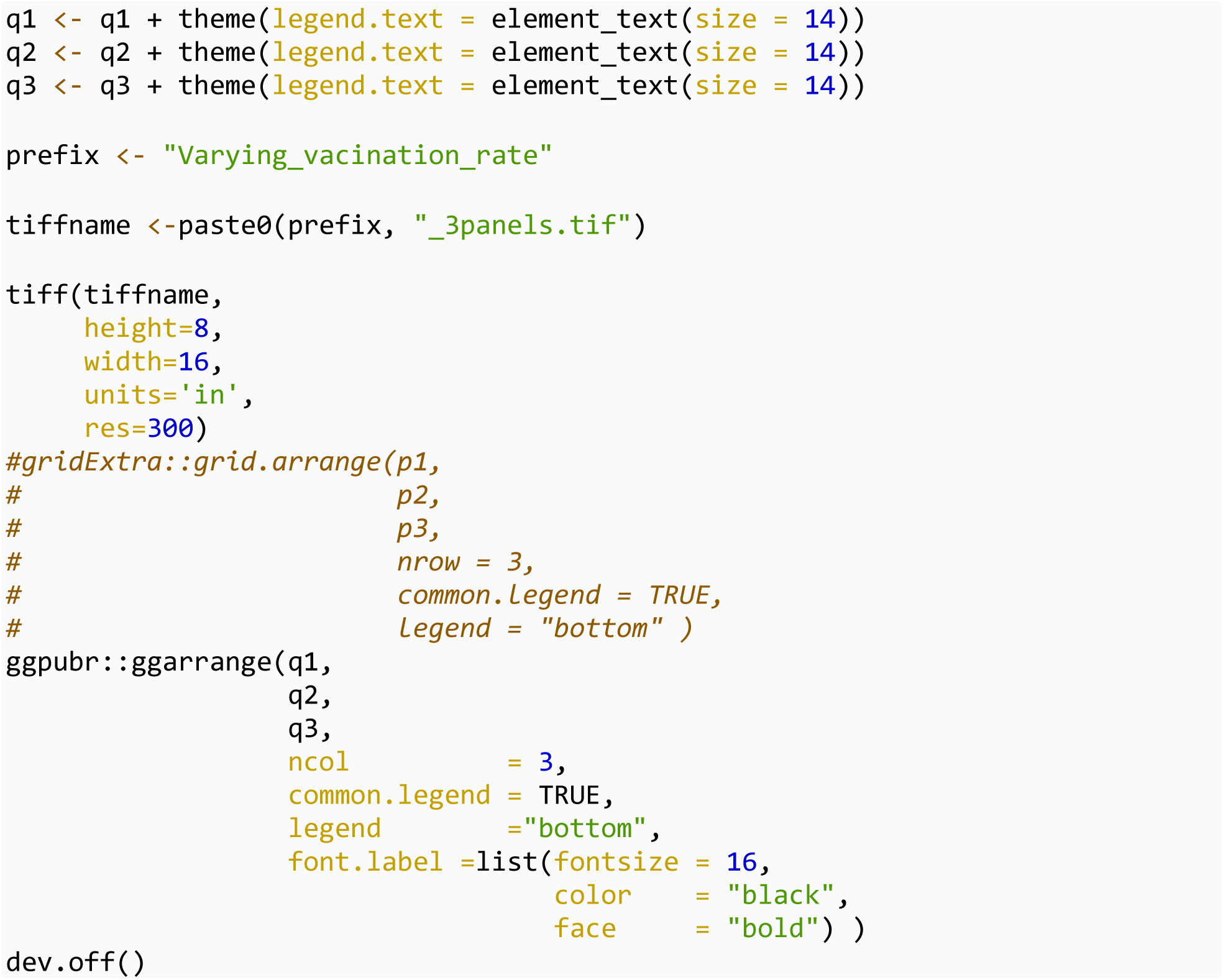

## Scenario analyses presented in heat maps

We estimated the outcomes by varying both the decline in transmission rate and increase in vaccination rate.

To simulate values for the two parameters over the range to facilitate plotting

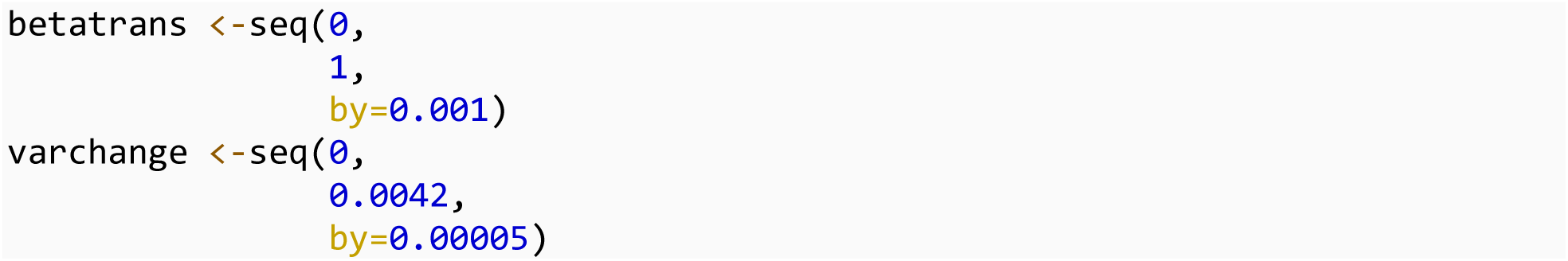

To create a matrix for the set of values and assign column and row names

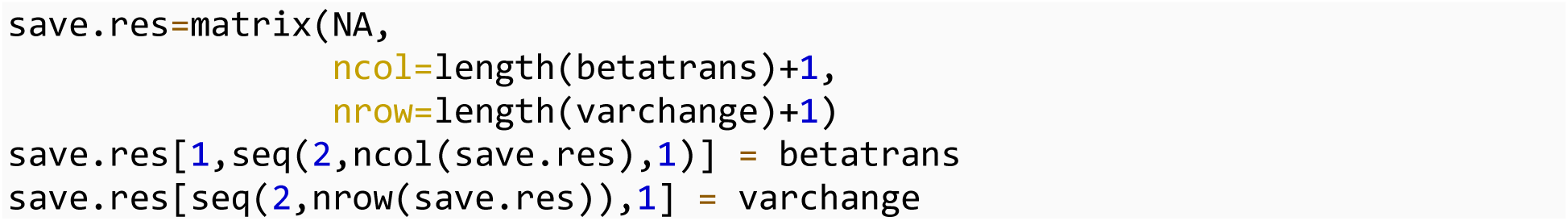

To create a nested loop for the given values of *l* and *v*

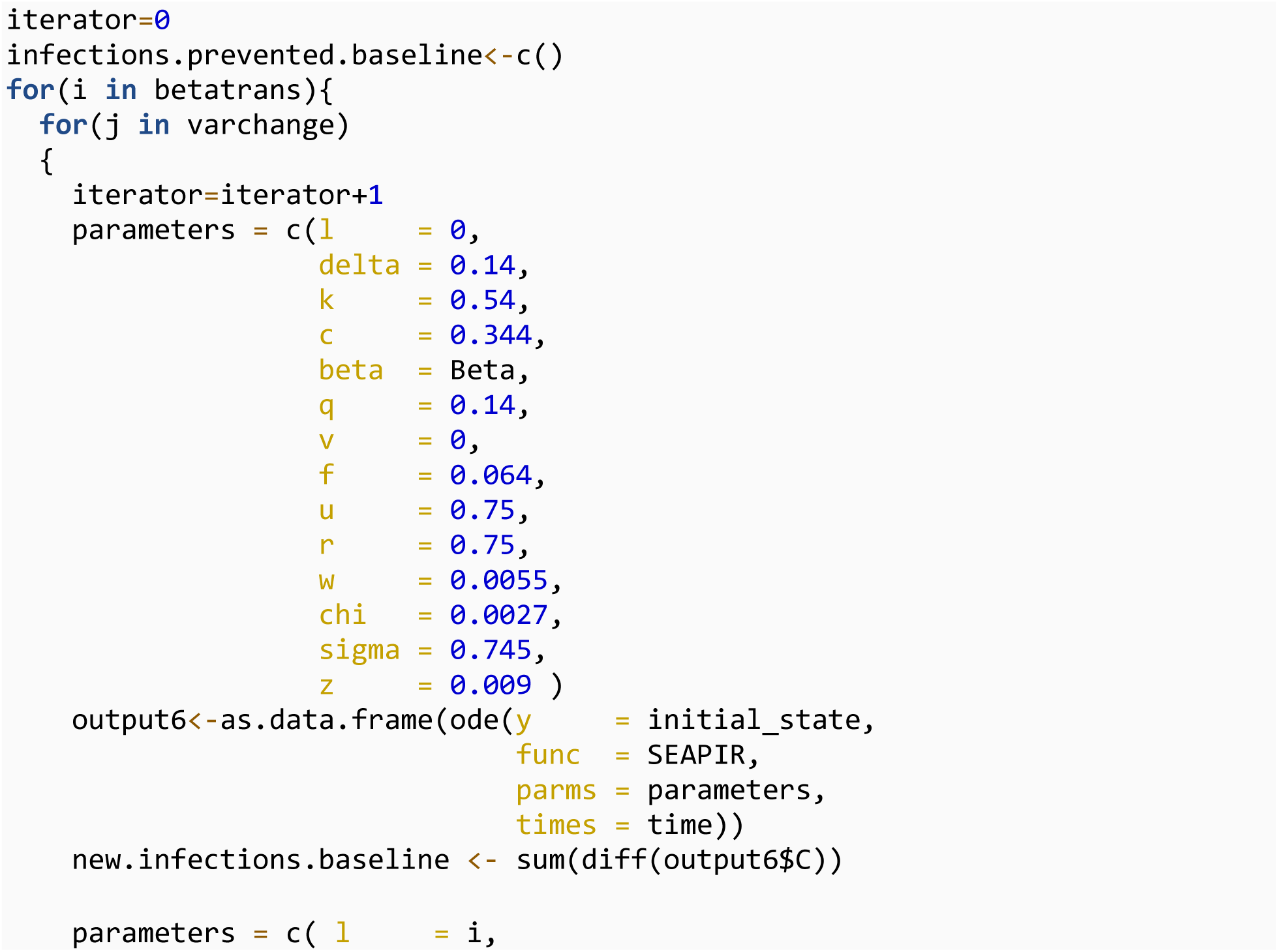
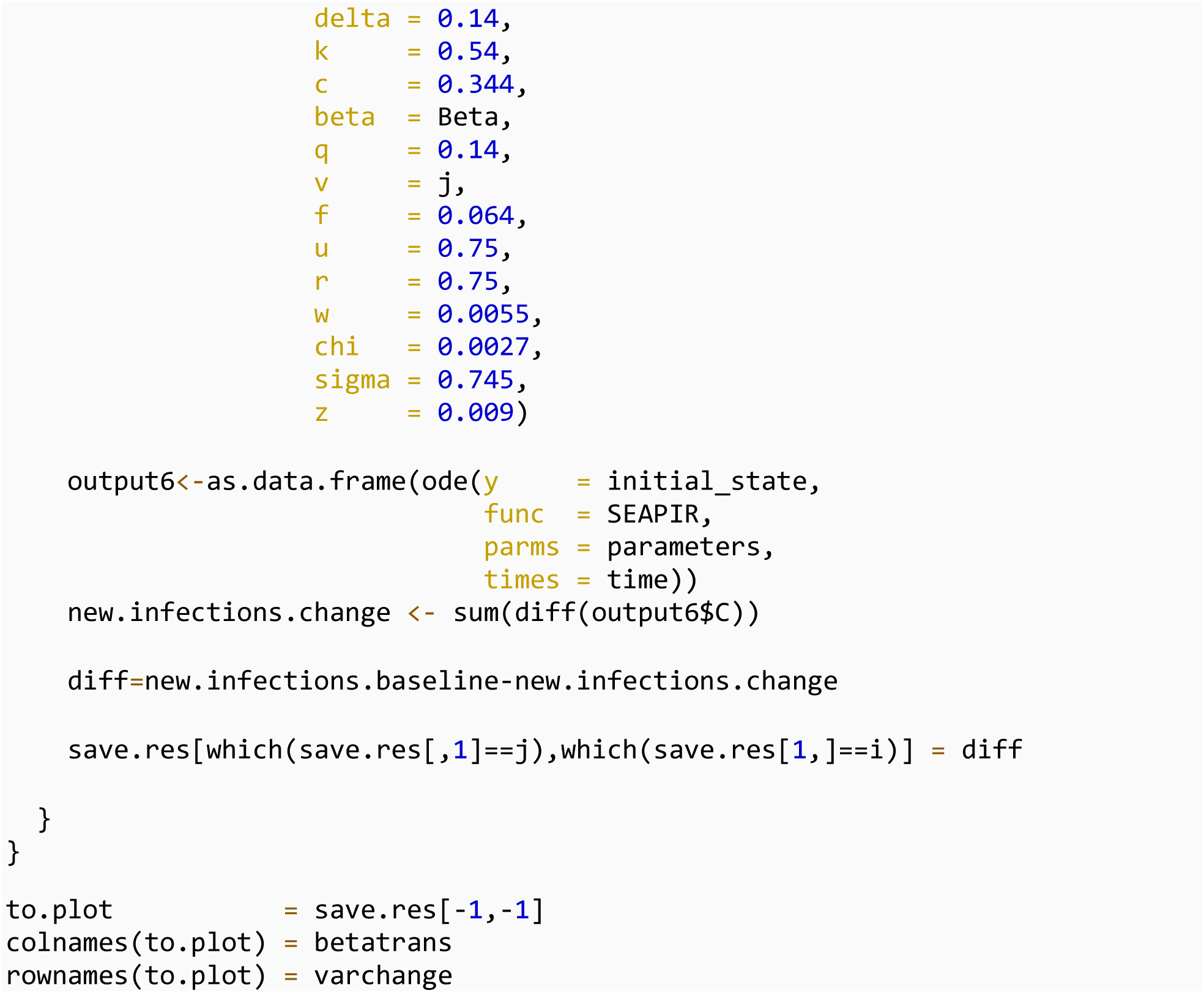

You may save the results as an rds file as needed:

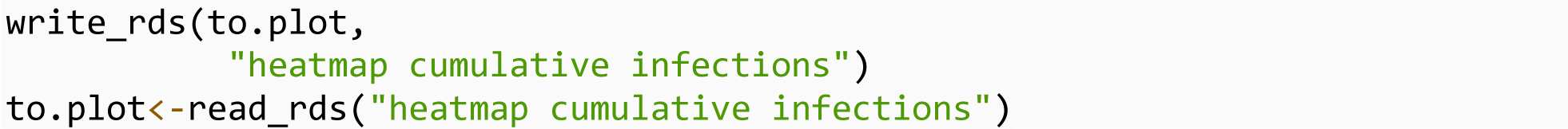

Save the heatmap as a tiff file:

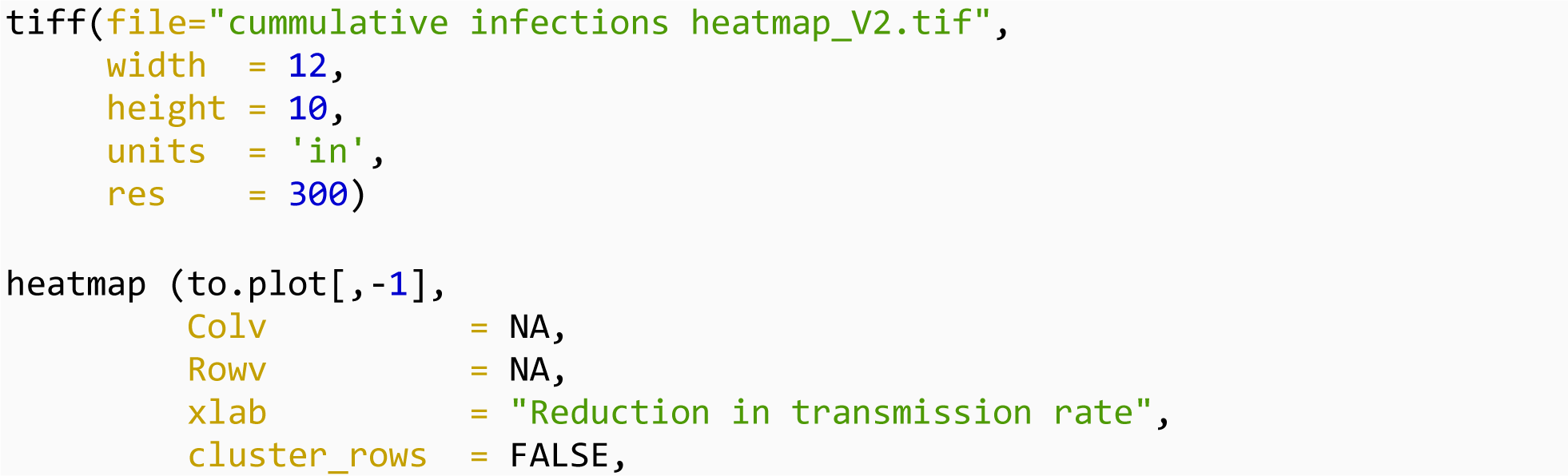
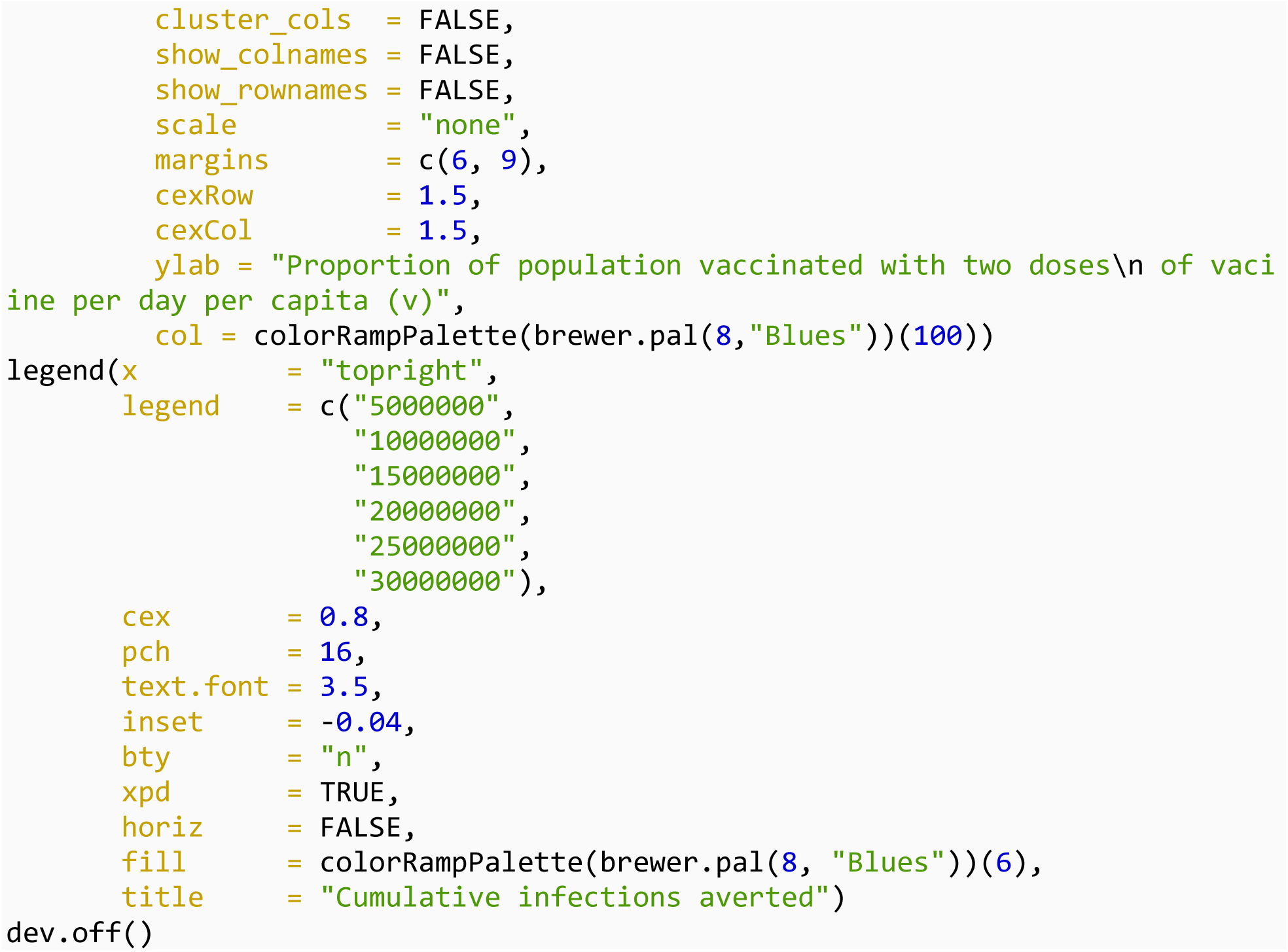

### Cumulative deaths averted

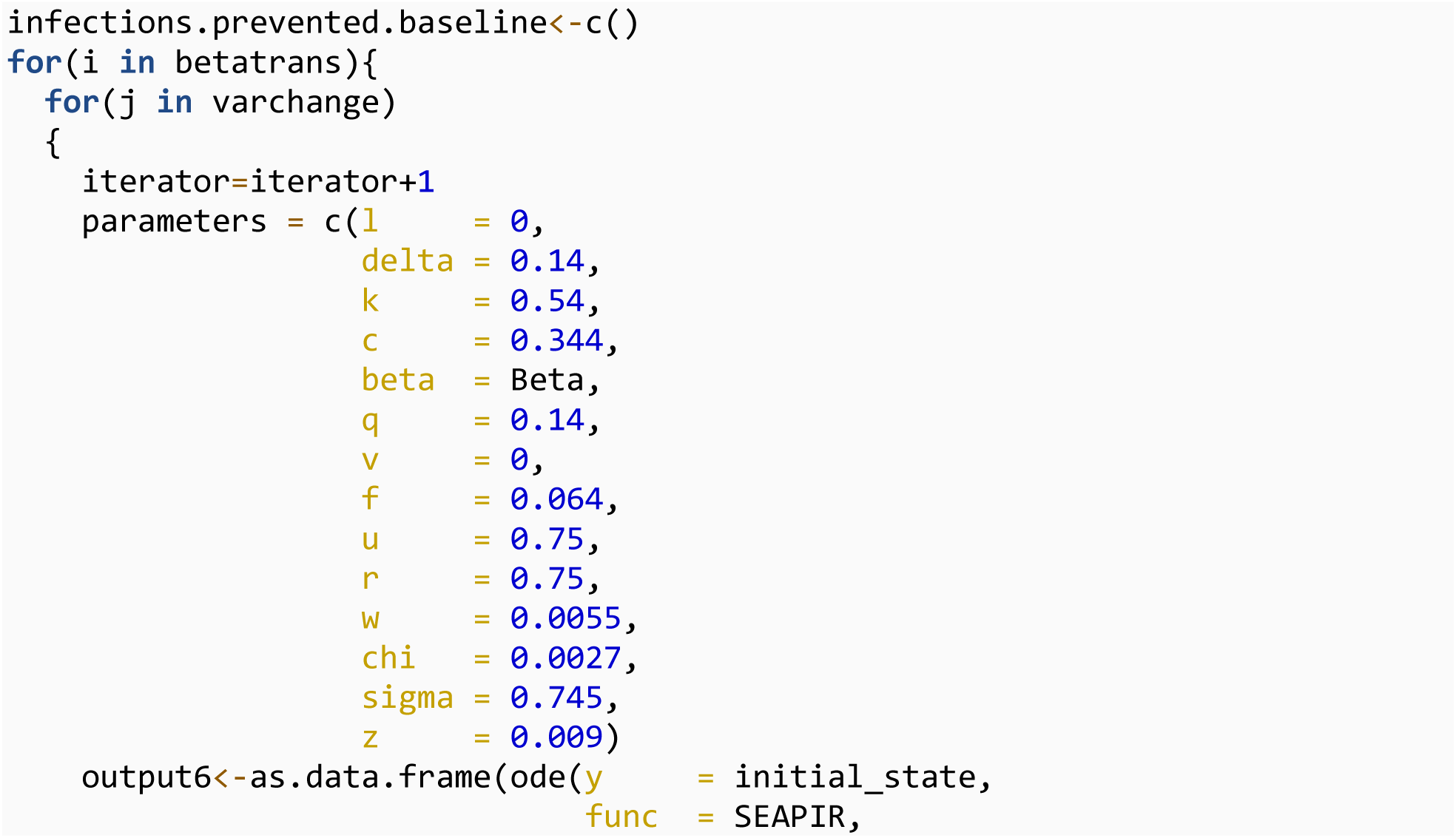
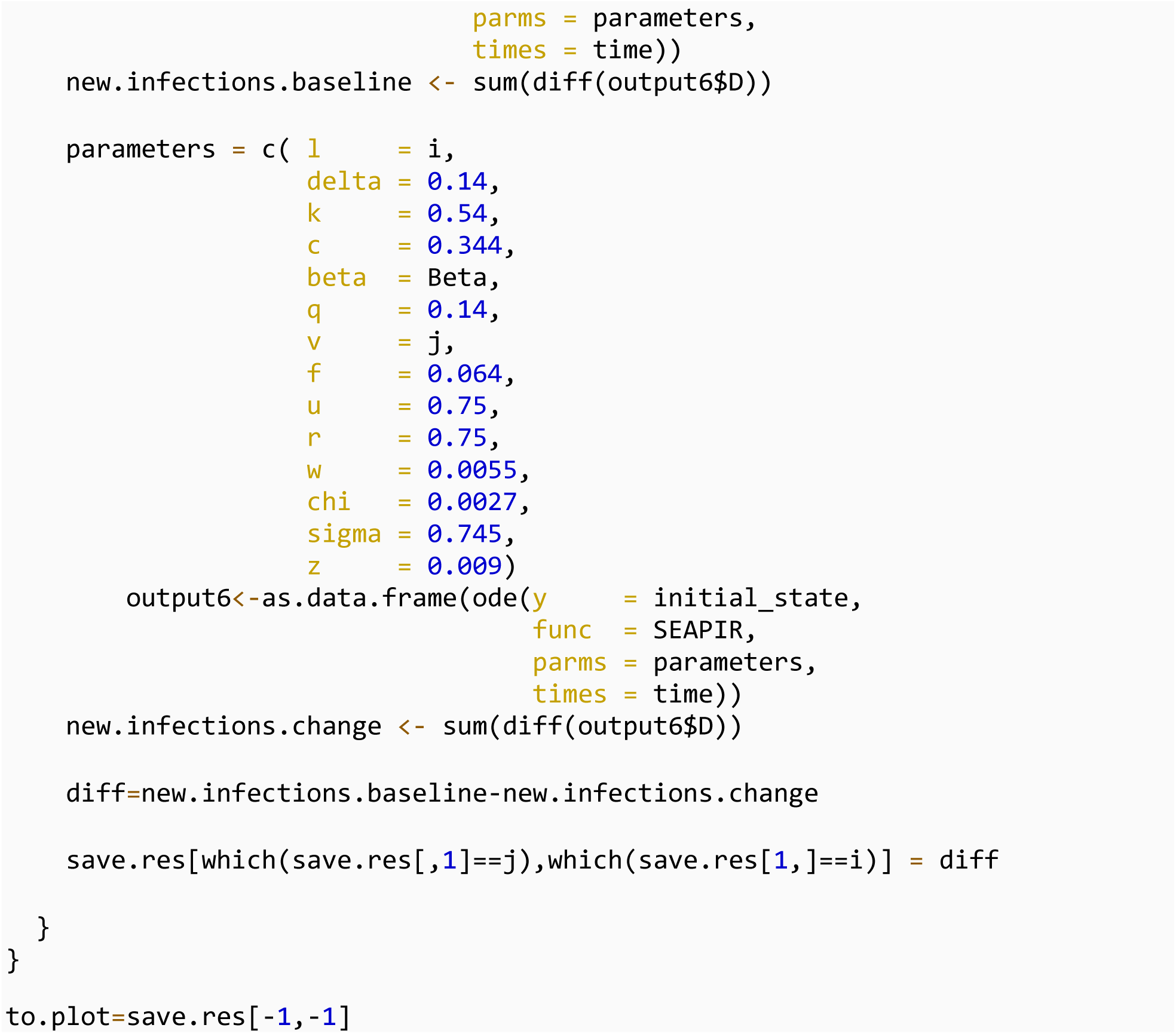

You may save the results as an rds file as needed:

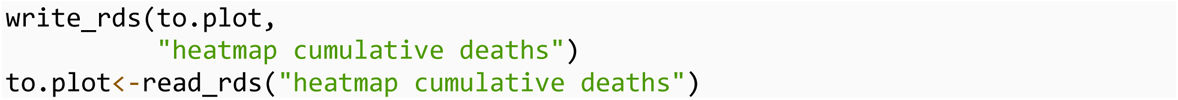

Save the heatmap as a tiff file

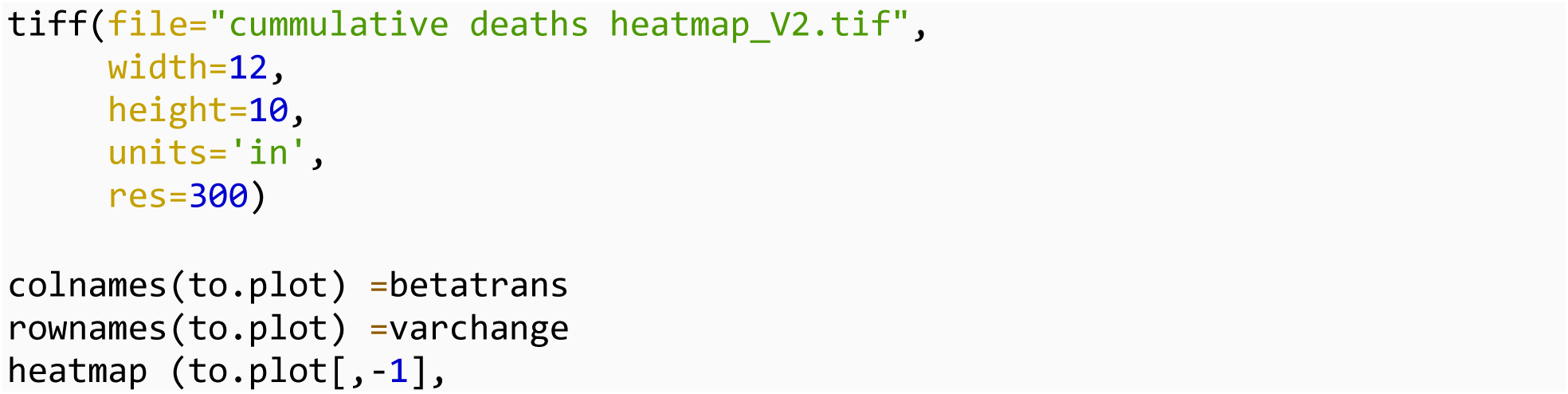
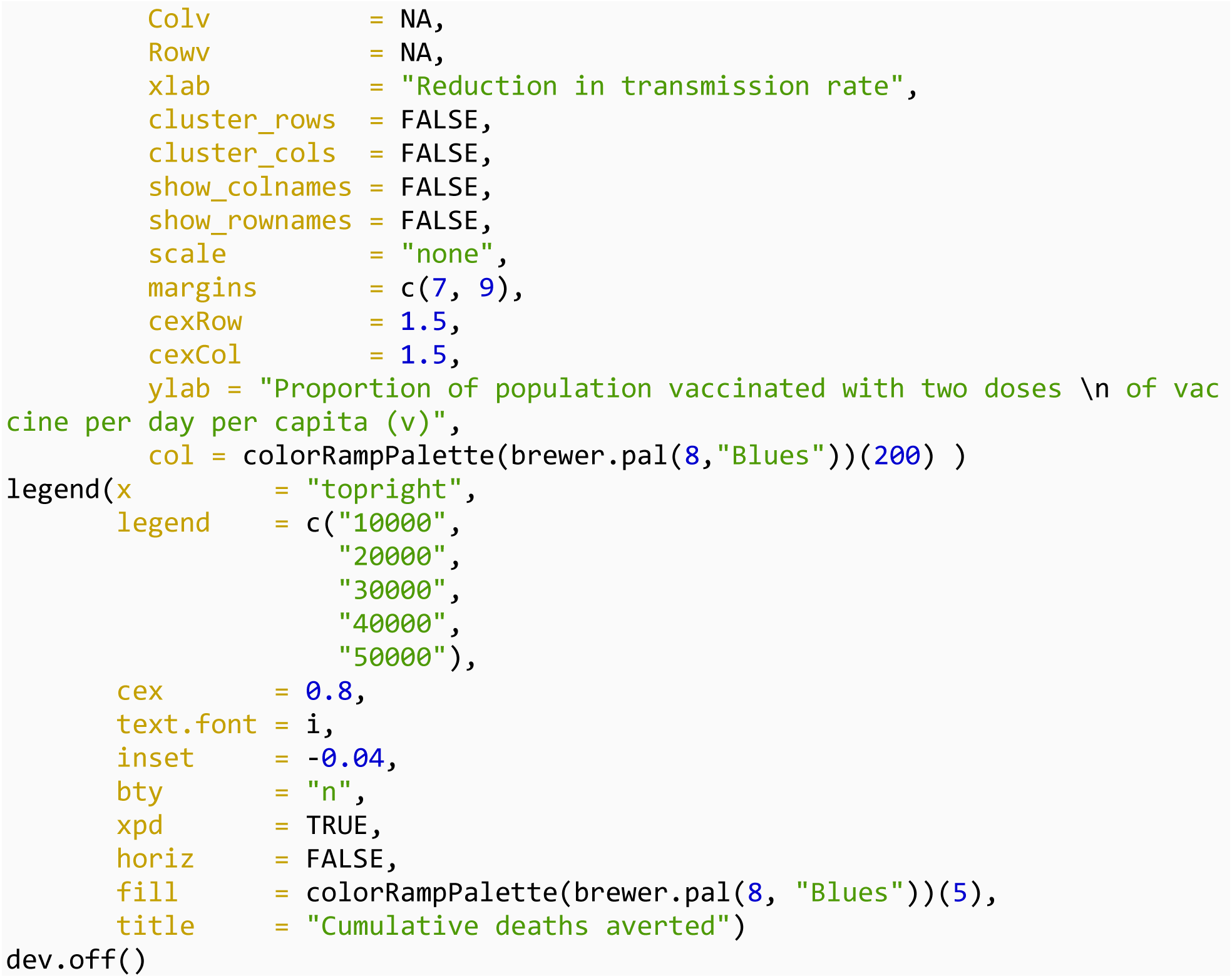

## Modeling deaths and cumulative infections averted with decline in transmission and increase vaccination respectively

We first start with cumulative infections averted. To model deaths averted,

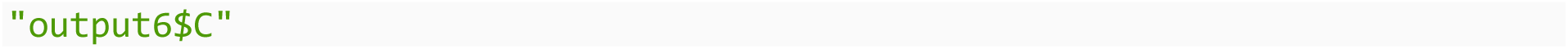

is replaced with

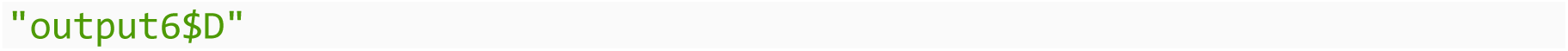

in the following codes

### The cumulative infections averted by varying the decline in transmission rate from 0 to 1

We created a loop to estimate the infections averted as: “cumulative infections when there was no change in transmission” minus “cumulative infections at any given rate that transmission was reduced”

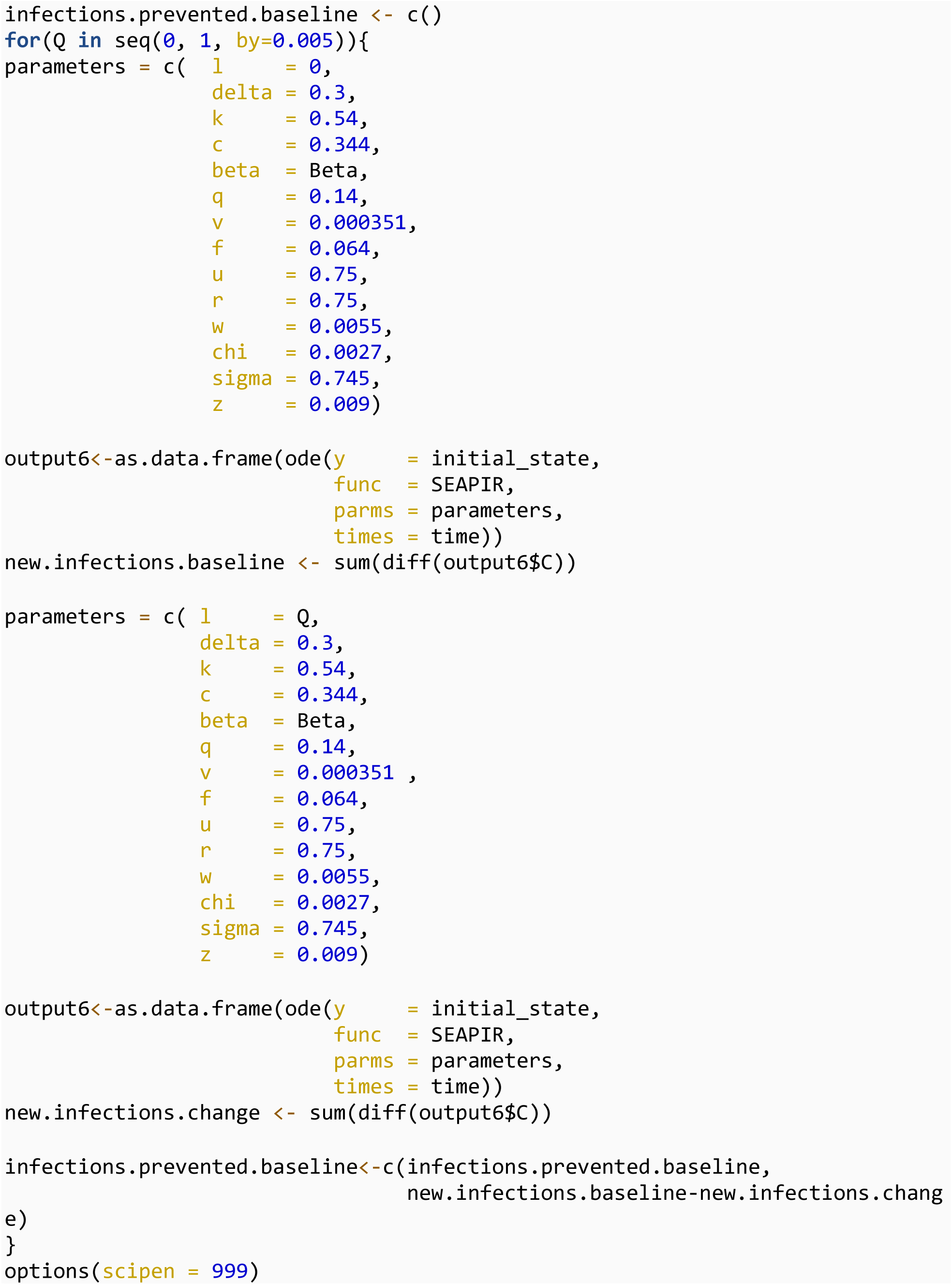

We can visualize the cumulative infections averted over time with a plot:

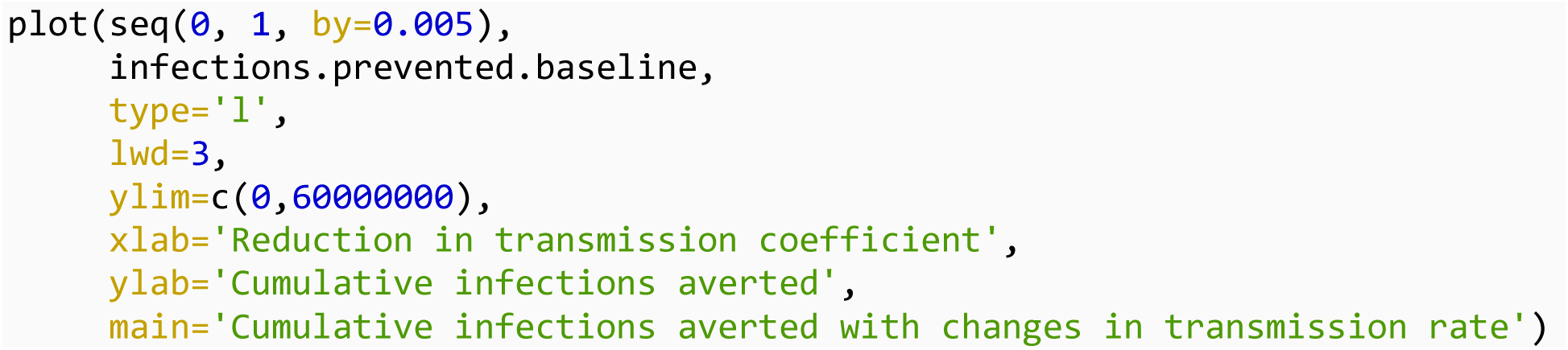

### Latin Hypercube Sampling

We assessed the uncertainty in the cumulative infections averted using Latin Hypercube sampling (LHS)

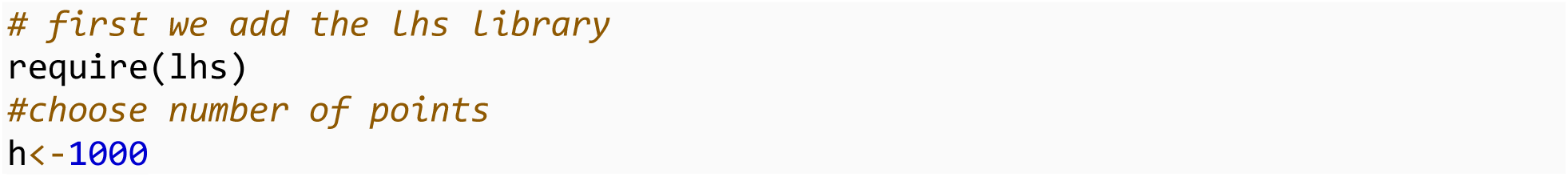

A Latin Hypercube Sample design was created for 13 uncertain parameters

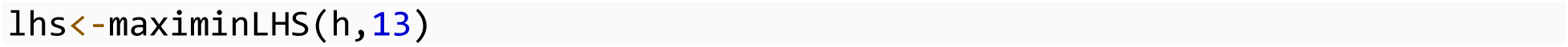

Values were assigned to the parameters for their uncertainties

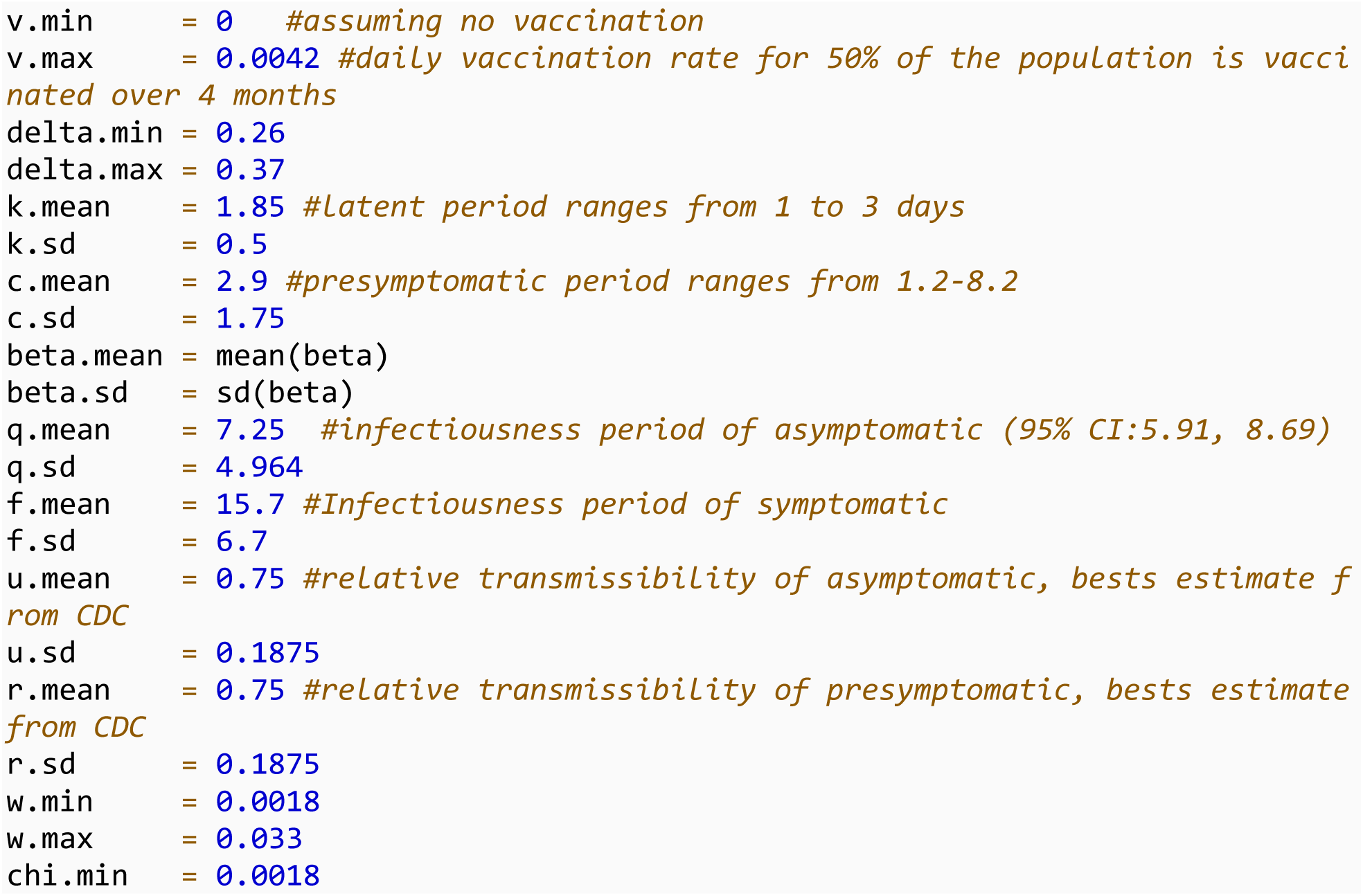
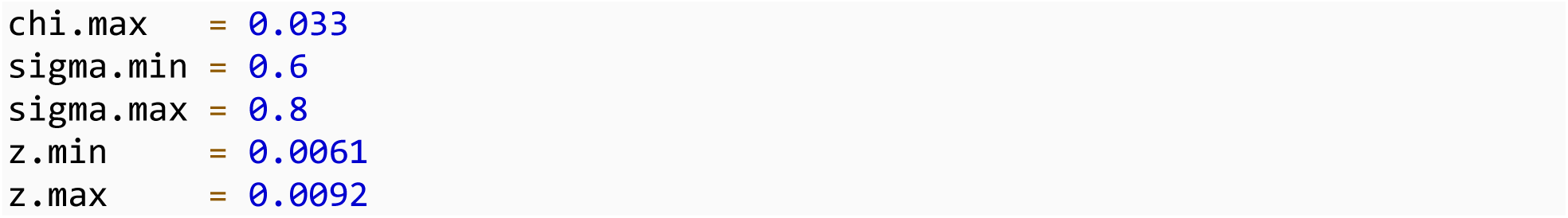

### Parameter set

Generate parameter set for the uncertainty analysis with their assumed distribution

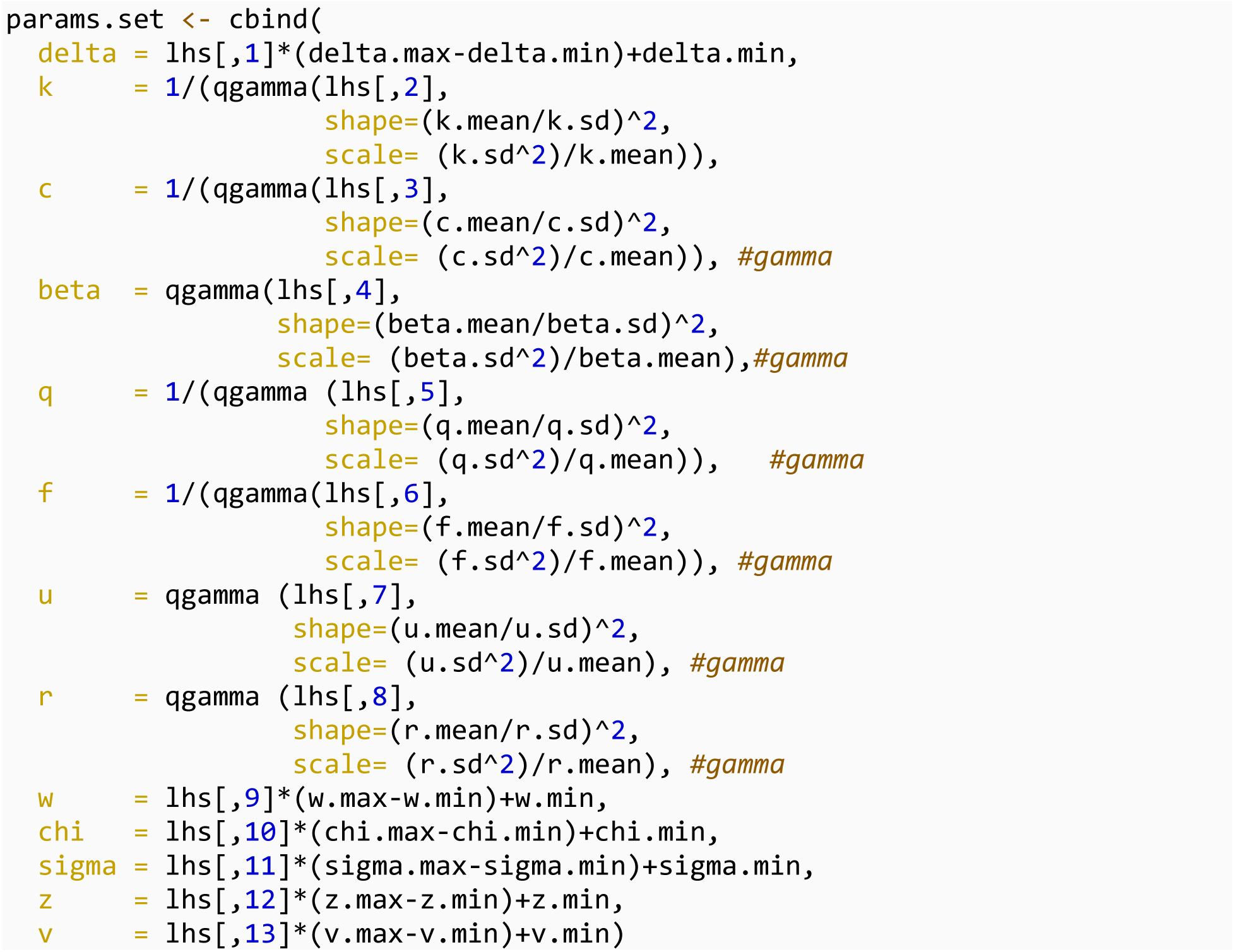

Facilaitate later plotting, consider levels of transmission rate reduction from 500 simulations:

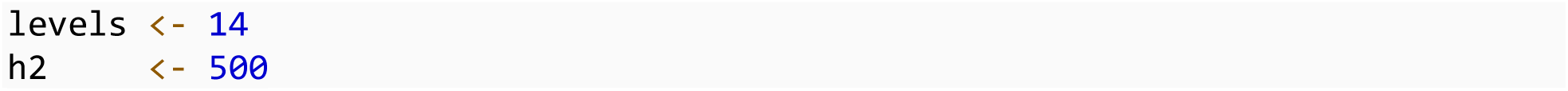

Finally, we set up a nested loop, to cycle through different values of r, then to cycle through different simulated parameter sets. Note the pre-allocated data frame and use of the counter j, also to speed up evaluation.

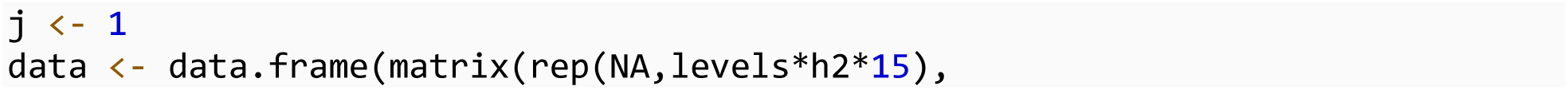
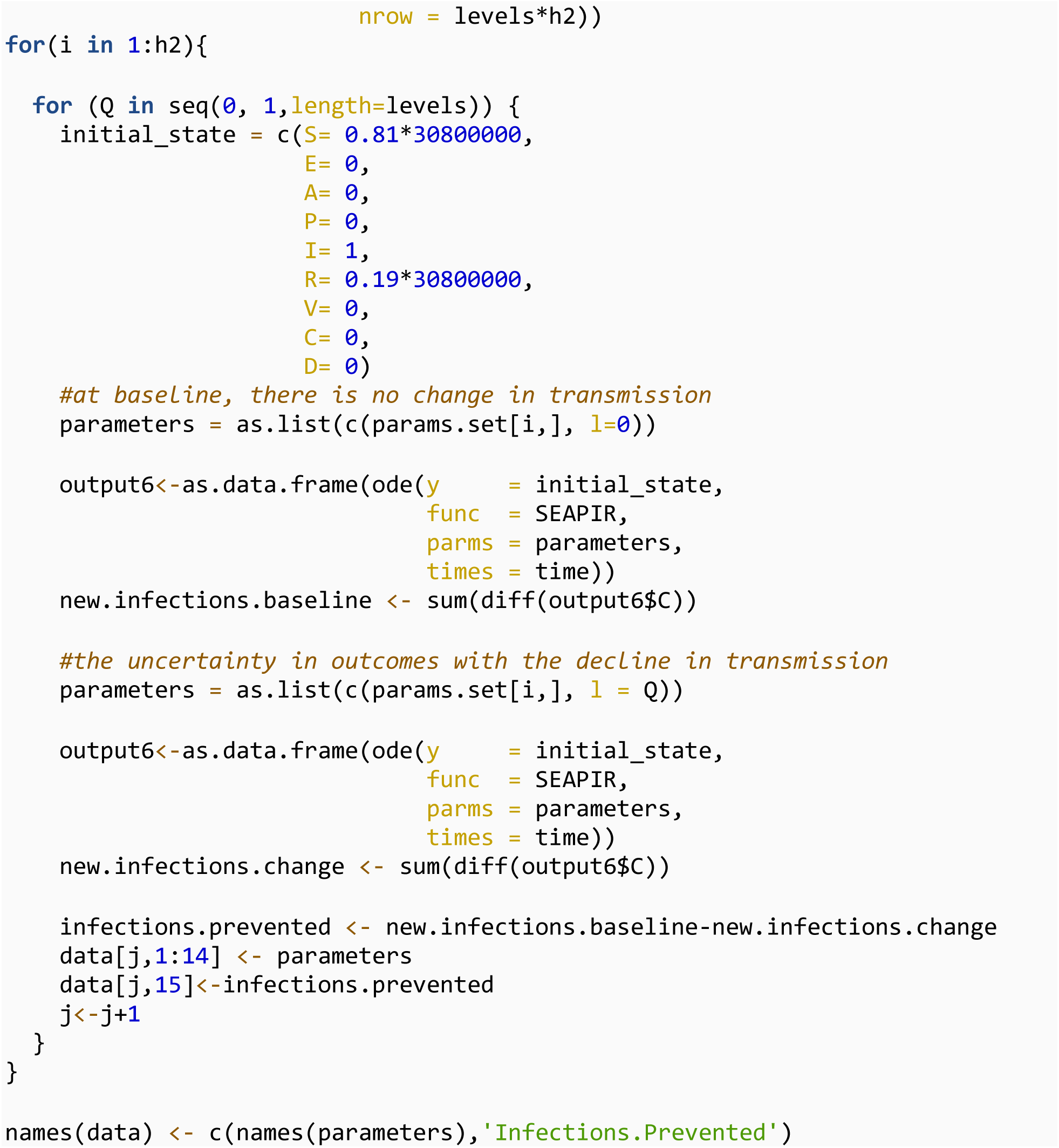

Saving the simulation results from the uncertainty analysis

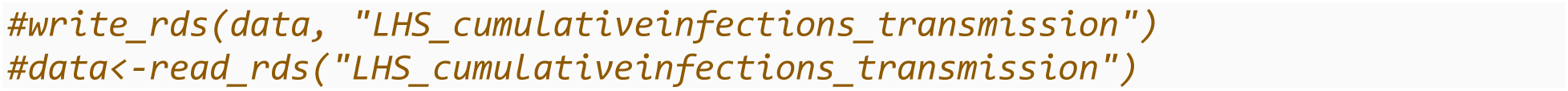

Now we plot the cumulative infections averted at baseline (black line) and uncertainty analysis (box plots)

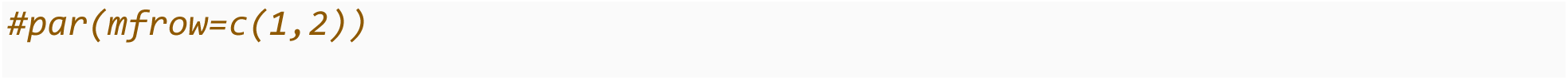
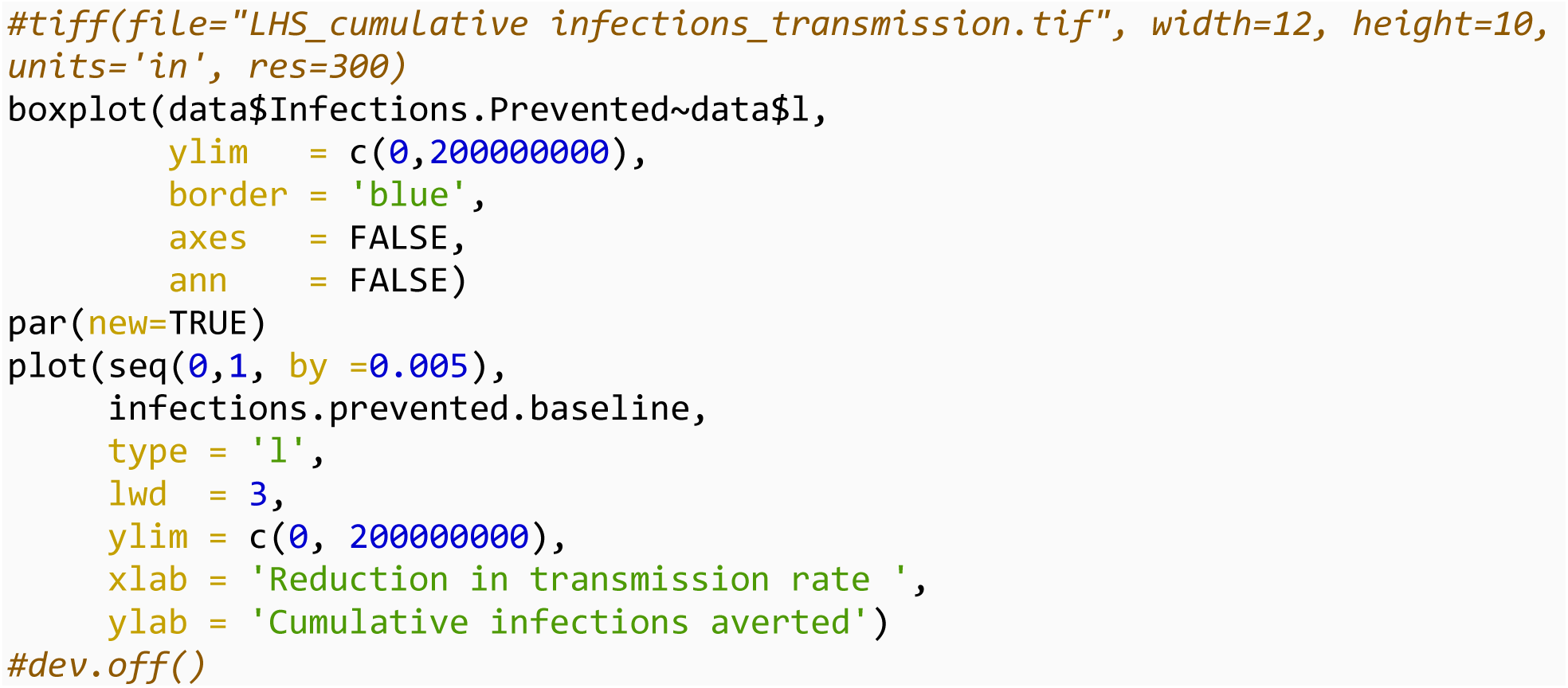

### Sensitivity analysis: Partial rank correlation coefficients

A global sensitivity analysis is performed to identify the parameters the outcome was sensitive or robust to add the sensitivity package to perform sensitivity analysis

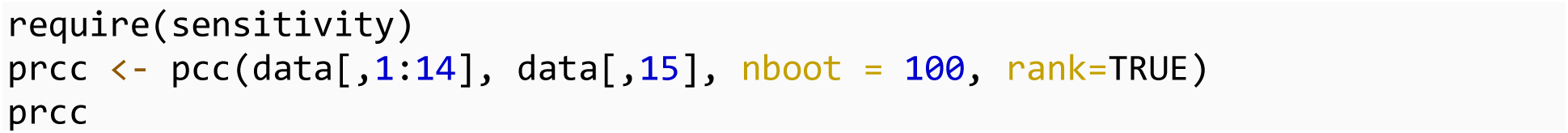

The result for sensitivity analysis is saved as an RDS file

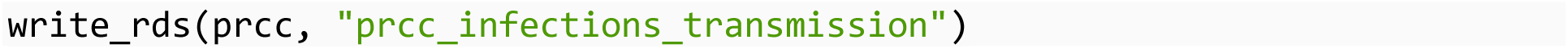

### Increasing vaccine coverage: cumulative infections and deaths averted

Lastly, we estimated the cumulative infections and deaths averted with increasing vaccine coverage. We repeated the codes from decline in transmission, however Q here represented the increase in vaccination rate.

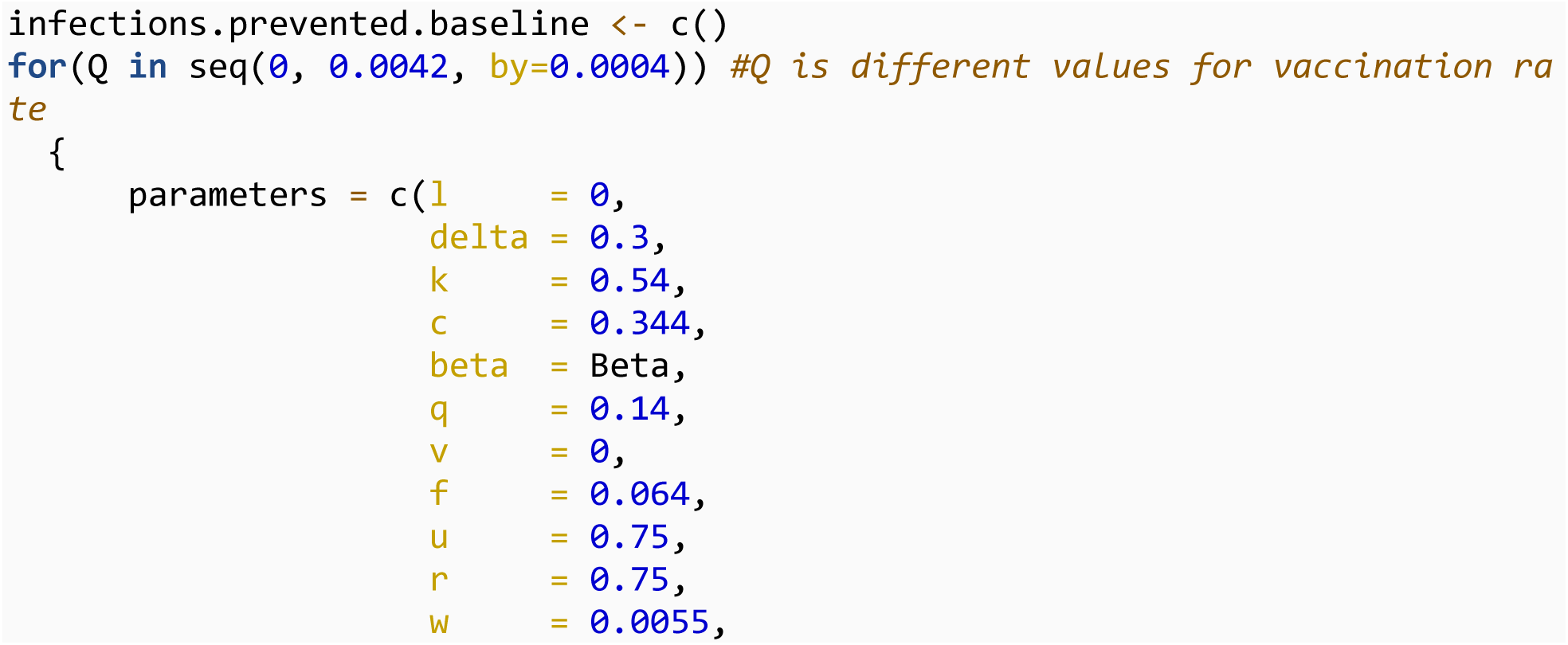
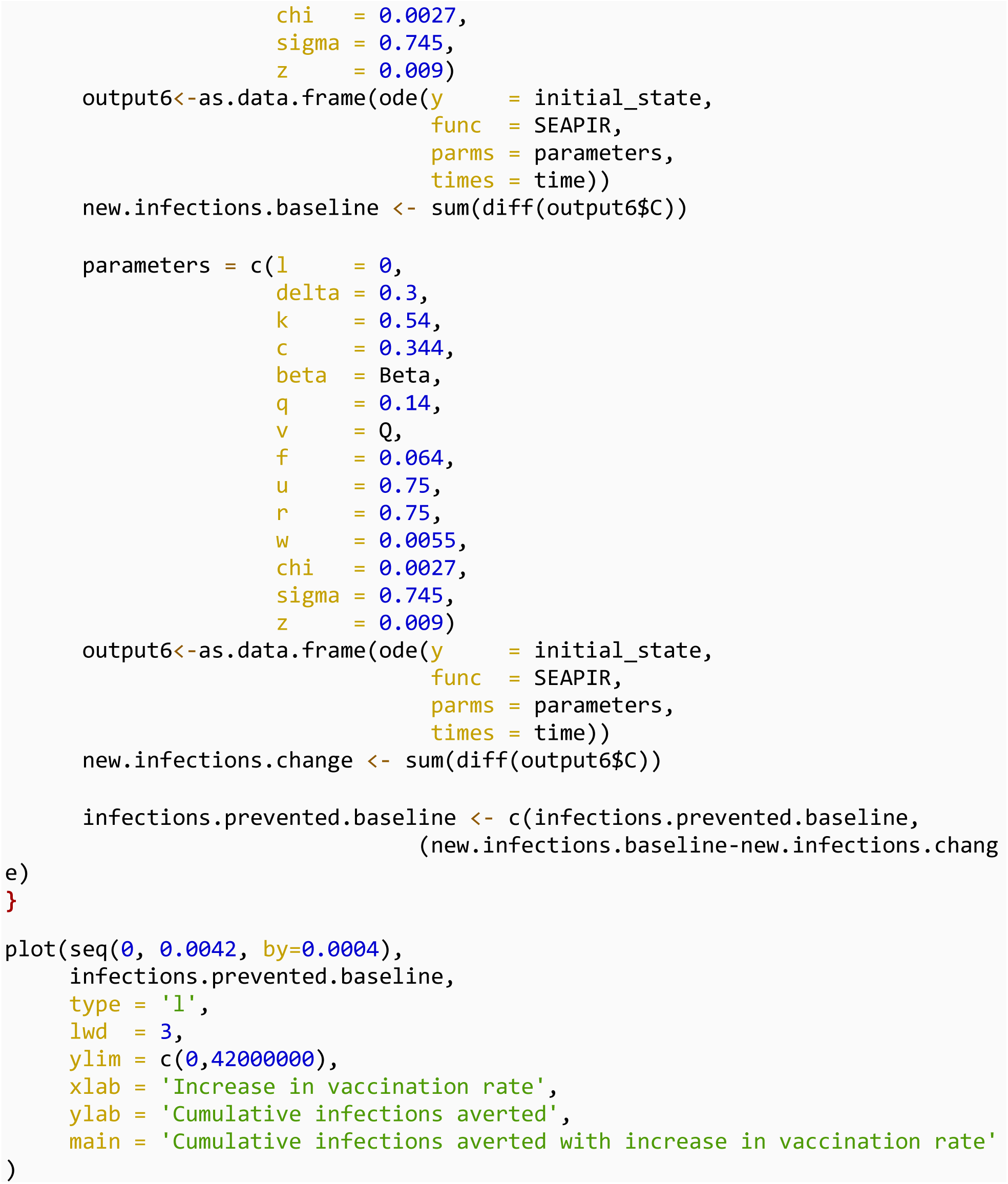

### Latin hypercube sampling

12 uncertain parameters introduces to create Latin hypercube design

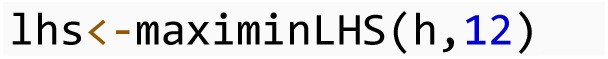

To facilitate plotting later, 14 levels of vaccination rates

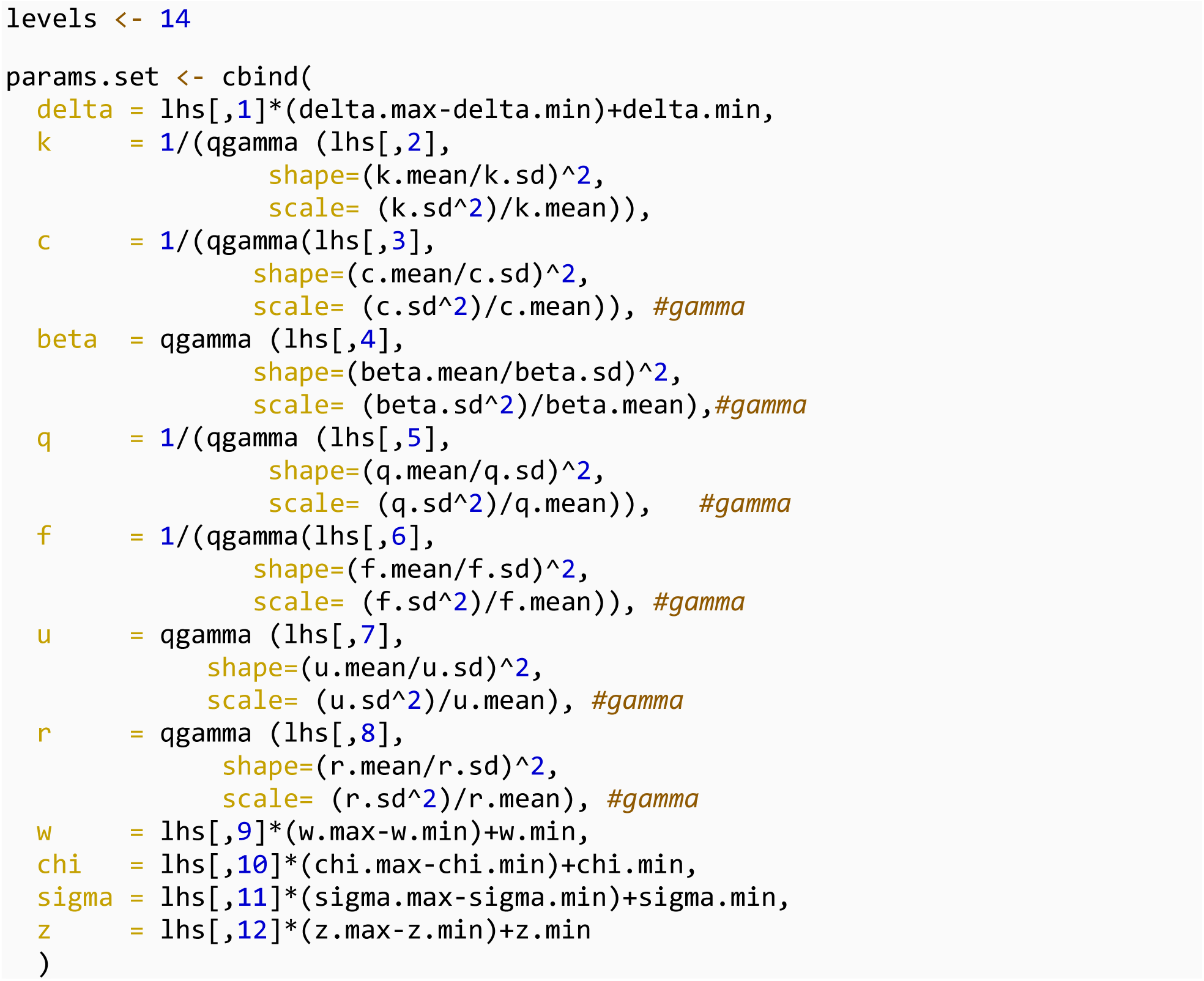

500 simulations were performed

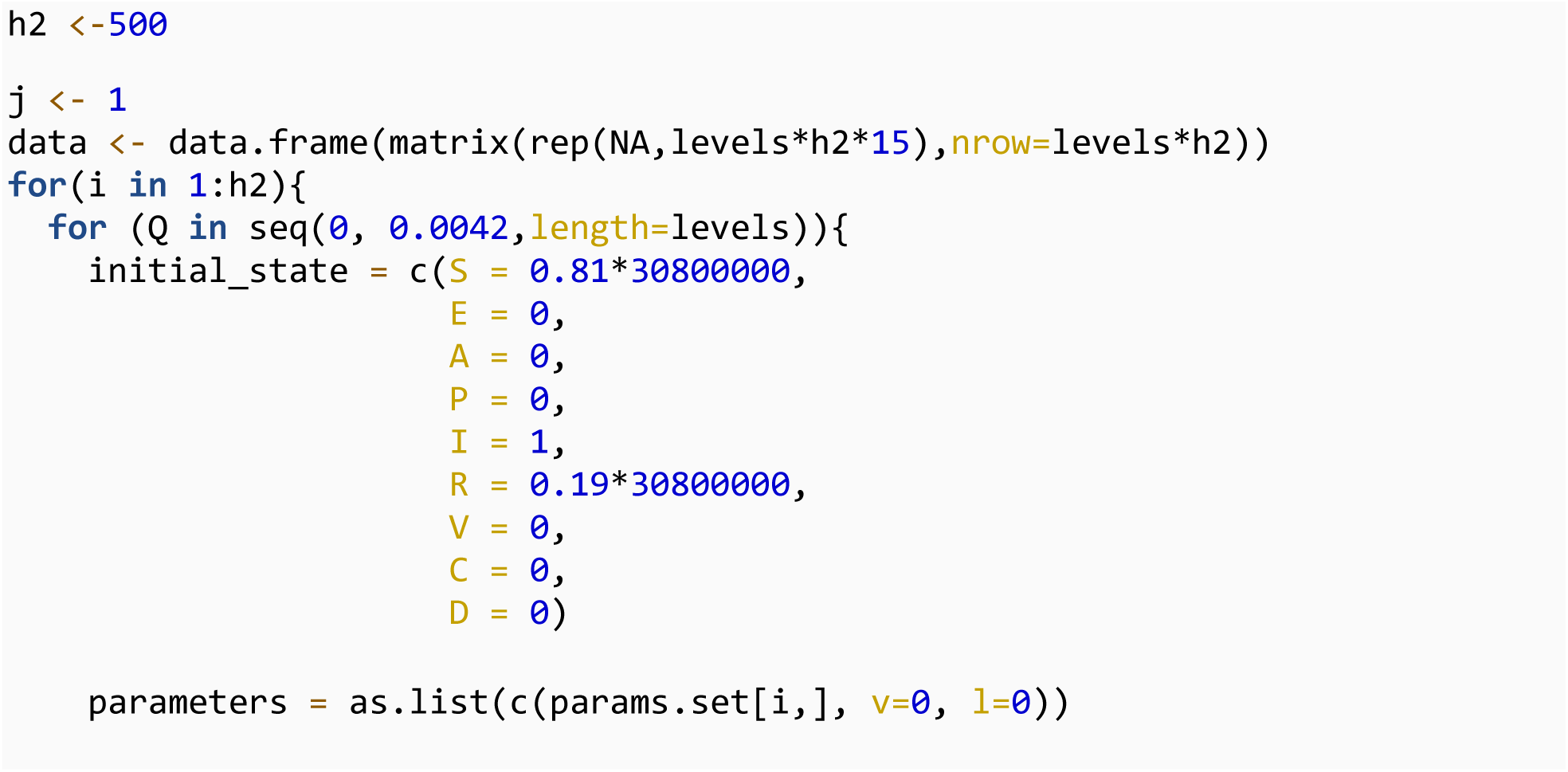
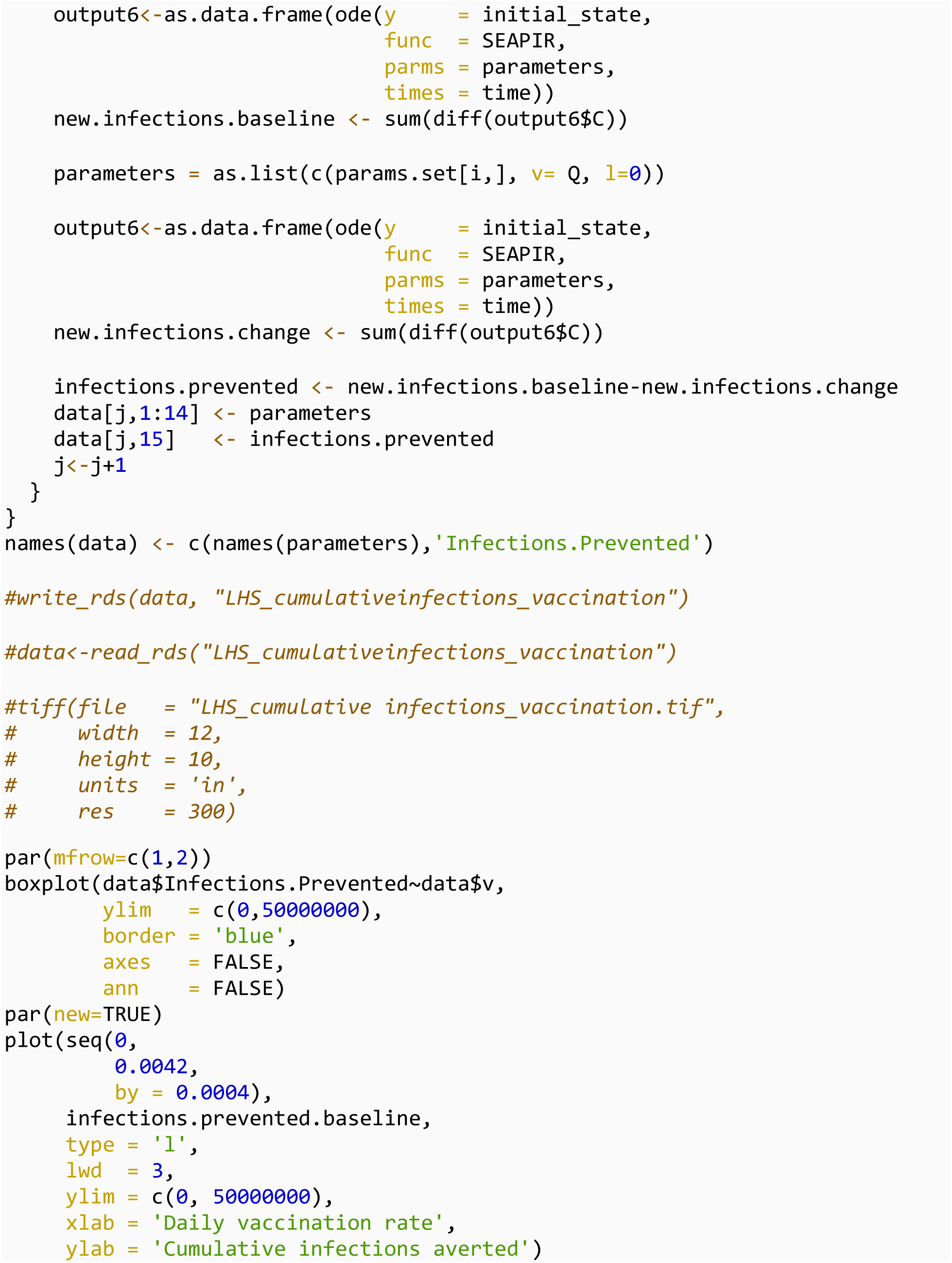
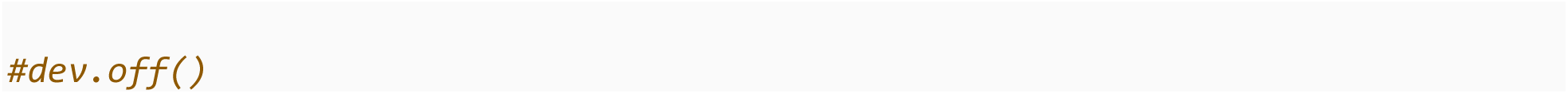

### Sensitivity analysis: Partial rank correlation coefficients

A global sensitivity analysis is performed to identify the parameters the outcome was sensitive or robust to add the sensitivity package to perform sensitivity analysis

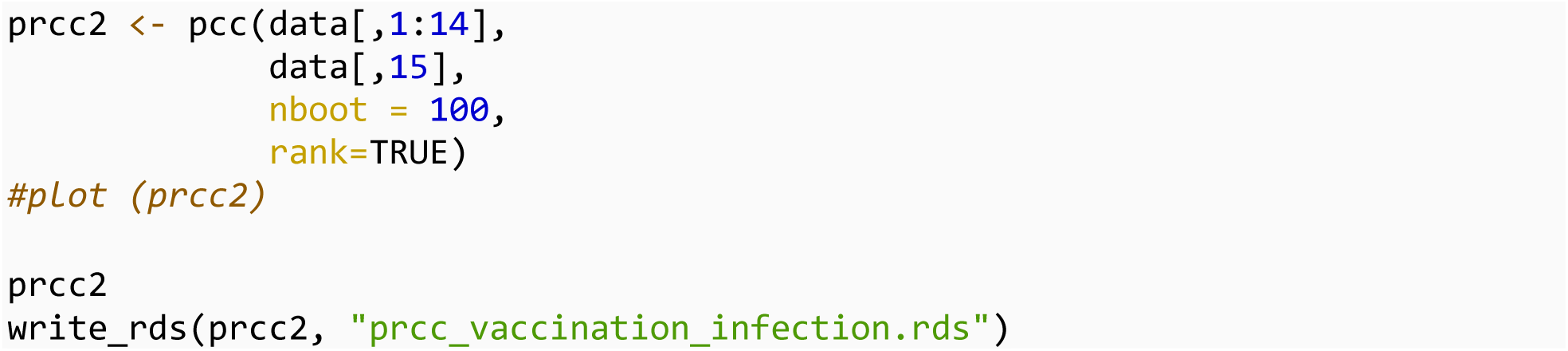

